# Peripheral blood leukocyte signatures as biomarkers in relapsed ovarian cancer patients receiving combined anti-CD73/anti-PD-L1 immunotherapy in Arm A of the NSGO-OV-UMB1/ENGOT-OV30 trial

**DOI:** 10.1101/2024.10.02.24314755

**Authors:** Luka Tandaric, Annika Auranen, Katrin Kleinmanns, René DePont Christensen, Liv Cecilie Vestrheim Thomsen, Cara Ellen Wogsland, Emmet McCormack, Johanna Mäenpää, Kristine Madsen, Karen Stampe Petersson, Mansoor Raza Mirza, Line Bjørge

## Abstract

**Background:** Clinical trials of immune checkpoint inhibitors in epithelial ovarian cancer (EOC) have not shown clear survival benefit, likely due to the complex immunosuppressive mechanisms of the EOC tumor microenvironment. Still, certain patients experience long-term treatment benefit. However, we lack reliable biomarkers for distinguishing dominant immunosuppressive mechanisms and for identifying patients with EOC who are responsive to immunotherapy. The present high-dimensional single-cell study analyzed patients with relapsed EOC enrolled in arm A of the NSGO-OV-UMB1/ENGOT-OV30 trial, wherein the patients underwent combination oleclumab (anti-CD73) and durvalumab (anti-PD-L1) immunotherapy. The objective of the study was the identification of blood-based immunophenotypic signatures conducive to the development of improved strategies for patient selection, response monitoring, and personalized targeting of immunosuppressive mechanisms.

**Methods:** A 40-marker suspension mass cytometry panel was utilized for comprehensive phenotypic and functional characterization of longitudinally sampled peripheral blood leukocytes from patients. Artificial neural network-based unsupervised clustering and manual metacluster curation were used to identify leukocyte subsets for differential discovery and correlation analyses.

**Results:** At baseline, short-term and long-term survivors differed with regard to the relative abundances of total peripheral blood mononuclear cells (PBMCs). We observed a significant increase in CD14^+^CD16^−^ myeloid cells during treatment, initially driven by classical monocyte proliferation and subsequently driven by the expansion of monocytic myeloid-derived suppressor cells (M-MDSCs). This M-MDSC expansion occurred only in patients with shorter progression-free survival, who also showed a continuous decrease in central memory T-cell abundances after baseline. Throughout treatment, we observed upregulation of PD-L1 expression on most T-cell subsets in all patients. Higher expression of CD73 and IDO1 on select leukocyte subsets at baseline was significantly positively correlated with longer progression-free survival.

**Conclusions:** Our study delineates the phenotypic and functional alterations in peripheral blood leukocytes occurring during combination oleclumab/durvalumab immunotherapy in patients with relapsed EOC. We propose a set of biomarkers with potential for treatment personalization and response monitoring: relative abundances of PBMCs at baseline, relative abundances of M-MDSCs and central memory T cells during treatment; PD-L1 expression levels over time; and baseline expression of CD73 and IDO1 on specific leukocyte subsets. However, validation of these biomarkers through larger-scale studies is required.

**KEY MESSAGES:** *WHAT IS ALREADY KNOWN ON THIS TOPIC:* Despite promising preclinical results and moderate efficacy in lung cancer, combination anti-CD73/anti-PD-L1 immunotherapy in relapsed epithelial ovarian cancer (EOC) has shown only modest response rates, consistent with other EOC immunotherapy trials. Durable clinical benefit remains rare, and there are currently no biomarkers available for the effective selection of EOC patients for personalized immunotherapy and for the characterization of sustained responses.

*WHAT THIS STUDY ADDS:* This study demonstrates that, at baseline of combination anti-CD73/anti-PD-L1 treatment, EOC patients with higher relative abundances of peripheral blood mononuclear cells (PBMCs) and elevated expression of CD73 and IDO1 on certain PBMC subsets may experience greater overall survival and progression-free survival benefit, respectively. Additionally, patients with faster disease progression exhibited a shift in CD14^+^CD16^−^ myeloid cells towards a more immunosuppressive phenotype and had lower abundances of central memory T cells over time compared to patients with slower progression, while PD-L1 expression increased significantly over time on T cells of all patients, regardless of the rate of disease progression.

*HOW THIS STUDY MIGHT AFFECT RESEARCH, PRACTICE OR POLICY:* While further clinical validation is required, we propose blood-based biomarkers for predicting and monitoring responses to immunotherapy in EOC, aiming to guide future research on immunotherapy resistance mechanisms and treatment personalization, with a focus on specific peripheral leukocyte subsets.

## INTRODUCTION

Epithelial ovarian cancer (EOC) is predominantly diagnosed at an advanced stage. For the past few decades, surgical cytoreduction followed by carboplatin-paclitaxel chemotherapy has been the default approach to treating EOC [1]. Recently, the clinical implementation of poly(ADP-ribose) polymerase inhibitors (PARPi’s) and the anti-angiogenesis agent bevacizumab as maintenance therapy has significantly improved survival [2]. However, despite most patients initially responding to primary treatment, 70%-80% of the responders eventually experience relapse [3]. Although multiple therapeutics are used to treat recurrent disease, the treatment intent shifts from curative to palliative. Drug resistance and the dearth of effective post-relapse treatment options have resulted in the long-term survival rates remaining low [4] and highlighted the need for novel treatment modalities.

Immunotherapy, particularly the clinical implementation of immune checkpoint inhibitors (ICIs), has improved the outcomes of several cancers by disrupting tumor immune evasion mechanisms, mainly the induction of T-cell exhaustion [5]. Seminal studies by Zhang et al. [6] and Curiel et al. [7], correlating treatment response and survival with the presence, position, and phenotypes of tumor-infiltrating lymphocytes in EOC, have indicated rationale for applying immunotherapy in EOC as well. Although numerous trials on immunotherapeutic regimens in EOC have been conducted, including studies incorporating patient pre-selection strategies based on predictive biomarkers, such as PD-L1 [8], none have shown sufficient efficacy to be considered clinically viable [9]. Immunotherapy has failed to achieve potency in EOC on account of multiple immunoinhibitory mechanisms simultaneously active in its tumor microenvironment (TME). This facilitates a high degree of immunosuppression, unsurmountable by immunotherapeutics delivered either as a monotherapy or in combination with chemotherapeutics [10]. In preclinical studies on EOC, combination immunotherapy regimens have resulted in notable increases in antitumor efficacy over single-agent immunotherapeutics, predominantly by inhibition of PD-1/PD-L1 interaction alongside simultaneous activation of an immunostimulatory receptor on T cells [11].

Building off of promising preclinical results of combination immunotherapies in murine models of solid cancers, which had shown significant synergistic effects in terms of tumor reduction and survival improvement, as well as elevated abundance and anti-tumor activity of CD8^+^ T cells, compared to the corresponding monotherapies [12–14], the non-randomized phase II trial NSGO-OV-UMB1/ENGOT-OV30 was designed as a three-arm umbrella trial for testing the clinical efficacy of several novel combinations of immunotherapeutics in patients with relapsed EOC [15]. The treatment in arm A comprised oleclumab and durvalumab - human monoclonal IgG1 antibodies that target and inhibit CD73 and PD-L1, respectively. CD73, an adenosine-generating extracellular enzyme present on most normal tissues [16], is overexpressed in most cancers [17] and associated with poor prognosis in EOC [18]. Binding of adenosine to A2A receptors on antigen-activated T cells results in activation blockage and exhaustion [19]. Accordingly, inhibition of the CD73/A2A receptor axis is considered an effective means of re-invigorating anti-tumor immunity [20]. Furthermore, simultaneous inhibition of CD73 and the PD-1/PD-L1 interaction in preclinical *in vitro* and *in vivo* human cell line models was shown to have a synergistic anti-tumor effect compared to the individual inhibition of either [21]. Based on these results, the oleclumab/durvalumab combination was approved for testing in a phase II clinical trial setting [15]. While it demonstrated only marginal clinical efficacy in a relapsed EOC cohort, a number of patients exhibited prolonged survival.

This translational study presents the results of the high-dimensional single-cell suspension cytometry by time-of-flight (CyTOF) analysis of blood samples collected from trial cohort A prior to treatment and at regular intervals during treatment. The aim was the elucidation of clinically actionable phenotypic signatures of the response of relapsed EOC to oleclumab/durvalumab immunotherapy. We identified potential leukocyte-based predictive, prognostic, and response biomarkers for the purposes of improved patient selection, preemptive detection of cancer progression, and portrayal of *in vivo* drug activity, respectively.

## METHODS

### Patient cohort and samples

In the international NSGO-OV-UMB1/ENGOT-OV-30 umbrella trial, 25 immunotherapy-naïve patients with relapsed EOC who had CD73-positive archival tumor samples were included in arm A and intravenously administered 3000 mg oleclumab and 1500 mg durvalumab every two and four weeks, respectively (figure 1A). The primary endpoint was response evaluation (disease control rate) at the 16-week timepoint. Peripheral blood samples for single-cell suspension CyTOF were collected in 10 mL EDTA Vacutainer tubes (Becton Dickinson, REF 367525) at baseline (pre-treatment), at minimum every two 28-day cycles, and at end of trial or at disease progression. Within one hour of collection, samples were processed using BD Phosflow Lyse/Fix buffer (Becton Dickinson, REF 558049) and frozen at −80°C (figure 1B,C) (protocol described in the online supplemental appendix). Samples were stored in the biobank of Haukeland University Hospital, Bergen, Norway.

**Figure 1.**
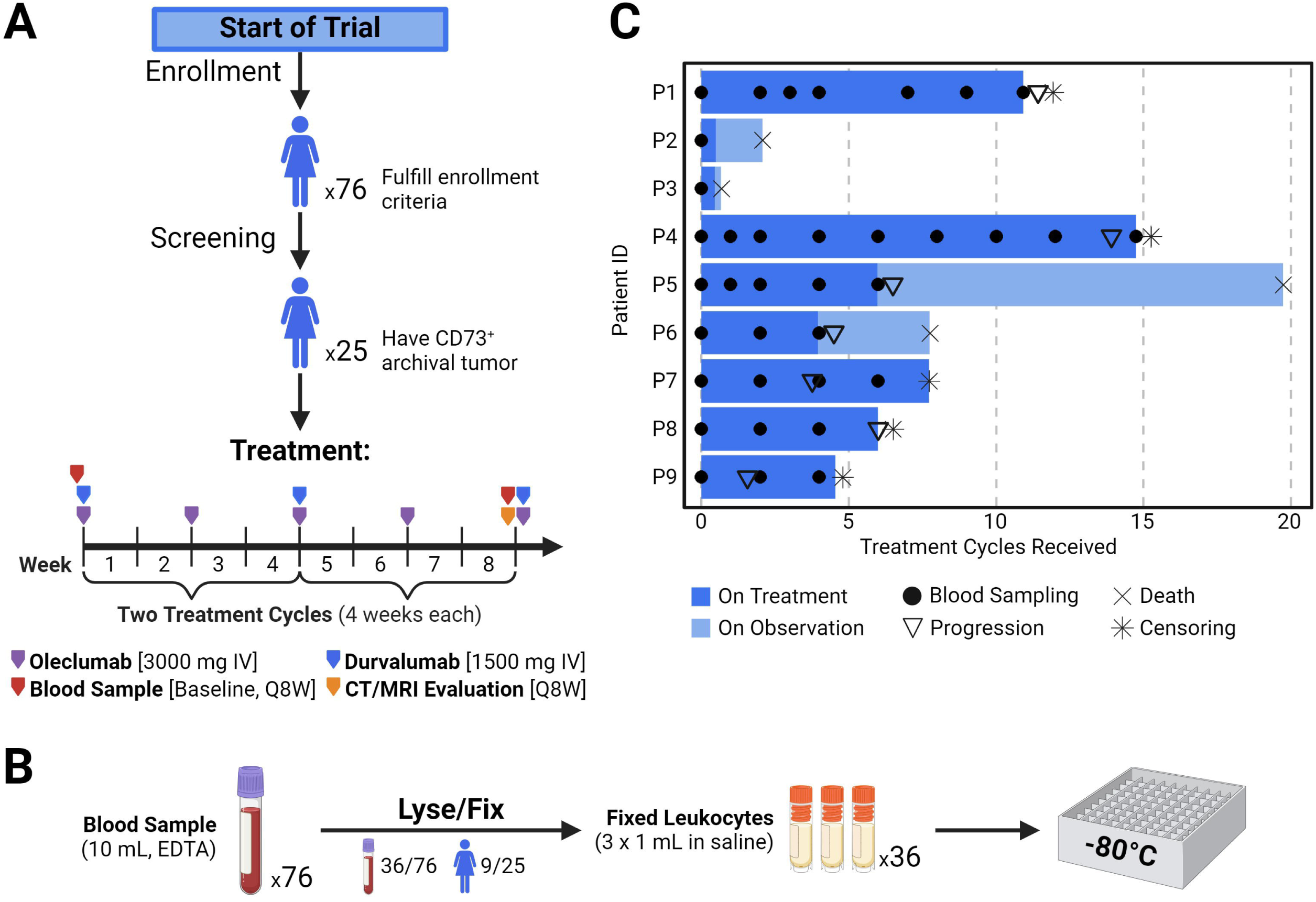
Trial overview, sample collection and processing. (A) Schema of the patient disposition and immunotherapy administration schedule of the NSGO-OV-UMB1/ENGOT-OV30 trial. One treatment cycle had a duration of four weeks. (B) Blood sample acquisition and processing workflow (full protocol provided in the online supplemental appendix). (C) Swimmer plot illustrating the duration of treatment and observation for each patient included in this study, as well as blood sampling timepoints. IV – intravenous

### Mass cytometry panel development

A 40-marker antibody panel (online supplemental table S1) encompassing the entire human peripheral blood leukocyte repertoire was designed and developed in-house for in-depth single-cell analysis of the leukocyte samples. As the aim of oleclumab/durvalumab immunotherapy was the reinvigoration of T cells from an exhausted state, a central objective of the panel was ascertainment of information about T-cell states before and during treatment. Given that myeloid cells in the peripheral blood have been observed to significantly influence the outcome of immunotherapy of solid cancers [22], as well as have a crucial influence on treatment outcome upon settlement into the TME of EOC [23], another objective of the panel was the characterization of the myeloid cells in the samples.

For antibody clones not commercially available in a pre-metal-conjugated format, 100 µg of purified, carrier-protein-free antibody was conjugated in-house to cadmium- or lanthanide-loaded polymers using Maxpar MCP9 or X8 antibody labeling kits, respectively, according to the manufacturer’s protocol (Standard BioTools).

Antibodies were titrated on leukocytes from whole blood collected from healthy female donors and processed using the Lyse/Fix buffer. Titration of lineage and cell state markers was performed on unstimulated leukocytes, and peripheral blood mononuclear cells (PBMCs) stimulated *in vitro* by interleukin-2 (IL-2) (Sigma, Cat.No. I2644) and phytohemagglutinin-P (PHA-P) (Sigma, Cat.No. 61764), respectively, according to the protocol in the online supplemental appendix. To ensure consistency, identical sample processing and staining protocols were maintained for the titration samples and clinical samples. The optimal concentration of Cell-ID Intercalator-Ir (Standard BioTools, Cat.No. 201192B) for the identification of nucleated cells was determined to be 250 nM by titration. The panel was validated for use on the CyTOF XT mass cytometer (Standard BioTools).

### Sample staining and data acquisition

To facilitate uniform staining, batches containing up to 20 samples were designed, wherein a unique palladium-based tag was assigned to each sample. For batch effect correction, each barcoded batch contained two different anchor samples consisting of Lyse/Fix-treated healthy donor leukocytes, one of which had an admixture of PHA-P/IL-2-treated healthy donor PBMCs to enable batch correction of activation/exhaustion markers. Frozen fixed leukocyte samples were thawed and treated with 0.25 mg/mL DNAse I (Sigma, Cat.No. DN25) dissolved in Dulbecco’s phosphate-buffered saline containing Ca^2+^ and Mg^2+^ (Sigma, Cat.No. D8662) (henceforth referred to as “DNAse”). Cells were counted, and a maximum of 3.5×10^6^ cells from each sample were tagged using a palladium-based barcoding method according to the manufacturer’s protocol (Standard BioTools, Cat.No. 201060). Barcoding reagents were washed away, and all cells within a batch were pooled, counted, and frozen at −80°C.

To maintain consistency in the composition of staining antibody mixes across sample batches, two mixes of antibodies, targeting either surface or intracellular markers, were pre-made in quantities large enough to stain the full set of samples, aliquoted, and stored at −80°C. Comparative testing of frozen versus freshly-made antibody mixes on Lyse/Fix-treated healthy donor leukocytes demonstrated that freezing did not affect the effectiveness or specificity of the panel antibodies.

For staining, barcoded sample mixes were thawed and counted. Cells were then incubated in MaxPar Cell Staining Buffer (CSB) (Standard BioTools, Cat.No. 201068) containing human FcR Blocking Reagent (Miltenyi Biotec, Cat.No. 130-059-901), and heparin (Ratiopharm, Reg.No. 5394.00.00) to prevent non-specific antibody binding. Aliquots of the antibody mix for staining cell surface proteins were thawed and added to the cells at 3×10^6^ cells per 100 µL of staining volume. After incubating for 45 minutes at room temperature (RT), superfluous antibody was washed away with MaxPar CSB, then with MaxPar phosphate-buffered saline (PBS) (Standard BioTools, Cat.No. 201058), after which cells were permeabilized by a 10-minute incubation in pure methanol (Sigma, Cat.No. 32213) at −20°C. The methanol was washed away with MaxPar PBS, followed by MaxPar CSB. Aliquots of the antibody mix for targeting intracellular proteins were thawed and applied to the permeabilized cells for 30 minutes at RT, at a concentration of 3×10^6^ cells per 100 µL of staining volume. For DNA staining, washed cells were incubated with iridium intercalator diluted in MaxPar PBS containing 4% V/V formaldehyde for 20 min at RT. The cells were washed, resuspended in a 10% V/V solution of dimethyl-sulfoxide (Sigma, Cat.No. W387520) in fetal bovine serum (Sigma, Cat.No. F7524) and frozen at −80°C.

For data acquisition, fully stained cells were thawed, washed using MaxPar CSB, and incubated in DNAse. Cells were then washed and pelleted in MaxPar Cell Acquisition Solution (Standard BioTools, Cat.No 201240), kept on ice until data acquisition, and acquired in MaxPar Cell Acquisition Solution Plus (Standard BioTools, Cat.No. 201244). High-dimensional data from three barcoded batches was acquired on a CyTOF XT mass cytometer at ∼400 events per second, with EQ Six-Element Calibration Beads (Standard BioTools, Cat.No. 201245) present in the suspension at a 1:10 dilution (detailed barcoding and staining methodologies provided in the online supplemental appendix).

### Data pre-processing

Bead-based longitudinal signal intensity normalization was done automatically by the CyTOF XT software upon data acquisition. Events were de-barcoded using the MATLAB Compiler Runtime (R2013a(8.1)) implementation of The Single Cell Debarcoder by Zunder et al. [24]. De-barcoded events were uploaded to Cytobank (Beckman Coulter Inc.), where irregular events were removed by manual gating (online supplemental figure S1). The resulting single-cell events were batch corrected with the R-based GUI implementation of CytofBatchAdjust by Schuyler et al. (v0.0.0.9001) [25], using the composite anchor sample of each batch. Batch correction effectiveness was assessed by analyzing the corrected data of the alternate anchor sample.

### Clustering and differential analyses

Unsupervised clustering, dimensionality reduction, manually curated metaclustering, and differential analyses were performed using the “flowSOM” [26], “umap” [27], “CATALYST” [28], and “diffcyt” [29] R packages, respectively.

Total leukocyte data from all patient samples was first clustered into a 64-cluster (8×8) self-organizing map (SOM) and manually metaclustered into granulocyte, PBMC, and debris subsets based on the expression of 13 markers (figure 2A). The assignment of cell identity was rigorously quality checked using biaxial plots, heatmaps, UMAPs, and hierarchical clustering of clusters. The PBMC subset was further sub-clustered into a 256-cluster (16×16) SOM and manually grouped into 24 subsets corresponding to widely established PBMC types using 22 markers (figure 2B). Subset assignment was validated in the same manner as that for total leukocytes.

**Figure 2.**
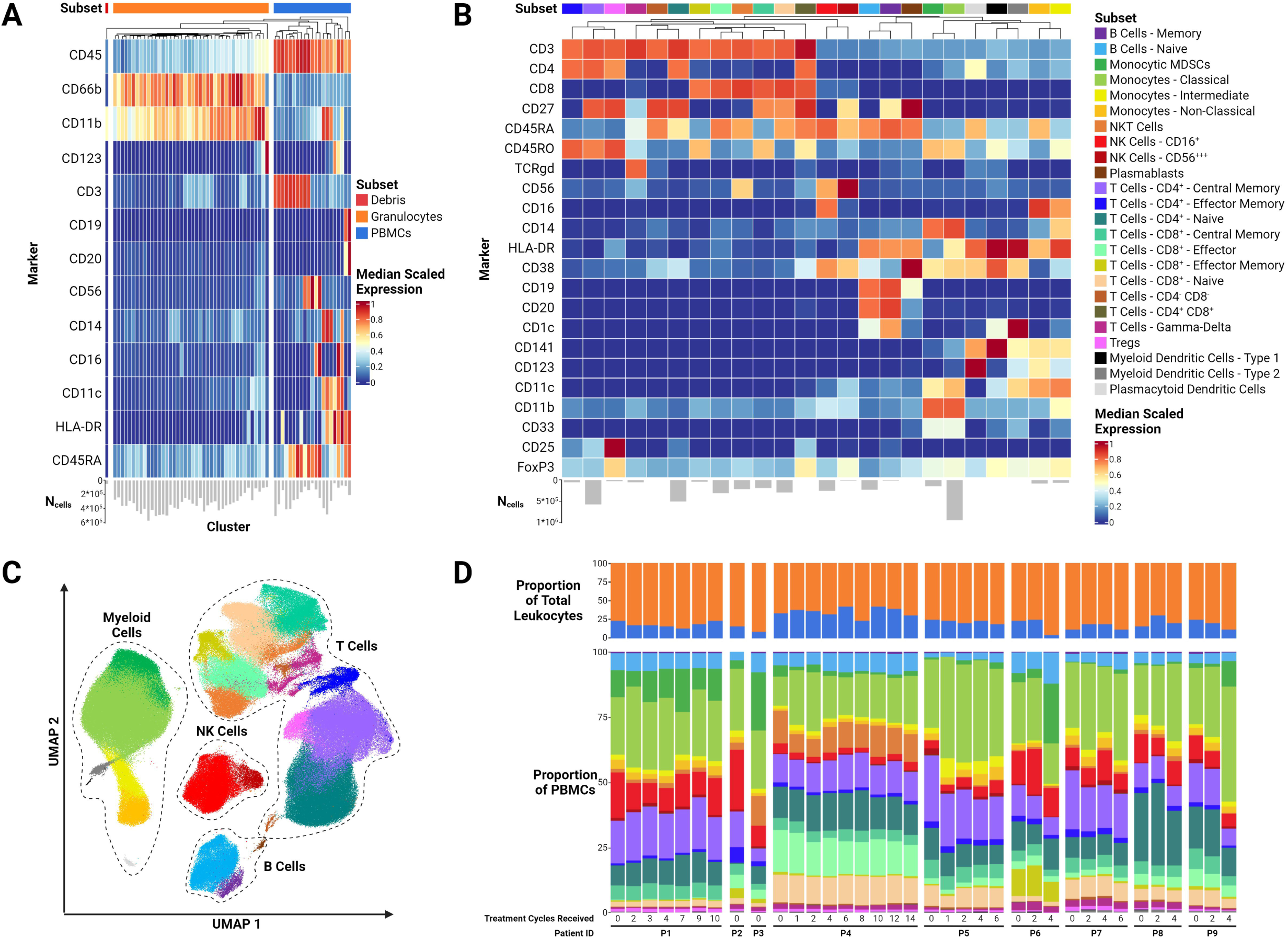
Formation and overall view of the study dataset. (A) Heatmap of the normalized median expression of the 13 markers (rows) used for the clustering of total leukocytes. Each column represents a cluster. Appropriateness of leukocyte subset assignment* was confirmed by unsupervised hierarchical clustering (visualized above heatmap). (B) Heatmap of the normalized median expression of the 22 markers (rows) used for the clustering of PBMCs. Each column represents a leukocyte subset. Subset assignment* was performed and validated in the same manner as for the total leukocyte clusters. (C) UMAP projection of an aggregated sample of PBMCs, consisting of an equal number of cells sampled from each blood sample. Each cell is colored by subset assignment*. (D) Overview of the leukocyte (upper stacked bar plots) and PBMC (lower stacked bar plots) composition of all blood samples (detailed composition of each sample is given in online supplemental table S2). *Leukocyte subset assignment coloring is consistent throughout figure 2. TCRgd - gamma-delta T-cell receptor

### Statistical analyses

Differential subset abundance and cell state marker expression between clinical sample groups, stratified by length of survival, duration of progression-free survival (PFS), or sampling timepoint, were analyzed using the edgeR and limma functions of the diffcyt R package, respectively. Multiple hypothesis testing correction was conducted using the Benjamini-Hochberg method, giving adjusted p-values (p_adj_).

To identify potential predictive biomarkers among baseline leukocyte characteristics, the relationships between PFS duration, and either relative cell subset abundances or state marker expression medians for each cell subset were analyzed using GraphPad Prism (v10.2.0, GraphPad Software). Given that patients in the short-term survivor group had died prior to the first tumor evaluation timepoint, this analysis was restricted to the seven patients in the long-term survivor group. According to the results of Shapiro-Wilk normality testing (alpha=0.05), either a Pearson or Spearman correlation test was performed for each marker-subset combination (dataset) alongside simple linear regression to assess the relationship between PFS duration and relative abundances or expression medians. The p-values in the correlation analyses were not corrected for multiple hypothesis testing due to the small size of datasets (n=7) and the discovery-driven nature of this study. To maintain robustness, we considered significant correlations with an absolute correlation coefficient of at least 0.5 and an R-squared value of at least 0.75 as meaningful.

## RESULTS

### Study cohort demographics

In this retrospective translational study, we analyzed total leukocytes longitudinally sampled from patients with relapsed EOC undergoing combination oleclumab/durvalumab immunotherapy (figure 1A) with the aim of identifying predictive, prognostic, and response biomarkers for the administered treatment. Out of the 25-patient trial cohort, 16 patients were not eligible for this study due to samples either missing or being of inadequate quality (figure 1B). Therefore, the data in this study stems from 36 leukocyte samples of nine patients (figure 1C). Nonetheless, our study cohort is highly representative of the clinical trial cohort [15], as the patient age range and distributions of FIGO stage, ECOG status, and clinical response are similar (table 1). In contrast to the patients in the clinical trial, the majority of the patients included in this study had undergone more extensive treatment prior to inclusion.

**Table 1.** Characteristics of the included patients.

| Characteristic | No. of patients (n=9) |
| --- | --- |
| Age, years (%) |  |
| Median (range) | 66 (47-74) |
| 40-49 | 1 (11.1) |
| 50-59 | 1 (11.1) |
| 60-69 | 3 (33.3) |
| ≥70 | 4 (44.4) |
| FIGO stage at initial diagnosis (%) |  |
| I | 1 (11.1) |
| II | 0 (0.0) |
| III | 3 (33.3) |
| IV-A | 4 (44.4) |
| IV-B | 1 (11.1) |
| ECOG PS at baseline (%) |  |
| 0 | 3 (33.3) |
| 1 | 6 (66.7) |
| Prior treatment lines (%) |  |
| 2 | 3 (33.3) |
| ≥3 | 6 (66.7) |
| Treatment cycles received* (%) |  |
| Median (range) | 6 (1-15) |
| ≤4 | 4 (44.4) |
| >4 | 5 (55.6) |
| Time to progression (weeks) (%) |  |
| ≤16 | 3 (33.3) |
| >16 | 4 (44.4) |
| N/A** | 2 (22.2) |
| Best clinical response (%) |  |
| CR | 0 (0.0) |
| PR | 1 (11.1) |
| SD | 5 (55.6) |
| PD | 1 (11.1) |
| N/A** | 2 (22.2) |

\*One cycle corresponds to four weeks of treatment.
\*\* Patients passed away prior to the first post-baseline tumor evaluation.
FIGO - International Federation of Gynecology and Obstetrics
ECOG PS - Eastern Cooperative Oncology Group performance status

### CyTOF analysis enabled in-depth characterization of patients’ leukocyte profiles

We utilized single-cell suspension CyTOF for high-dimensional phenotypic profiling of patient leukocytes. Approximately 21 million total events were acquired from the 36 leukocyte samples. After quality controls, single-cell data of about 19 million leukocytes remained for further analysis. After batch correction, unsupervised clustering using FlowSOM and manual metaclustering of the total leukocytes produced three leukocyte subsets - granulocytes, PBMCs, and debris (figure 2A). To achieve the intended analytical depth, the same clustering and metaclustering procedures were applied in higher resolution to the PBMCs, resulting in the clear delineation of 24 PBMC subsets (figure 2B). The robustness of the PBMC (meta)clustering was confirmed by using the UMAP dimensionality reduction algorithm on an aggregated dataset comprising 10,000 PBMCs from each of the 36 samples (figure 2C).

All PBMC subtypes, except for the rarest populations (plasmablasts, CD141^+^ myeloid dendritic cells, and CD4^+^CD8^+^ T cells), were detected in all samples (figure 2D, online supplemental table S2). Inspection of the complete dataset revealed that each patient’s leukocyte profile remained largely consistent over time, but inter-patient heterogeneity was markedly more pronounced (figure 2D). Nevertheless, certain trends in compositional changes of leukocyte profiles over time were observed, such as the expansion of myeloid cells at the cost of overall T-cell proportions in nearly all patients. To determine the significance of these observations for biomarker discovery, we performed differential analyses of leukocyte composition and cell state marker expression.

### Long-term survivors exhibit larger abundances of PBMCs at baseline

To identify clinically relevant predictive biomarkers, patients were first stratified into short-term survivors (≤16 weeks, n=2) and long-term survivors (>16 weeks, n=7). We observed that long-term survivors had, on average, significantly higher relative abundance of total PBMCs at baseline (p_adj_=0.0279) (figure 3A,B). No significant differences were observed in the relative abundances of other cell subsets between the groups at baseline. As the short-term survivors provided only baseline samples, we could not compare other timepoints between the groups.

**Figure 3.**
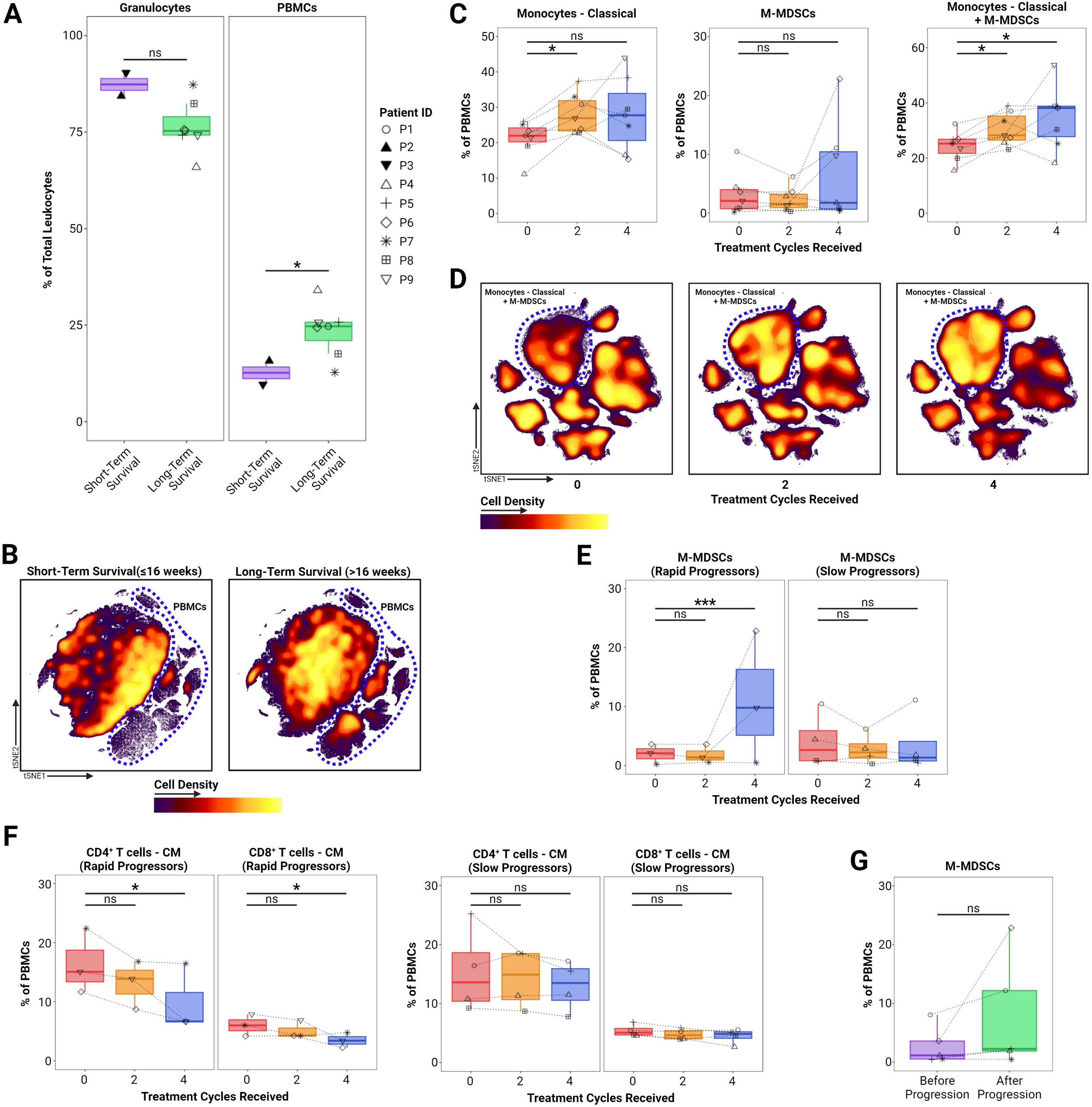
Significant differences, changes, and trends in the relative abundances of leukocyte populations. (A) Comparison of the relative baseline abundances of granulocytes and total PBMCs between survival duration groups (short-term survival is defined as observed survival of at most 16 weeks since the start of treatment; long-term survival is defined as observed survival of more than 16 weeks since the start of treatment). Symbols representing each patient are consistent throughout all figures. (B) UMAP projections of aggregated baseline samples of total leukocytes stratified by survival duration group. Both projections consist of the same number of cells. The significant difference in the density of the total PBMC subset between groups is accented with a dashed blue outline. (C) Changes in the relative abundances of classical monocytes and monocytic myeloid-derived suppressor cells (M-MDSCs) during treatment in the group of long-term survivors (n=7). The two PBMC subsets composed of CD14^+^CD16^−^ myeloid cells (Monocytes - Classical and M-MDSCs), are shown separately in the left and middle panel. The level of HLA-DR expression forms the basis for the separation of classical monocytes (HLA-DR^+/hi^) from M-MDSCs (HLA-DR^−/lo^) (online supplemental figure S2). In the right panel, the classical monocyte and M-MDSC subsets were merged to illustrate the significant and continued increase of the abundance of CD14^+^CD16^−^ myeloid cells during treatment. (D) UMAP projections of aggregated PBMC samples from the long-term survivors (n=7), stratified by treatment timepoint. Cells of the “Monocytes-Classical” and “M-MDSCs” subsets are delineated with a blue outline. (E) Changes in the relative abundances of M-MDSCs during treatment. Patients were stratified into rapid progressors (progression confirmed after at most 16 weeks since the start of treatment) and slow progressors (progression confirmed after more than 16 weeks since the start of treatment). (F) Changes in the relative abundances of central memory (CM) T-cell subsets during treatment. Patients were stratified into rapid- and slow progressors. (G) Comparison of the relative abundances of M-MDSCs in samples collected at blood sampling timepoints immediately before and after confirmation of disease progression. ns - not significant; * - p_adj_ < 0.05; *** - p_adj_ < 0.001

### Oleclumab/durvalumab treatment was accompanied by significant CD14^+^CD16^−^ myeloid cell expansion

To ensure consistency and sufficient sample size for adequate statistical power when analyzing longitudinal changes, we evaluated only the samples from long-term survivors taken at baseline (n=7) and after two (n=7) and four treatment cycles (n=7). Compared to baseline, the average frequency of classical monocytes (CD14^+^CD16^−^HLA-DR^+/hi^) in the peripheral blood samples significantly increased (p_adj_=0.0248) after two treatment cycles and remained elevated even after four treatment cycles, although not significantly (p_adj_=0.299) (figure 3C, left panel). Although there were no significant differences in the relative abundances of monocytic myeloid-derived suppressor cells (M-MDSCs) (CD14^+^CD16^−^HLA-DR^−/lo^) over time when analyzed separately from classical monocytes (figure 3C, middle panel), removing the division between classical monocytes and M-MDSCs based on HLA-DR expression (online supplemental figure S2) revealed a significant expansion of CD14^+^CD16^−^ myeloid cells relative to baseline. This expansion was evident after two treatment cycles (p_adj_=0.0165) and persisted even after four treatment cycles (p_adj_=0.0231) (figure 3C, right panel). We visualized this progressive increase in abundance by using UMAP to generate density-colored projections of aggregate samples of PBMCs from each timepoint (figure 3D).

### Myeloid MDSC expansion and central memory T-cell contraction are tied to progression onset

All seven patients in the long-term survivor group experienced tumor progression and were subsequently categorized either as rapid progressors (n=3), who experienced disease progression within the first 16 weeks (four cycles) of treatment, or slow progressors (n=4), who had disease progression more than 16 weeks from the start of treatment. Differential abundance analyses showed significant expansion of the M-MDSC subset after four treatment cycles in the rapid progressors (p_adj_=6.89×10^−8^) (figure 3E, left panel), with no significant longitudinal changes in subset abundance occurring in the slow progressors (figure 3E, right panel). Furthermore, among rapid progressors, we observed a consistent decline, leading to a statistically significant contraction, of both CD4^+^ (p_adj_=0.0477) and CD8^+^ (p_adj_=0.0477) central memory (CD27^+^CD45RO^+^CD45RA^−^) T-cell (T_CM_) subsets after four treatment cycles (figure 3F, two left panels). This trend was not observed among the slow progressors (figure 3F, two right panels).

For five out of the seven long-term survivors, samples from before and after confirmation of progression were available. We noted a marked trend towards increased abundance of M-MDSCs following progression (figure 3G); however, this did not reach statistical significance (p_adj_=0.0537).

### PD-L1 expression is upregulated on T cells during oleclumab/durvalumab treatment

We subsequently performed differential analyses of cell state marker expression levels to investigate inter-group differences and longitudinal changes in the activation, exhaustion, or terminal differentiation of leukocytes, as well as in the activity of signaling pathways potentially influenced by inhibition of adenosinergic signaling [30]. Differential analyses of state marker expression levels between treatment timepoints and baseline in the long-term survivors revealed significant upregulation of PD-L1 expression across nearly all CD4^+^ and CD8^+^ T-cell subsets (figure 4). After two treatment cycles, PD-L1 expression was significantly increased in all memory (CD45RO^+^) T-cell subsets and across all CD4+ T-cell subsets, with the CD4^+^ memory T cells showing the highest expression and largest increases (figure 4A - left panel; figure 4B). Following four treatment cycles, PD-L1 expression was significantly elevated compared to baseline in all T-cell subsets except the effector memory (CD27^−^CD45RO^+^CD45RA^−^) T-cell subsets (figure 4A - right panel; figure 4B). Notably, the lowest PD-L1 expression among the T-cell subsets was observed in the naive (CD27^+^CD45RO^−^CD45RA^+^) T-cell subsets (figure 4B).

**Figure 4.**
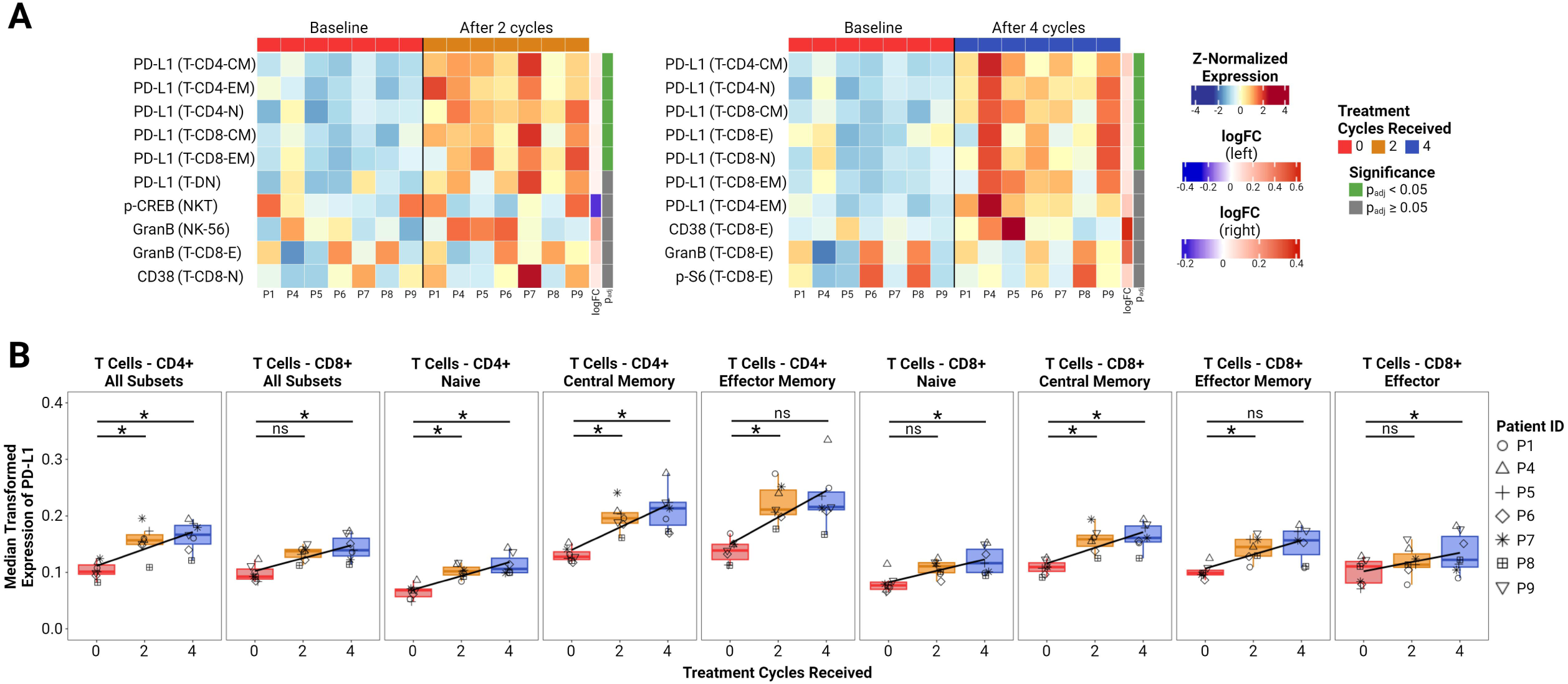
Significant changes in cell state marker expression during treatment. (A) Heatmaps of the top significant differences in the relative state marker expression in long-term survivor samples taken at baseline (n=7) (left half of the heatmaps) and after either two (right half of left heatmap) or four treatment cycles (right half of right heatmap). Each row of the heatmap is labeled with the combination of marker (left) and subset (right, in brackets) it represents, sorted from top to bottom based on increasing p-value. (B) Median arcSinh(5)-transformed expression of PD-L1 over time for all T-cell subsets in the long-term survivor (n=7) samples. The trendline was calculated using simple linear regression. * - p_adj_ < 0.05; CM - central memory; E - effector; EM - effector memory; GranB - granzyme B; logFC - log_2_(fold change); Mono - monocytes; N - naïve; NC - non-classical; NK-56 - CD56^+++^ NK cells; NKT - NKT cells; ns - not significant; T- CD4 - CD4^+^ T cells; T-CD8 - CD8^+^ T cells; T-DN - CD4^−^CD8^−^ (double-negative) T cells; Tgd - gamma-delta T cells

Comparisons of cell state marker expression between rapid and slow progressors, as well as between pre- and post-progression samples, did not reveal any significant differences. Interestingly, no significant changes or trends were observed in the expression levels of markers associated with the adenosinergic signaling pathway (CD39, CD73, p-CREB, and p-S6) (data not shown).

### Baseline expression levels of CD73 and IDO1 positively correlate with the duration of progression-free survival

To assess the impact of baseline abundances and phenotypes of peripheral blood leukocytes on PFS duration, we performed correlation analyses. No significant valid correlations were found between baseline subset abundances and PFS duration (figure 5A) (online supplemental table S3). However, analyses of cell state marker expression revealed a strong positive correlation between baseline CD73 expression and PFS duration for most of the subsets comprising the patients’ peripheral blood leukocyte repertoire (figure 5B). This correlation met the predefined robustness criteria in approximately one third of leukocyte subsets, predominantly encompassing the majority of the T-cell repertoire. Similarly, a strongly positive correlation with PFS duration was also observed for baseline IDO1 expression in a number of leukocyte subsets, although this correlation was significant in fewer subsets than observed for CD73 expression (figure 5C).

**Figure 5.**
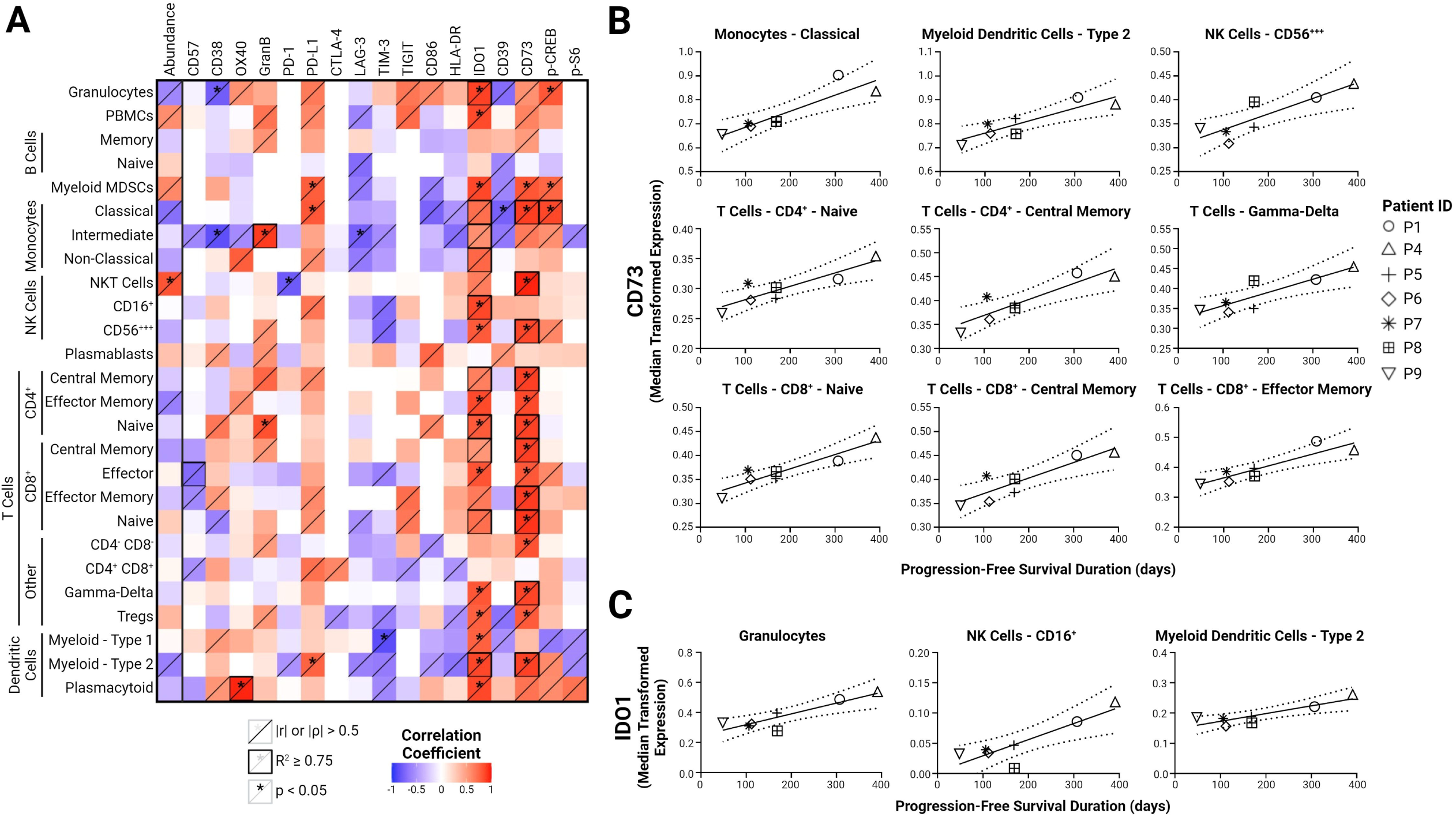
Correlations between progression-free survival duration and either the relative leukocyte subset abundances or state marker expression levels at baseline for long-term survivors (n=7). (A) Heatmap showing correlation analysis results (online supplemental table S3). Pearson or Spearman correlation analysis was selected based on the normality of the dataset, assessed using the Shapiro-Wilk test. (B) Plots depicting significant correlations between transformed median CD73 expression at baseline and time to progression. Symbols representing each patient are consistent throughout Fig. 5. (C) Plots illustrating most significant correlations between transformed median IDO1 expression at baseline and time to progression. |r| - absolute value of Pearson’s correlation coefficient; |ρ| - absolute value of Spearman’s correlation coefficient; GranB - granzyme B

## DISCUSSION

In this study, we report the findings from a high-dimensional single-cell analysis of blood samples from patients with relapsed EOC undergoing combination oleclumab/durvalumab immunotherapy. To the best of our knowledge, this is the first study to employ single-cell CyTOF to assess blood-based biomarkers in patients with ovarian cancer undergoing immunotherapy. Through differential and correlative analyses of the abundances and phenotypes of peripheral blood leukocytes, we identified phenotypic signatures potentially indicative of disease progression and treatment efficacy. We show that patients with relapsed EOC who present with lower proportions of PBMCs in their peripheral blood leukocyte pool may derive inferior survival benefit from combined oleclumab/durvalumab treatment than patients with higher proportions of PBMCs. During treatment, we saw an increase in the relative abundance of circulating CD14^+^CD16^−^ myeloid cells in the total patient cohort, along with a decrease in T_CM_ cell proportions specific to rapidly progressing patients. We also observed a strong trend towards an increased relative abundance of circulating M-MDSCs at progression. Furthermore, we noted a significant increase in PD-L1 expression on circulating T cells during treatment. Finally, we demonstrate that PFS duration was positively correlated with pre-treatment expression levels of CD73 and IDO1 on several circulating leukocyte subsets.

In our study cohort, long-term survivors exhibited significantly higher proportions of total PBMCs at baseline compared to short-term survivors, which aligns with studies on PD-1/PD-L1 checkpoint blockade in lung cancer and melanoma demonstrating significant association between lower baseline neutrophil-to-lymphocyte ratios and improved progression-free and overall survival [31,32]. Correspondingly, patient P4, who exhibited the highest relative abundance of lymphocytes at baseline among all patients (figure 2D), was the sole responder in the trial and demonstrated the longest survival. This data suggests that EOC patients possessing an immune system more effectively primed for adaptive response may derive more consistent benefit from immunotherapy.

We observed a significant expansion of the CD14^+^CD16^−^ myeloid cell compartment over the course of the first four treatment cycles. This expansion was initially driven by an increased abundance of HLA-DR^+/hi^ classical monocytes in the total cohort, and further sustained by an elevated frequency of HLA-DR^−/lo^ M-MDSCs in rapid progressors. This indicates a shift of the CD14^+^CD16^−^ myeloid compartment towards a more immunosuppressive phenotype. Therefore, a sudden increase in the proportions of M-MDSCs in the peripheral blood of EOC patients treated with anti-CD73/PD-L1 immunotherapy may serve as a potential biomarker of disease progression. Accordingly, higher HLA-DR expression on cells within the CD14^+^CD16^−^ myeloid cell compartment has been positively associated with improved response to oleclumab [21] and immunotherapy targeting the PD-1/PD-L1 pathway [33]. Furthermore, cancer immunotherapy trials have consistently demonstrated that elevated MDSC abundance is associated with worse outcome in EOC [34], and various other cancers [35–37]. Growth factors and proinflammatory cytokines produced by cancer cells chronically stimulate myeloid progenitors, driving their differentiation towards M-MDSCs. Similarly, tumor-derived factors can induce the reprogramming of classical monocytes already present in peripheral blood towards M-MDSCs [38]. This expanded population of CD14^+^CD16^−^ myeloid cells, likely representing precursors to tumor-associated macrophages, can be recruited to ovarian tumors, resulting in enhanced immunosuppression [34]. We hypothesize that EOC’s concurrent utilization of a multitude of immunosuppressive mechanisms allows it to adapt and effectively resist targeted immunotherapy through enhancement of immunosuppression mechanisms not currently affected by treatment. Such adaptation may be addressed by modifications or additions to the existing therapeutic approach. In line with this notion, MDSC-targeting agents have been preclinically and clinically evaluated in EOC with promising beneficial effect, also in combination with other immunotherapeutic approaches [34,39].

Our analysis showed a significant decline in the frequencies of T_CM_ cells in the peripheral blood of rapid progressors. Such a decline may have occurred as a consequence of the evolving immunosuppression in the underlying cancer. This hypothesis is supported by the observation that the T_CM_-cell decline coincides with a substantial increase in peripheral M-MDSCs in these patients. T_CM_-cell homeostasis may have been disrupted by the collective inhibition of the activation and differentiation of effector T cells into T_CM_ cells, and the inhibition of the reactivation and proliferation of tumor-proximal T_CM_ cells through an adaptive increase in adenosine generation. This effect may have been mediated by upregulation or induction of CD39/CD73 expression on tumor-recruited MDSCs [40], creating what is known as a “purinergic halo” [41]. The role of adenosine in driving the contraction of the T_CM_ subset is further supported the study of Mastelic-Gavillet et al., which showed that T_CM_ cells are especially vulnerable to adenosine-mediated dysfunction due to their high expression of the A2A receptor [30].

A novel observation of this study was the significant increase of PD-L1 on circulating T cells over time during administration of an anti-PD-L1 ICI. Research on other solid cancers has identified intratumoral PD-L1^+^CD8^+^ T cells as drivers of T-cell exhaustion via the PD-L1/PD-1 axis, and as promoters of M2 macrophage polarization [42,43]. Given that PD-L1 upregulation on T cells occurs following activation, we postulate that the observed longitudinal upregulation of PD-L1 on circulating T cells in patients with EOC receiving oleclumab/durvalumab is a consequence of rapidly induced and progressively mutual exhaustion of activated T-cells.

Interestingly, we did not observe any significant longitudinal changes or inter-group differences in the expression of markers associated with adenosinergic signaling (CD39, CD73, p-S6, and p-CREB). This contradicts previous findings that documented oleclumab-induced inhibition and internalization of CD73 on CD4^+^ and CD8^+^ T cells [44]. The stability of adenosinergic signaling observed in our study may be attributable to compensatory upregulation of CD73 expression, which possibly counteracted the expected oleclumab-mediated internalization.

Our study demonstrates a significant positive correlation between baseline CD73 expression across various leukocyte subsets and PFS duration. This contradicts the prevailing view that CD73 expression in cancer generally correlates with poorer treatment outcome [45]. A limited number of studies have investigated CD73 expression in EOC [18,46], and ours is the first to link baseline CD73 expression with outcome in CD73-targeting cancer therapy. Although the positive correlation between baseline CD73 expression and PFS duration was observed for a range of leukocyte subsets, nearly all T-cell subsets are present in this set and represent a majority of the significant results. Thus, elevated baseline CD73 expression on peripheral blood T cells may serve as a predictive marker of longer PFS in patients with EOC set to undergo anti-CD73 immunotherapy. We hypothesize that patients with higher baseline CD73 expression may have had tumors more heavily dependent on adenosinergic signaling for immune evasion, potentially enhancing the effectiveness of anti-CD73 treatment. Nonetheless, this advantage appears to be temporary, as all patients ultimately experienced progression. This development may be attributed to the inherent immunosuppressive adaptability of EOC, as evident by the observed expansion of M-MDSCs and upregulation of PD-L1 in T cells.

In addition to the observed positive correlation between CD73 and longer PFS, our observation of a positive correlation between baseline IDO1 expression and longer PFS during EOC immunotherapy is in alignment with previous reports [47,48]. Although IDO1 is generally viewed as a contributor to immunosuppression, our findings may be explained by its role in upregulating PD-L1 through the activation of the aryl hydrocarbon receptor by IDO1-generated tryptophan catabolites, as observed in solid tumors [49] and murine models of EOC [50]. Consequently, higher IDO1 expression in specific cell subsets could lead to increased PD-L1 expression on the same cells, potentially enhancing their susceptibility to durvalumab. Indeed, consistent with the study by Fujiwara et al. [47], our data also demonstrated a positive correlation between IDO1 and PD-L1 expression in leukocytes (online supplemental table S4). This correlation was significant and robust in subsets that showed the strongest associations between PD-L1 expression and PFS duration, namely, classical monocytes, M-MDSCs, and CD141^+^ myeloid dendritic cells. Therefore, IDO1 expression in the peripheral blood myeloid compartment may serve as a potential biomarker for the efficacy of EOC immunotherapy targeting PD-L1. Despite strong correlation coefficients and significant p-values, the validity of correlations between PD-L1 and PFS duration was limited by a low goodness-of-fit parameter, likely due to the small sample size. Therefore, a larger study is needed to more definitively establish baseline PD-L1 expression in peripheral blood leukocytes as a predictive biomarker for EOC immunotherapy utilizing anti-PD-L1 ICIs.

This study examined sequentially sampled peripheral blood. An advantage of using blood samples, as opposed to tumor tissue biopsies, is that the information provided is not limited to a specific anatomical location. This is particularly valuable in cases of metastatic disease and offers a more comprehensive view of the patient’s overall condition. Additionally, blood sampling is more flexible, clinically accessible, cost-effective, less invasive, and avoids the need for radiological imaging or surgical procedures for sample acquisition. However, the implementation of our 40-marker panel in the clinical setting is constrained by the high costs and complexity of the CyTOF methodology. Therefore, a more focused set of markers or leukocyte subsets, as suggested in this study, could represent a biomarker panel analyzable via methods already utilized in routine clinical diagnostics, such as flow cytometry or ELISA.

Overall, the small number of patient samples in this study affects the statistical power and limits the generalizability of the results. However, the diversity within the patient population - encompassing a wide range of ages, disease stages, and prior treatments - adds to the robustness and relevance of our observations for a broader spectrum of patients with relapsed EOC undergoing immunotherapy. Furthermore, the use of single-cell suspension CyTOF allowed us to generate extensive data from the limited sample set. Coupled with high-dimensional unsupervised clustering algorithms, this approach provided numerous insights and perspectives, strengthening the conclusions of the study.

Another limitation of this study is the absence of control arms comprising patients treated with either oleclumab or durvalumab alone [15]. Such control groups would have provided data integral for the conclusive attribution of the observed differences in leukocyte abundances and states to the effects of a certain immunotherapeutic. In addition, patients had undergone multiple rounds of cancer therapy prior to inclusion, which may have exerted developmental pressure on the tumors and potentially influenced response to oleclumab/durvalumab treatment. Although an expanded trial including single-agent arms and newly diagnosed EOC patients would help clarify the individual contribution of each compound to treatment outcome, the limited clinical activity of the oleclumab/durvalumab combination in the NSGO-OV-UMB1 trial [15] makes it difficult and ethically questionable to initiate such a randomized phase II or III trial.

In conclusion, our findings offer comprehensive insights into the effects of combination oleclumab/durvalumab immunotherapy on the peripheral blood leukocytes of relapsed EOC patients and elucidate potential immunosuppressive mechanisms employed by EOC to counteract the effects of immunotherapy. We propose blood-based biomarkers for better patient selection and non-invasive monitoring of disease progression; however, validation in larger patient cohorts is necessary.

## DECLARATIONS

## Acknowledgements

We thank the patients for their consent and participation in the study. We thank Brith Bergum and Jørn Skavland at the Flow Cytometry Core Facility of the University of Bergen for providing support for our mass cytometry work. We acknowledge BioRender.com as the platform used to create the figures in this manuscript.

## Authors’ Contributions

Study conceptualization and design: LT, MRM, LB. Clinical sample collection: AA, JM, KM, KSP, MRM, LB. Clinical database administration: RdPC, KM, KSP. Data generation: LT, CEW. Data analysis and statistics: LT. Data interpretation: LT, KK, LCVT, EMC, LB. Manuscript writing and editing: LT, KK, LCVT, LB. Manuscript review: All authors. Project administration: LT, KM, KSP, MRM, LB. Funding acquisition: MRM, LB. Guarantors: MRM, LB.

## Funding

This work was supported by grants from the Western Norway Regional Health Authority (Project No. 28543) and the Research Council of Norway through its Centres of Excellence funding scheme (Project No. 223250). AstraZeneca provided a grant to partially fund the trial and the investigational medicinal products (oleclumab and durvalumab). The funding sources were not involved in the study design, the collection of data, or the analysis and interpretation of data.

## Competing interests

AA has participated on the advisory boards of GlaxoSmithKline and Merck Sharp and Dohme; RdPC is employed by and is a shareholder of Y-mAbs Pharmaceuticals; LCVT has received personal fees for lectures from Bayer and AstraZeneca, personal fee payments from Eisai for participating on an expert board, and has received a grant related to a clinical trial from AstraZeneca; CEW has received financial support for conference attendance and travel expenses from Beckman Coulter Inc.; EMC is a shareholder of KinN Therapeutics AS; JM has received a honorarium for a lecture from Eisai; KM has received speakers’ honoraria and received compensation for travel expenses from GlaxoSmithKline and AstraZeneca, has participated in a trial-specific safety review committee for Kancera AB, and is a deputy medical director for NSGO-CTU; MRM has received an institutional study grant and investigational medicinal product from AstraZeneca (no personal grants were received); LB has received honoraria for lectures from GlaxoSmithKline and Merck Sharp and Dohme, has received a research grant from AstraZeneca for a researcher-initiated trial, and has had leadership roles in Onkologisk Forum between 2018 and 2022 and in the NSGO and NSGO-CTU since 2021; LT, KK and KSP report no personal conflicts of interest.

## Data availability statement

Data are available from the corresponding author upon request.

## Ethics approval

The study protocol and use of patient material in this study was approved by the regional ethical committees or institutional review boards of the participating sites: The Scientific Ethics Committee for the Capital Region of Denmark (VEK) (approval no. H-17025483), The Regional Committee for Medical Research Ethics Western Norway (REK West) (approval no. 2018/580), and The Regional Ethics Committee of the Expert Responsibility area of Tampere University Hospital (TAYS) (approval no. R18078M). The study was conducted in accordance with the good clinical practice guidelines and provisions of the Declaration of Helsinki, as well as according to all local regulations.

## Patient consent

All patients provided written informed consent for use of the clinical information and biological materials.

## Provenance and peer review

Not commissioned; externally peer reviewed.

## Supplemental material

This content has been supplied by the author(s). It has not been vetted by BMJ Publishing Group Limited (BMJ) and may not have been peer-reviewed. Any opinions or recommendations discussed are solely those of the author(s) and are not endorsed by BMJ. BMJ disclaims all liability and responsibility arising from any reliance placed on the content. Where the content includes any translated material, BMJ does not warrant the accuracy and reliability of the translations (including but not limited to local regulations, clinical guidelines, terminology, drug names and drug dosages), and is not responsible for any error and/or omissions arising from translation and adaptation or otherwise.

## Open access

This is an open access article distributed in accordance with the Creative Commons Attribution (CC BY 4.0) license, which permits others to copy, redistribute, remix, transform and build upon this work for any purpose, provided the original work is properly cited, appropriate credit is given, any changes made are indicated. See: https://creativecommons.org/licenses/by/4.0/.

## Notes

### Funding Statement

This study was funded by grants from the Western Norway Regional Health Authority (Project No. 28543) and the Research Council of Norway through its Centres of Excellence funding scheme (Project No. 223250). AstraZeneca provided a grant to partially fund the trial and the investigational medicinal products (oleclumab and durvalumab). The funding sources were not involved in the study design, the collection of data, or the analysis and interpretation of data.

### Author Declarations

Ethics committee/IRB of the Capital Region of Denmark (VEK) gave ethical approval for this work (approval no. H-17025483). Ethics committee/IRB for Medical Research Ethics Western Norway (REK West) gave ethical approval for this work (approval no. 2018/580). Ethics committee/IRB of the Expert Responsibility area of Tampere University Hospital (TAYS) gave ethical approval for this work (approval no. R18078M).

