## Supplementary figures and images for "Peripheral blood leukocyte signatures as biomarkers in relapsed ovarian cancer patients receiving combined anti-CD73/anti-PD-L1 immunotherapy in Arm A of the NSGO-OV-UMB1/ENGOT-OV30 trial"

### Supplemental Figure S1

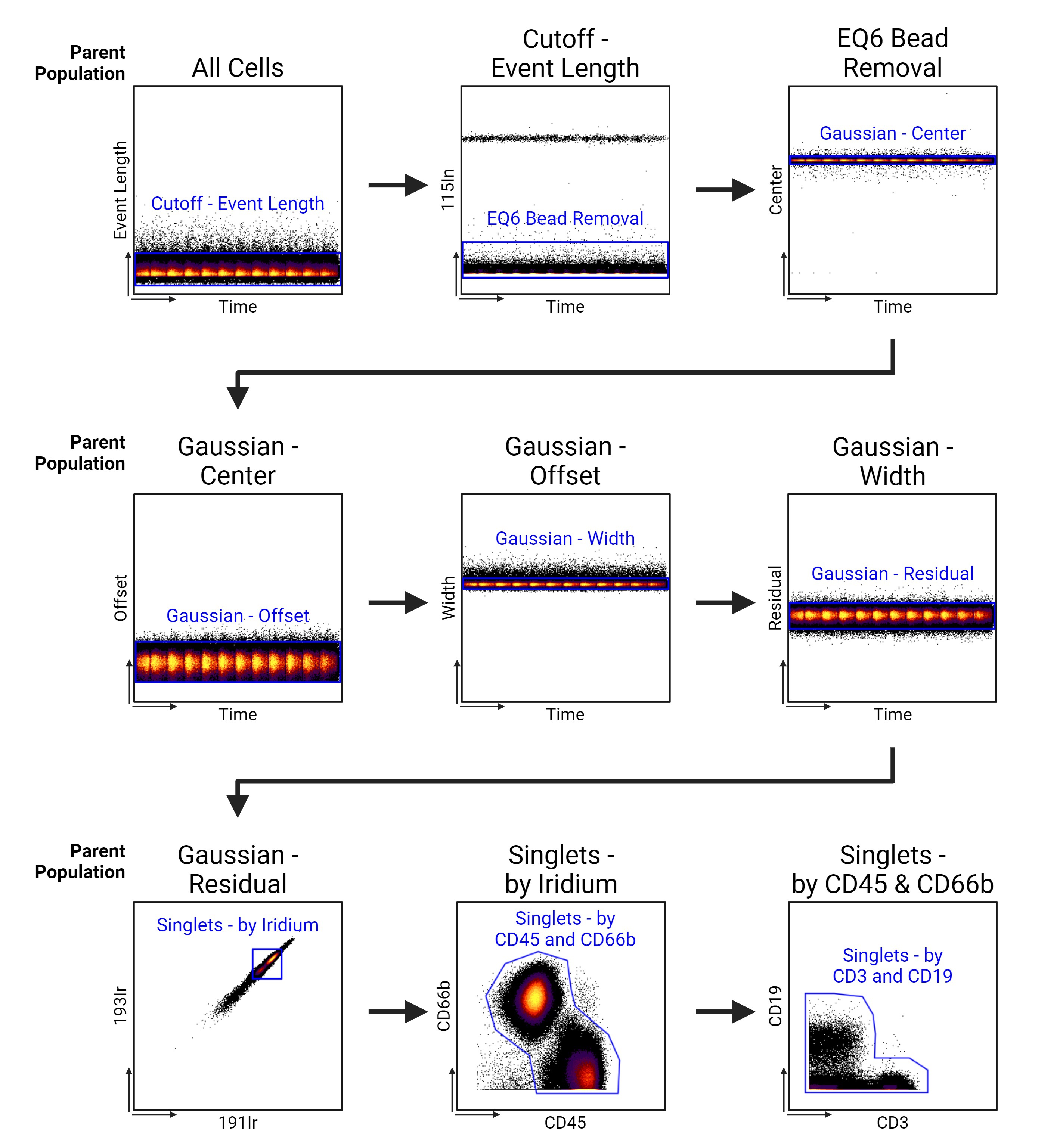

### Supplemental Figure S2

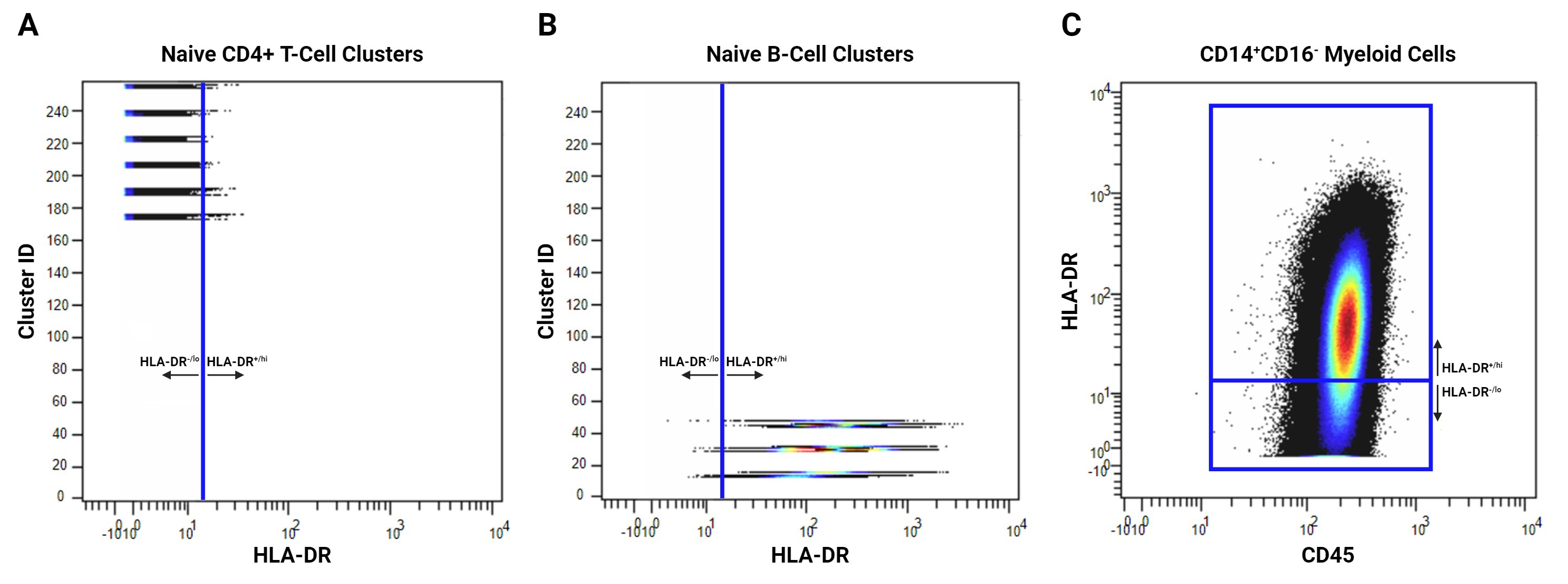
