## Supplemental Table S1 for "Peripheral blood leukocyte signatures as biomarkers in relapsed ovarian cancer patients receiving combined anti-CD73/anti-PD-L1 immunotherapy in Arm A of the NSGO-OV-UMB1/ENGOT-OV30 trial"

**Online supplemental table S1** The 40-marker mass cytometry panel used for immunophenotyping in this study. *Titrated on both non-stimulated leukocytes and stimulated PBMCs; †Titrated on stimulated PBMCs; ‡Intracellular target; §Conjugated in-house

| Metal | Target | Clone | Dilution | Host | Isotype | Vendor | Cat. No. |
|---|---|---|---|---|---|---|---|
| 89Y | CD45 | HI30 | 1:200 | Mouse | IgG1, κ | Standard BioTools | 3089003B |
| 111Cd | CD4 | RPA-T4§ | 2 µg / mL | Mouse | IgG1, κ | BioLegend | 300541 |
| 113Cd | CD8 | HIT8a§ | 0,25 µg / mL | Mouse | IgG1, κ | BioLegend | 300902 |
| 116Cd | CD57* | HNK-1§ | 0,25 µg / mL | Mouse | IgM, κ | BioLegend | 359602 |
| 141Pr | CD3 | UCHT1 | 1:400 | Mouse | IgG1, κ | Standard BioTools | 3141019B |
| 142Nd | OX40† | ACT35 | 1:100 | Mouse | IgG1, κ | Standard BioTools | 3142018B |
| 143Nd | CD19 | HIB19§ | 0,5 µg / mL | Mouse | IgG1, κ | BioLegend | 302202 |
| 144Nd | CD38 | HIT2 | 1:400 | Mouse | IgG1, κ | Standard BioTools | 3144014B |
| 145Nd | CD20 | 2H7§ | 1 µg / mL | Mouse | IgG2b, κ | BioLegend | 302302 |
| 146Nd | CD11c | Bu15§ | 1 µg / mL | Mouse | IgG1, κ | BioLegend | 337202 |
| 147Sm | CD45RA | HI100§ | 0,5 µg / mL | Mouse | IgG2b, κ | BioLegend | 304143 |
| 148Nd | CD14 | RMO52 | 1:1600 | Mouse | IgG2a, κ | Standard BioTools | 3148010B |
| 149Sm | CD66b | G10F5§ | 0,5 µg / mL | Mouse | IgM, κ | Novus Biologicals | NB100-77808 |
| 150Nd | LAG-3† | 11C3C65 | 1:100 | Mouse | IgG1, κ | Standard BioTools | 3150030B |
| 151Eu | CD123 | 6H6 | 1:200 | Mouse | IgG1, κ | Standard BioTools | 3151001B |
| 152Sm | TCRgd | 11F2 | 1:100 | Mouse | IgG1, κ | Standard BioTools | 3152008B |
| 153Eu | TIGIT† | MBSA43 | 1:100 | Mouse | IgG1, κ | Standard BioTools | 3153019B |
| 154Sm | TIM-3† | F38-2E2 | 1:100 | Mouse | IgG1, κ | Standard BioTools | 3154010B |
| 155Gd | CD27 | L128 | 1:200 | Mouse | IgG1, κ | Standard BioTools | 3155001B |
| 156Gd | CD86† | IT2.2 | 1:200 | Mouse | IgG2b, κ | Standard BioTools | 3156008B |
| 158Gd | CD33 | WM53 | 1:100 | Mouse | IgG1, κ | Standard BioTools | 3158001B |
| 159Tb | PD-L1† | 29E.2A3 | 1:150 | Mouse | IgG2b, κ | Standard BioTools | 3159029B |
| 160Gd | CD39† | A1 | 1:100 | Mouse | IgG1, κ | Standard BioTools | 3160004B |
| 161Dy | IDO1*‡ | GT273§ | 3 µg / mL | Mouse | IgG2a, κ | GeneTex | GTX634652 |
| 162Dy | FoxP3*‡ | 236A/E7§ | 8 µg / mL | Mouse | IgG1, κ | ThermoFisher | 14-4777-82 |
| 163Dy | CD56 | NCAM16.2 | 1:4000 | Mouse | IgG2b, κ | Standard BioTools | 3163007B |
| 164Dy | CD45RO | UCHL1 | 1:800 | Mouse | IgG2a, κ | Standard BioTools | 3164007B |
| 165Ho | p-CREB [S133]†‡ | 87G3 | 1:200 | Rabbit | IgG | Standard BioTools | 3165009A |
| 166Er | CD34 | 581 | 1:1600 | Mouse | IgG1, κ | Standard BioTools | 3166012B |
| 167Er | CD73* | 7G2§ | 8 µg / mL | Mouse | IgG2a, κ | ThermoFisher | 41-0200 |
| 168Er | CD16 | B73.1§ | 2 µg / mL | Mouse | IgG1, κ | BioLegend | 360702 |
| 169Tm | CD25* | 2A3 | 1:2000 | Mouse | IgG1, κ | Standard BioTools | 3169003B |
| 170Er | CTLA-4† | 14D3 | 1:100 | Mouse | IgG2a, κ | Standard BioTools | 3170005B |
| 171Yb | Granzyme B*‡ | GB11 | 1:100 | Mouse | IgG1, κ | Standard BioTools | 3171002B |
| 172Yb | p-S6 [pS235/pS236]†‡ | N7-548 | 1:200 | Mouse | IgG1, κ | Standard BioTools | 3172008A |
| 173Yb | CD141 | 1A4 | 1:1600 | Mouse | IgG1, κ | Standard BioTools | 3173002B |
| 174Yb | HLA-DR | L243 | 1:100 | Mouse | IgG2a, κ | Standard BioTools | 3174001B |
| 175Lu | PD-1† | EH12.2H7 | 1:400 | Mouse | IgG1, κ | Standard BioTools | 3175008B |
| 176Yb | CD1c | L161§ | 0,5 µg / mL | Mouse | IgG1, κ | BioLegend | 331502 |
| 191Ir | DNA | N/A | 62,5 nM | N/A | N/A | Standard BioTools | 201192B |
| 193Ir | DNA | N/A | 62,5 nM | N/A | N/A | Standard BioTools | 201192B |
| 209Bi | CD11b | ICRF44 | 1:100 | Mouse | IgG1, κ | Standard BioTools | 3209003B |
