## Supplemental Table S2 for "Peripheral blood leukocyte signatures as biomarkers in relapsed ovarian cancer patients receiving combined anti-CD73/anti-PD-L1 immunotherapy in Arm A of the NSGO-OV-UMB1/ENGOT-OV30 trial"

### **Online supplemental table S2** Detailed sample composition data.

| Data | Subset | C0_P1 | C2_P1 | C3_P1 | 4_P1 | C7_P1 | C9_P1 | C10_P1 | C0_P2 | C0_P3 | C0_P4 | C1_P4 | C2_P4 |
|---|---|---|---|---|---|---|---|---|---|---|---|---|---|
| Cell Count | Granulocytes | 446665 | 464764 | 471224 | 413459 | 507321 | 444116 | 389894 | 506580 | 483765 | 355809 | 351765 | 379578 |
| Cell Count | PBMCs | 146396 | 103720 | 107213 | 86797 | 84043 | 112168 | 118156 | 94267 | 51374 | 183766 | 229348 | 220960 |
| % of Total Leukocytes | Granulocytes | 75,32 % | 81,75 % | 81,47 % | 82,65 % | 85,79 % | 79,84 % | 76,74 % | 84,31 % | 90,40 % | 65,94 % | 60,53 % | 63,21 % |
| % of Total Leukocytes | PBMCs | 24,68 % | 18,25 % | 18,53 % | 17,35 % | 14,21 % | 20,16 % | 23,26 % | 15,69 % | 9,60 % | 34,06 % | 39,47 % | 36,79 % |
| Cell Count | B Cells - Memory | 896 | 610 | 697 | 543 | 408 | 694 | 750 | 104 | 319 | 1023 | 1301 | 979 |
| Cell Count | B Cells - Naive | 9502 | 6963 | 7251 | 5230 | 5093 | 6458 | 6992 | 3043 | 3783 | 8650 | 12086 | 10009 |
| Cell Count | Monocytic MDSCs | 15298 | 6404 | 12873 | 9624 | 14007 | 9033 | 14382 | 3014 | 11436 | 8076 | 2951 | 6299 |
| Cell Count | Monocytes - Classical | 32098 | 31880 | 27397 | 24041 | 13618 | 28586 | 27207 | 22264 | 11316 | 20303 | 48097 | 50141 |
| Cell Count | Monocytes - Intermediate | 2818 | 1950 | 1744 | 1733 | 1754 | 1807 | 2405 | 2198 | 918 | 1758 | 4552 | 3517 |
| Cell Count | Monocytes - Non-Classical | 4940 | 2852 | 2063 | 2557 | 2908 | 2949 | 3988 | 2059 | 567 | 1876 | 3541 | 2750 |
| Cell Count | NKT Cells | 2120 | 1537 | 1909 | 1502 | 1450 | 1870 | 1475 | 2621 | 5733 | 22702 | 15834 | 15031 |
| Cell Count | NK Cells - CD16+ | 25313 | 10265 | 8090 | 7532 | 10461 | 9736 | 16413 | 21344 | 4175 | 6896 | 5397 | 4292 |
| Cell Count | NK Cells - CD56+++ | 1438 | 987 | 1069 | 843 | 853 | 1118 | 1021 | 715 | 198 | 671 | 854 | 766 |
| Cell Count | Plasmablasts | 79 | 86 | 177 | 93 | 76 | 117 | 99 | 41 | 134 | 79 | 43 | 26 |
| Cell Count | T Cells - CD4+ - Central Memory | 24013 | 19171 | 20196 | 14903 | 13851 | 22335 | 19375 | 12880 | 2525 | 19806 | 23294 | 24988 |
| Cell Count | T Cells - CD4+ - Effector Memory | 1085 | 808 | 1073 | 627 | 630 | 894 | 871 | 5404 | 1068 | 2620 | 2578 | 2439 |
| Cell Count | T Cells - CD4+ - Naive | 11090 | 9319 | 10334 | 7977 | 10229 | 13191 | 10437 | 685 | 3377 | 22227 | 28205 | 30489 |
| Cell Count | T Cells - CD8+ - Central Memory | 7951 | 5454 | 6289 | 4814 | 3971 | 6482 | 6326 | 3667 | 1128 | 8412 | 10023 | 8883 |
| Cell Count | T Cells - CD8+ - Effector | 340 | 171 | 236 | 227 | 222 | 294 | 378 | 4998 | 1773 | 29547 | 35066 | 28115 |
| Cell Count | T Cells - CD8+ - Effector Memory | 687 | 404 | 532 | 399 | 369 | 495 | 508 | 3526 | 832 | 1970 | 1732 | 1531 |
| Cell Count | T Cells - CD8+ - Naive | 3026 | 2061 | 2487 | 1933 | 1985 | 2931 | 2791 | 2048 | 802 | 19506 | 24831 | 22357 |
| Cell Count | T Cells - CD4- CD8- | 179 | 188 | 201 | 137 | 202 | 237 | 204 | 215 | 81 | 727 | 970 | 947 |
| Cell Count | T Cells - CD4+ CD8+ | 28 | 30 | 18 | 11 | 8 | 26 | 13 | 26 | 2 | 67 | 151 | 141 |
| Cell Count | T Cells - Gamma-Delta | 588 | 369 | 522 | 374 | 308 | 447 | 423 | 1528 | 708 | 3533 | 3551 | 2972 |
| Cell Count | Tregs | 1769 | 1379 | 1476 | 1181 | 1138 | 1647 | 1434 | 743 | 248 | 2077 | 2351 | 2763 |
| Cell Count | Myeloid Dendritic Cells - Type 1 | 38 | 34 | 30 | 26 | 31 | 36 | 46 | 49 | 19 | 33 | 52 | 52 |
| Cell Count | Myeloid Dendritic Cells - Type 2 | 583 | 315 | 269 | 228 | 164 | 370 | 308 | 576 | 107 | 595 | 1148 | 885 |
| Cell Count | Plasmacytoid Dendritic Cells | 339 | 265 | 149 | 169 | 183 | 201 | 189 | 240 | 61 | 387 | 327 | 294 |
| % of PBMCs | B Cells - Memory | 0,12 % | 0,09 % | 0,10 % | 0,09 % | 0,06 % | 0,10 % | 0,12 % | 0,02 % | 0,05 % | 0,14 % | 0,16 % | 0,12 % |
| % of PBMCs | B Cells - Naive | 1,29 % | 1,04 % | 1,06 % | 0,89 % | 0,76 % | 0,97 % | 1,12 % | 0,44 % | 0,65 % | 1,21 % | 1,51 % | 1,23 % |
| % of PBMCs | Monocytic MDSCs | 2,08 % | 0,96 % | 1,88 % | 1,65 % | 2,08 % | 1,36 % | 2,31 % | 0,44 % | 1,95 % | 1,13 % | 0,37 % | 0,77 % |
| % of PBMCs | Monocytes - Classical | 4,36 % | 4,76 % | 4,01 % | 4,11 % | 2,02 % | 4,30 % | 4,36 % | 3,22 % | 1,93 % | 2,83 % | 5,99 % | 6,16 % |
| % of PBMCs | Monocytes - Intermediate | 0,38 % | 0,29 % | 0,26 % | 0,30 % | 0,26 % | 0,27 % | 0,39 % | 0,32 % | 0,16 % | 0,25 % | 0,57 % | 0,43 % |
| % of PBMCs | Monocytes - Non-Classical | 0,67 % | 0,43 % | 0,30 % | 0,44 % | 0,43 % | 0,44 % | 0,64 % | 0,30 % | 0,10 % | 0,26 % | 0,44 % | 0,34 % |
| % of PBMCs | NKT Cells | 0,29 % | 0,23 % | 0,28 % | 0,26 % | 0,22 % | 0,28 % | 0,24 % | 0,38 % | 0,98 % | 3,17 % | 1,97 % | 1,85 % |
| % of PBMCs | NK Cells - CD16+ | 3,44 % | 1,53 % | 1,18 % | 1,29 % | 1,55 % | 1,46 % | 2,63 % | 3,09 % | 0,71 % | 0,96 % | 0,67 % | 0,53 % |
| % of PBMCs | NK Cells - CD56+++ | 0,20 % | 0,15 % | 0,16 % | 0,14 % | 0,13 % | 0,17 % | 0,16 % | 0,10 % | 0,03 % | 0,09 % | 0,11 % | 0,09 % |
| % of PBMCs | Plasmablasts | 0,01 % | 0,01 % | 0,03 % | 0,02 % | 0,01 % | 0,02 % | 0,02 % | 0,01 % | 0,02 % | 0,01 % | 0,01 % | 0,00 % |
| % of PBMCs | T Cells - CD4+ - Central Memory | 3,26 % | 2,86 % | 2,96 % | 2,55 % | 2,06 % | 3,36 % | 3,11 % | 1,86 % | 0,43 % | 2,76 % | 2,90 % | 3,07 % |
| % of PBMCs | T Cells - CD4+ - Effector Memory | 0,15 % | 0,12 % | 0,16 % | 0,11 % | 0,09 % | 0,13 % | 0,14 % | 0,78 % | 0,18 % | 0,37 % | 0,32 % | 0,30 % |
| % of PBMCs | T Cells - CD4+ - Naive | 1,51 % | 1,39 % | 1,51 % | 1,36 % | 1,52 % | 1,98 % | 1,67 % | 0,10 % | 0,58 % | 3,10 % | 3,51 % | 3,75 % |
| % of PBMCs | T Cells - CD8+ - Central Memory | 1,08 % | 0,81 % | 0,92 % | 0,82 % | 0,59 % | 0,97 % | 1,01 % | 0,53 % | 0,19 % | 1,17 % | 1,25 % | 1,09 % |
| % of PBMCs | T Cells - CD8+ - Effector | 0,05 % | 0,03 % | 0,03 % | 0,04 % | 0,03 % | 0,04 % | 0,06 % | 0,72 % | 0,30 % | 4,12 % | 4,37 % | 3,45 % |
| % of PBMCs | T Cells - CD8+ - Effector Memory | 0,09 % | 0,06 % | 0,08 % | 0,07 % | 0,05 % | 0,07 % | 0,08 % | 0,51 % | 0,14 % | 0,27 % | 0,22 % | 0,19 % |
| % of PBMCs | T Cells - CD8+ - Naive | 0,41 % | 0,31 % | 0,36 % | 0,33 % | 0,29 % | 0,44 % | 0,45 % | 0,30 % | 0,14 % | 2,72 % | 3,09 % | 2,75 % |
| % of PBMCs | T Cells - CD4- CD8- | 0,02 % | 0,03 % | 0,03 % | 0,02 % | 0,03 % | 0,04 % | 0,03 % | 0,03 % | 0,01 % | 0,10 % | 0,12 % | 0,12 % |
| % of PBMCs | T Cells - CD4+ CD8+ | 0,00 % | 0,00 % | 0,00 % | 0,00 % | 0,00 % | 0,00 % | 0,00 % | 0,00 % | 0,00 % | 0,01 % | 0,02 % | 0,02 % |
| % of PBMCs | T Cells - Gamma-Delta | 0,08 % | 0,06 % | 0,08 % | 0,06 % | 0,05 % | 0,07 % | 0,07 % | 0,22 % | 0,12 % | 0,49 % | 0,44 % | 0,37 % |
| % of PBMCs | Tregs | 0,24 % | 0,21 % | 0,22 % | 0,20 % | 0,17 % | 0,25 % | 0,23 % | 0,11 % | 0,04 % | 0,29 % | 0,29 % | 0,34 % |
| % of PBMCs | Myeloid Dendritic Cells - Type 1 | 0,01 % | 0,01 % | 0,00 % | 0,00 % | 0,00 % | 0,01 % | 0,01 % | 0,01 % | 0,00 % | 0,00 % | 0,01 % | 0,01 % |
| % of PBMCs | Myeloid Dendritic Cells - Type 2 | 0,08 % | 0,05 % | 0,04 % | 0,04 % | 0,02 % | 0,06 % | 0,05 % | 0,08 % | 0,02 % | 0,08 % | 0,14 % | 0,11 % |
| % of PBMCs | Plasmacytoid Dendritic Cells | 0,05 % | 0,04 % | 0,02 % | 0,03 % | 0,03 % | 0,03 % | 0,03 % | 0,03 % | 0,01 % | 0,05 % | 0,04 % | 0,04 % |

| Data | Subset | C4_P4 | C6_P4 | C8_P4 | C10_P4 | C12_P4 | C14_P4 | C0_P5 | C1_P5 | C2_P5 | C4_P5 | C6_P5 | C0_P6 |
|---|---|---|---|---|---|---|---|---|---|---|---|---|---|
| Cell Count | Granulocytes | 373817 | 407736 | 492530 | 338282 | 394526 | 430703 | 293239 | 282802 | 298716 | 291809 | 344423 | 280455 |
| Cell Count | PBMCs | 186438 | 314279 | 153107 | 251649 | 261792 | 198529 | 101962 | 83290 | 79692 | 93619 | 84243 | 90490 |
| % of Total Leukocytes | Granulocytes | 66,72 % | 56,47 % | 76,29 % | 57,34 % | 60,11 % | 68,45 % | 74,20 % | 77,25 % | 78,94 % | 75,71 % | 80,35 % | 75,61 % |
| % of Total Leukocytes | PBMCs | 33,28 % | 43,53 % | 23,71 % | 42,66 % | 39,89 % | 31,55 % | 25,80 % | 22,75 % | 21,06 % | 24,29 % | 19,65 % | 24,39 % |
| Cell Count | B Cells - Memory | 935 | 1960 | 997 | 1472 | 1834 | 1243 | 607 | 280 | 410 | 402 | 485 | 357 |
| Cell Count | B Cells - Naive | 12936 | 22910 | 10193 | 18702 | 21448 | 13278 | 1837 | 1388 | 2264 | 2422 | 3055 | 6984 |
| Cell Count | Monocytic MDSCs | 3290 | 5912 | 1539 | 1834 | 3015 | 3771 | 668 | 31 | 1270 | 389 | 1905 | 3258 |
| Cell Count | Monocytes - Classical | 30634 | 45368 | 25562 | 42282 | 44433 | 36873 | 26498 | 32184 | 29756 | 35849 | 28074 | 21035 |
| Cell Count | Monocytes - Intermediate | 2404 | 4360 | 2201 | 3472 | 2604 | 4389 | 2181 | 6098 | 2166 | 4682 | 3549 | 548 |
| Cell Count | Monocytes - Non-Classical | 3717 | 4270 | 1925 | 3221 | 2724 | 2903 | 2549 | 2164 | 2063 | 4049 | 3981 | 1637 |
| Cell Count | NKT Cells | 17683 | 30580 | 13848 | 29803 | 26655 | 21316 | 338 | 131 | 299 | 343 | 436 | 644 |
| Cell Count | NK Cells - CD16+ | 5496 | 6864 | 2242 | 7200 | 5685 | 6398 | 3043 | 2011 | 2762 | 3804 | 4131 | 11227 |
| Cell Count | NK Cells - CD56+++ | 441 | 1210 | 503 | 980 | 1070 | 645 | 2383 | 603 | 1006 | 749 | 809 | 416 |
| Cell Count | Plasmablasts | 205 | 42 | 14 | 48 | 41 | 35 | 181 | 201 | 111 | 191 | 196 | 64 |
| Cell Count | T Cells - CD4+ - Central Memory | 21379 | 42599 | 20211 | 28327 | 31748 | 22405 | 25658 | 16736 | 14701 | 14469 | 13649 | 10549 |
| Cell Count | T Cells - CD4+ - Effector Memory | 1833 | 4209 | 1992 | 3032 | 3316 | 2487 | 2536 | 2005 | 1783 | 1817 | 1880 | 1912 |
| Cell Count | T Cells - CD4+ - Naive | 26787 | 42456 | 23311 | 30677 | 33811 | 22978 | 12559 | 7350 | 6877 | 6830 | 6085 | 10005 |
| Cell Count | T Cells - CD8+ - Central Memory | 4950 | 14649 | 6540 | 9552 | 11071 | 7891 | 6999 | 4511 | 4691 | 4790 | 4501 | 3769 |
| Cell Count | T Cells - CD8+ - Effector | 26856 | 39359 | 19205 | 34235 | 32673 | 23444 | 2232 | 1299 | 1490 | 2753 | 2715 | 2638 |
| Cell Count | T Cells - CD8+ - Effector Memory | 1212 | 2106 | 1039 | 1722 | 1677 | 1511 | 687 | 497 | 396 | 665 | 654 | 9289 |
| Cell Count | T Cells - CD8+ - Naive | 19369 | 33860 | 16820 | 26211 | 27760 | 19002 | 8040 | 3824 | 5761 | 7084 | 5999 | 1928 |
| Cell Count | T Cells - CD4- CD8- | 849 | 1320 | 575 | 920 | 1076 | 818 | 493 | 392 | 351 | 402 | 367 | 188 |
| Cell Count | T Cells - CD4+ CD8+ | 153 | 270 | 153 | 126 | 373 | 183 | 38 | 35 | 8 | 21 | 12 | 9 |
| Cell Count | T Cells - Gamma-Delta | 2651 | 4556 | 2037 | 4045 | 4394 | 3536 | 621 | 444 | 408 | 605 | 594 | 2838 |
| Cell Count | Tregs | 1699 | 3316 | 1324 | 2216 | 2217 | 2088 | 976 | 584 | 612 | 621 | 631 | 510 |
| Cell Count | Myeloid Dendritic Cells - Type 1 | 39 | 52 | 30 | 45 | 65 | 47 | 18 | 9 | 6 | 16 | 16 | 25 |
| Cell Count | Myeloid Dendritic Cells - Type 2 | 495 | 1213 | 412 | 838 | 1182 | 753 | 466 | 250 | 346 | 370 | 301 | 418 |
| Cell Count | Plasmacytoid Dendritic Cells | 219 | 539 | 238 | 476 | 607 | 317 | 135 | 60 | 71 | 103 | 62 | 106 |
| % of PBMCs | B Cells - Memory | 0,13 % | 0,19 % | 0,13 % | 0,18 % | 0,20 % | 0,15 % | 0,12 % | 0,06 % | 0,09 % | 0,08 % | 0,09 % | 0,08 % |
| % of PBMCs | B Cells - Naive | 1,75 % | 2,23 % | 1,28 % | 2,24 % | 2,36 % | 1,62 % | 0,37 % | 0,31 % | 0,50 % | 0,51 % | 0,60 % | 1,53 % |
| % of PBMCs | Monocytic MDSCs | 0,44 % | 0,58 % | 0,19 % | 0,22 % | 0,33 % | 0,46 % | 0,14 % | 0,01 % | 0,28 % | 0,08 % | 0,37 % | 0,71 % |
| % of PBMCs | Monocytes - Classical | 4,13 % | 4,42 % | 3,22 % | 5,07 % | 4,89 % | 4,49 % | 5,36 % | 7,19 % | 6,52 % | 7,51 % | 5,49 % | 4,60 % |
| % of PBMCs | Monocytes - Intermediate | 0,32 % | 0,42 % | 0,28 % | 0,42 % | 0,29 % | 0,53 % | 0,44 % | 1,36 % | 0,47 % | 0,98 % | 0,69 % | 0,12 % |
| % of PBMCs | Monocytes - Non-Classical | 0,50 % | 0,42 % | 0,24 % | 0,39 % | 0,30 % | 0,35 % | 0,52 % | 0,48 % | 0,45 % | 0,85 % | 0,78 % | 0,36 % |
| % of PBMCs | NKT Cells | 2,39 % | 2,98 % | 1,74 % | 3,58 % | 2,93 % | 2,60 % | 0,07 % | 0,03 % | 0,07 % | 0,07 % | 0,09 % | 0,14 % |
| % of PBMCs | NK Cells - CD16+ | 0,74 % | 0,67 % | 0,28 % | 0,86 % | 0,63 % | 0,78 % | 0,62 % | 0,45 % | 0,60 % | 0,80 % | 0,81 % | 2,45 % |
| % of PBMCs | NK Cells - CD56+++ | 0,06 % | 0,12 % | 0,06 % | 0,12 % | 0,12 % | 0,08 % | 0,48 % | 0,13 % | 0,22 % | 0,16 % | 0,16 % | 0,09 % |
| % of PBMCs | Plasmablasts | 0,03 % | 0,00 % | 0,00 % | 0,01 % | 0,00 % | 0,00 % | 0,04 % | 0,04 % | 0,02 % | 0,04 % | 0,04 % | 0,01 % |
| % of PBMCs | T Cells - CD4+ - Central Memory | 2,88 % | 4,15 % | 2,54 % | 3,40 % | 3,49 % | 2,73 % | 5,19 % | 3,74 % | 3,22 % | 3,03 % | 2,67 % | 2,31 % |
| % of PBMCs | T Cells - CD4+ - Effector Memory | 0,25 % | 0,41 % | 0,25 % | 0,36 % | 0,36 % | 0,30 % | 0,51 % | 0,45 % | 0,39 % | 0,38 % | 0,37 % | 0,42 % |
| % of PBMCs | T Cells - CD4+ - Naive | 3,61 % | 4,14 % | 2,93 % | 3,68 % | 3,72 % | 2,80 % | 2,54 % | 1,64 % | 1,51 % | 1,43 % | 1,19 % | 2,19 % |
| % of PBMCs | T Cells - CD8+ - Central Memory | 0,67 % | 1,43 % | 0,82 % | 1,15 % | 1,22 % | 0,96 % | 1,41 % | 1,01 % | 1,03 % | 1,00 % | 0,88 % | 0,82 % |
| % of PBMCs | T Cells - CD8+ - Effector | 3,62 % | 3,84 % | 2,42 % | 4,11 % | 3,59 % | 2,86 % | 0,45 % | 0,29 % | 0,33 % | 0,58 % | 0,53 % | 0,58 % |
| % of PBMCs | T Cells - CD8+ - Effector Memory | 0,16 % | 0,21 % | 0,13 % | 0,21 % | 0,18 % | 0,18 % | 0,14 % | 0,11 % | 0,09 % | 0,14 % | 0,13 % | 2,03 % |
| % of PBMCs | T Cells - CD8+ - Naive | 2,61 % | 3,30 % | 2,12 % | 3,14 % | 3,05 % | 2,32 % | 1,63 % | 0,85 % | 1,26 % | 1,48 % | 1,17 % | 0,42 % |
| % of PBMCs | T Cells - CD4- CD8- | 0,11 % | 0,13 % | 0,07 % | 0,11 % | 0,12 % | 0,10 % | 0,10 % | 0,09 % | 0,08 % | 0,08 % | 0,07 % | 0,04 % |
| % of PBMCs | T Cells - CD4+ CD8+ | 0,02 % | 0,03 % | 0,02 % | 0,02 % | 0,04 % | 0,02 % | 0,01 % | 0,01 % | 0,00 % | 0,00 % | 0,00 % | 0,00 % |
| % of PBMCs | T Cells - Gamma-Delta | 0,36 % | 0,44 % | 0,26 % | 0,49 % | 0,48 % | 0,43 % | 0,13 % | 0,10 % | 0,09 % | 0,13 % | 0,12 % | 0,62 % |
| % of PBMCs | Tregs | 0,23 % | 0,32 % | 0,17 % | 0,27 % | 0,24 % | 0,25 % | 0,20 % | 0,13 % | 0,13 % | 0,13 % | 0,12 % | 0,11 % |
| % of PBMCs | Myeloid Dendritic Cells - Type 1 | 0,01 % | 0,01 % | 0,00 % | 0,01 % | 0,01 % | 0,01 % | 0,00 % | 0,00 % | 0,00 % | 0,00 % | 0,00 % | 0,01 % |
| % of PBMCs | Myeloid Dendritic Cells - Type 2 | 0,07 % | 0,12 % | 0,05 % | 0,10 % | 0,13 % | 0,09 % | 0,09 % | 0,06 % | 0,08 % | 0,08 % | 0,06 % | 0,09 % |
| % of PBMCs | Plasmacytoid Dendritic Cells | 0,03 % | 0,05 % | 0,03 % | 0,06 % | 0,07 % | 0,04 % | 0,03 % | 0,01 % | 0,02 % | 0,02 % | 0,01 % | 0,02 % |

| Data | Subset | C2_P6 | C4_P6 | C0_P7 | C2_P7 | C4_P7 | C6_P7 | C0_P8 | C2_P8 | C4_P8 | C0_P9 | C2_P9 | C4_P9 |
|---|---|---|---|---|---|---|---|---|---|---|---|---|---|
| Cell Count | Granulocytes | 326198 | 395405 | 319132 | 323814 | 321135 | 346989 | 374097 | 210386 | 293754 | 278695 | 287349 | 348196 |
| Cell Count | PBMCs | 108844 | 23055 | 46587 | 77520 | 79192 | 47040 | 79854 | 99559 | 77963 | 96458 | 77687 | 49322 |
| % of Total Leukocytes | Granulocytes | 74,98 % | 94,49 % | 87,26 % | 80,68 % | 80,22 % | 88,06 % | 82,41 % | 67,88 % | 79,03 % | 74,29 % | 78,72 % | 87,59 % |
| % of Total Leukocytes | PBMCs | 25,02 % | 5,51 % | 12,74 % | 19,32 % | 19,78 % | 11,94 % | 17,59 % | 32,12 % | 20,97 % | 25,71 % | 21,28 % | 12,41 % |
| Cell Count | B Cells - Memory | 199 | 67 | 357 | 312 | 457 | 359 | 264 | 548 | 353 | 807 | 498 | 150 |
| Cell Count | B Cells - Naive | 6914 | 2773 | 1468 | 2838 | 3872 | 2869 | 3258 | 4515 | 2734 | 3303 | 3009 | 1658 |
| Cell Count | Monocytic MDSCs | 3917 | 5259 | 94 | 421 | 362 | 734 | 684 | 280 | 693 | 2016 | 1038 | 4828 |
| Cell Count | Monocytes - Classical | 25888 | 3526 | 11655 | 25478 | 19581 | 15542 | 15187 | 22676 | 22937 | 20639 | 20883 | 21663 |
| Cell Count | Monocytes - Intermediate | 879 | 152 | 1388 | 1948 | 1746 | 934 | 2501 | 1564 | 2158 | 1199 | 1556 | 935 |
| Cell Count | Monocytes - Non-Classical | 1603 | 42 | 1678 | 2808 | 3089 | 1093 | 1835 | 1885 | 2283 | 2788 | 2439 | 1205 |
| Cell Count | NKT Cells | 1155 | 183 | 507 | 730 | 820 | 388 | 1496 | 1371 | 1365 | 213 | 191 | 64 |
| Cell Count | NK Cells - CD16+ | 18928 | 2508 | 3248 | 4811 | 9073 | 2891 | 8556 | 6446 | 7094 | 8623 | 4601 | 2522 |
| Cell Count | NK Cells - CD56+++ | 504 | 73 | 784 | 877 | 1209 | 619 | 242 | 316 | 265 | 1403 | 625 | 253 |
| Cell Count | Plasmablasts | 51 | 7 | 15 | 16 | 111 | 92 | 36 | 35 | 22 | 36 | 14 | 6 |
| Cell Count | T Cells - CD4+ - Central Memory | 9470 | 1527 | 10433 | 13000 | 13033 | 7922 | 7364 | 8665 | 6059 | 14509 | 10764 | 3282 |
| Cell Count | T Cells - CD4+ - Effector Memory | 2354 | 341 | 1416 | 1702 | 1955 | 935 | 1695 | 1635 | 1423 | 1657 | 1108 | 334 |
| Cell Count | T Cells - CD4+ - Naive | 8238 | 2770 | 3567 | 6402 | 6106 | 4026 | 20837 | 31263 | 16172 | 15322 | 13848 | 5388 |
| Cell Count | T Cells - CD8+ - Central Memory | 4612 | 514 | 2794 | 3262 | 3783 | 2121 | 3754 | 3971 | 3585 | 7581 | 5327 | 1677 |
| Cell Count | T Cells - CD8+ - Effector | 4009 | 501 | 787 | 1510 | 2046 | 710 | 3373 | 3304 | 2992 | 6126 | 4091 | 2180 |
| Cell Count | T Cells - CD8+ - Effector Memory | 13030 | 1746 | 410 | 569 | 777 | 342 | 1141 | 989 | 916 | 1232 | 823 | 373 |
| Cell Count | T Cells - CD8+ - Naive | 1820 | 437 | 3417 | 5845 | 5787 | 3149 | 3538 | 4888 | 3370 | 6002 | 4793 | 1814 |
| Cell Count | T Cells - CD4- CD8- | 251 | 35 | 261 | 771 | 681 | 354 | 428 | 752 | 374 | 271 | 223 | 92 |
| Cell Count | T Cells - CD4+ CD8+ | 11 | 0 | 2 | 14 | 7 | 11 | 14 | 21 | 6 | 50 | 15 | 3 |
| Cell Count | T Cells - Gamma-Delta | 3701 | 502 | 1139 | 1898 | 2488 | 1014 | 1920 | 2177 | 1696 | 823 | 636 | 246 |
| Cell Count | Tregs | 593 | 60 | 667 | 1209 | 1244 | 552 | 615 | 690 | 506 | 790 | 503 | 213 |
| Cell Count | Myeloid Dendritic Cells - Type 1 | 23 | 1 | 32 | 33 | 39 | 16 | 40 | 52 | 55 | 17 | 11 | 4 |
| Cell Count | Myeloid Dendritic Cells - Type 2 | 470 | 6 | 269 | 604 | 490 | 150 | 832 | 1220 | 655 | 505 | 380 | 249 |
| Cell Count | Plasmacytoid Dendritic Cells | 94 | 2 | 163 | 342 | 355 | 156 | 139 | 185 | 171 | 419 | 250 | 152 |
| % of PBMCs | B Cells - Memory | 0,04 % | 0,02 % | 0,09 % | 0,07 % | 0,10 % | 0,08 % | 0,05 % | 0,14 % | 0,08 % | 0,17 % | 0,11 % | 0,03 % |
| % of PBMCs | B Cells - Naive | 1,28 % | 0,63 % | 0,36 % | 0,60 % | 0,82 % | 0,65 % | 0,61 % | 1,11 % | 0,61 % | 0,70 % | 0,68 % | 0,37 % |
| % of PBMCs | Monocytic MDSCs | 0,73 % | 1,19 % | 0,02 % | 0,09 % | 0,08 % | 0,17 % | 0,13 % | 0,07 % | 0,16 % | 0,43 % | 0,24 % | 1,08 % |
| % of PBMCs | Monocytes - Classical | 4,80 % | 0,80 % | 2,84 % | 5,37 % | 4,12 % | 3,54 % | 2,86 % | 5,60 % | 5,14 % | 4,40 % | 4,74 % | 4,86 % |
| % of PBMCs | Monocytes - Intermediate | 0,16 % | 0,03 % | 0,34 % | 0,41 % | 0,37 % | 0,21 % | 0,47 % | 0,39 % | 0,48 % | 0,26 % | 0,35 % | 0,21 % |
| % of PBMCs | Monocytes - Non-Classical | 0,30 % | 0,01 % | 0,41 % | 0,59 % | 0,65 % | 0,25 % | 0,35 % | 0,47 % | 0,51 % | 0,59 % | 0,55 % | 0,27 % |
| % of PBMCs | NKT Cells | 0,21 % | 0,04 % | 0,12 % | 0,15 % | 0,17 % | 0,09 % | 0,28 % | 0,34 % | 0,31 % | 0,05 % | 0,04 % | 0,01 % |
| % of PBMCs | NK Cells - CD16+ | 3,51 % | 0,57 % | 0,79 % | 1,01 % | 1,91 % | 0,66 % | 1,61 % | 1,59 % | 1,59 % | 1,84 % | 1,04 % | 0,57 % |
| % of PBMCs | NK Cells - CD56+++ | 0,09 % | 0,02 % | 0,19 % | 0,18 % | 0,25 % | 0,14 % | 0,05 % | 0,08 % | 0,06 % | 0,30 % | 0,14 % | 0,06 % |
| % of PBMCs | Plasmablasts | 0,01 % | 0,00 % | 0,00 % | 0,00 % | 0,02 % | 0,02 % | 0,01 % | 0,01 % | 0,00 % | 0,01 % | 0,00 % | 0,00 % |
| % of PBMCs | T Cells - CD4+ - Central Memory | 1,76 % | 0,35 % | 2,54 % | 2,74 % | 2,74 % | 1,80 % | 1,39 % | 2,14 % | 1,36 % | 3,09 % | 2,44 % | 0,74 % |
| % of PBMCs | T Cells - CD4+ - Effector Memory | 0,44 % | 0,08 % | 0,35 % | 0,36 % | 0,41 % | 0,21 % | 0,32 % | 0,40 % | 0,32 % | 0,35 % | 0,25 % | 0,07 % |
| % of PBMCs | T Cells - CD4+ - Naive | 1,53 % | 0,63 % | 0,87 % | 1,35 % | 1,29 % | 0,92 % | 3,93 % | 7,72 % | 3,62 % | 3,27 % | 3,14 % | 1,21 % |
| % of PBMCs | T Cells - CD8+ - Central Memory | 0,86 % | 0,12 % | 0,68 % | 0,69 % | 0,80 % | 0,48 % | 0,71 % | 0,98 % | 0,80 % | 1,62 % | 1,21 % | 0,38 % |
| % of PBMCs | T Cells - CD8+ - Effector | 0,74 % | 0,11 % | 0,19 % | 0,32 % | 0,43 % | 0,16 % | 0,64 % | 0,82 % | 0,67 % | 1,31 % | 0,93 % | 0,49 % |
| % of PBMCs | T Cells - CD8+ - Effector Memory | 2,42 % | 0,40 % | 0,10 % | 0,12 % | 0,16 % | 0,08 % | 0,22 % | 0,24 % | 0,21 % | 0,26 % | 0,19 % | 0,08 % |
| % of PBMCs | T Cells - CD8+ - Naive | 0,34 % | 0,10 % | 0,83 % | 1,23 % | 1,22 % | 0,72 % | 0,67 % | 1,21 % | 0,75 % | 1,28 % | 1,09 % | 0,41 % |
| % of PBMCs | T Cells - CD4- CD8- | 0,05 % | 0,01 % | 0,06 % | 0,16 % | 0,14 % | 0,08 % | 0,08 % | 0,19 % | 0,08 % | 0,06 % | 0,05 % | 0,02 % |
| % of PBMCs | T Cells - CD4+ CD8+ | 0,00 % | 0,00 % | 0,00 % | 0,00 % | 0,00 % | 0,00 % | 0,00 % | 0,01 % | 0,00 % | 0,01 % | 0,00 % | 0,00 % |
| % of PBMCs | T Cells - Gamma-Delta | 0,69 % | 0,11 % | 0,28 % | 0,40 % | 0,52 % | 0,23 % | 0,36 % | 0,54 % | 0,38 % | 0,18 % | 0,14 % | 0,06 % |
| % of PBMCs | Tregs | 0,11 % | 0,01 % | 0,16 % | 0,25 % | 0,26 % | 0,13 % | 0,12 % | 0,17 % | 0,11 % | 0,17 % | 0,11 % | 0,05 % |
| % of PBMCs | Myeloid Dendritic Cells - Type 1 | 0,00 % | 0,00 % | 0,01 % | 0,01 % | 0,01 % | 0,00 % | 0,01 % | 0,01 % | 0,01 % | 0,00 % | 0,00 % | 0,00 % |
| % of PBMCs | Myeloid Dendritic Cells - Type 2 | 0,09 % | 0,00 % | 0,07 % | 0,13 % | 0,10 % | 0,03 % | 0,16 % | 0,30 % | 0,15 % | 0,11 % | 0,09 % | 0,06 % |
| % of PBMCs | Plasmacytoid Dendritic Cells | 0,02 % | 0,00 % | 0,04 % | 0,07 % | 0,07 % | 0,04 % | 0,03 % | 0,05 % | 0,04 % | 0,09 % | 0,06 % | 0,03 % |
