## Supplemental Table S3 for "Peripheral blood leukocyte signatures as biomarkers in relapsed ovarian cancer patients receiving combined anti-CD73/anti-PD-L1 immunotherapy in Arm A of the NSGO-OV-UMB1/ENGOT-OV30 trial"

**Online supplemental table S3** Full correlation testing results.

| Subset / Subset_Marker | Normal? | Pearson - r | Spearman - ρ | Pearson - p | Spearman - p | $R^2$ |
|---|---|---|---|---|---|---|
| Granulocytes | Yes | -0,58 | -0,393 | 0,1718 | 0,3956 | 0,337 |
| PBMCs | Yes | 0,58 | 0,393 | 0,1718 | 0,3956 | 0,337 |
| B Cells - Memory | Yes | -0,253 | -0,536 | 0,5838 | 0,2357 | 0,0641 |
| B Cells - Naive | Yes | 0,241 | 0,393 | 0,6019 | 0,3956 | 0,0583 |
| Monocytic MDSCs | Yes | 0,605 | 0,571 | 0,1496 | 0,2 | 0,367 |
| Monocytes - Classical | Yes | -0,694 | -0,464 | 0,0835 | 0,3024 | 0,482 |
| Monocytes - Intermediate | Yes | -0,163 | -0,0357 | 0,7275 | 0,9635 | 0,0265 |
| Monocytes - Non-Classical | Yes | -0,436 | -0,464 | 0,3277 | 0,3024 | 0,19 |
| NKT Cells | No | 0,79 | 0,821 | 0,0344 | 0,0341 | 0,625 |
| NK Cells - CD16+ | Yes | -0,0206 | 0,0357 | 0,9651 | 0,9635 | 0,000423 |
| NK Cells - CD56+++ | Yes | -0,394 | -0,536 | 0,3821 | 0,2357 | 0,155 |
| Plasmablasts | No | -0,0412 | 0,321 | 0,93 | 0,4976 | 0,0017 |
| T Cells - CD4+ - Central Memory | Yes | -0,258 | -0,357 | 0,577 | 0,4444 | 0,0664 |
| T Cells - CD4+ - Effector Memory | Yes | -0,616 | -0,5 | 0,1406 | 0,2667 | 0,38 |
| T Cells - CD4+ - Naive | Yes | -0,19 | -0,143 | 0,6831 | 0,7825 | 0,0361 |
| T Cells - CD8+ - Central Memory | Yes | -0,487 | -0,536 | 0,2679 | 0,2357 | 0,237 |
| T Cells - CD8+ - Effector | No | 0,513 | 0,0714 | 0,2393 | 0,9063 | 0,263 |
| T Cells - CD8+ - Effector Memory | No | -0,297 | -0,286 | 0,5171 | 0,556 | 0,0885 |
| T Cells - CD8+ - Naive | Yes | 0,267 | 0,0714 | 0,5623 | 0,9063 | 0,0714 |
| T Cells - CD4- CD8- | Yes | -0,166 | -0,179 | 0,7219 | 0,7131 | 0,0276 |
| T Cells - CD4+ CD8+ | Yes | 0,0487 | 0,0357 | 0,9175 | 0,9635 | 0,00237 |
| T Cells - Gamma-Delta | Yes | -0,198 | -0,321 | 0,6703 | 0,4976 | 0,0392 |
| Tregs | Yes | 0,37 | 0,179 | 0,4144 | 0,7131 | 0,137 |
| Myeloid Dendritic Cells - Type 1 | No | -0,298 | 0,0357 | 0,5165 | 0,9635 | 0,0887 |
| Myeloid Dendritic Cells - Type 2 | No | -0,37 | -0,607 | 0,4139 | 0,1667 | 0,137 |
| Plasmacytoid Dendritic Cells | Yes | -0,36 | -0,321 | 0,4281 | 0,4976 | 0,129 |
| Granulocytes_CD57 | Yes | -0,066 | 0,0357 | 0,8882 | 0,9635 | 0,00435 |
| PBMCs_CD57 | No | 0,0802 | 0,036 | 0,8642 | 0,9492 | 0,00644 |
| Granulocytes_OX40 | Yes | 0,566 | 0,5 | 0,1849 | 0,2667 | 0,321 |
| PBMCs_OX40 | Yes | -0,0239 | 0,143 | 0,9595 | 0,7825 | 0,00057 |
| Granulocytes_CD38 | Yes | -0,784 | -0,607 | 0,0371 | 0,1667 | 0,614 |
| PBMCs_CD38 | Yes | -0,173 | -0,0714 | 0,7105 | 0,9063 | 0,03 |
| Granulocytes_LAG3 | Yes | -0,195 | 0,143 | 0,6752 | 0,7825 | 0,038 |
| PBMCs_LAG3 | Yes | -0,594 | -0,464 | 0,1594 | 0,3024 | 0,353 |
| Granulocytes_TIGIT | No | 0,684 | 0,541 | 0,0899 | 0,2183 | 0,468 |
| PBMCs_TIGIT | No | 0,739 | 0,401 | 0,058 | 0,4286 | 0,545 |
| Granulocytes_TIM3 | Yes | 0,317 | 0,0357 | 0,4881 | 0,9635 | 0,101 |
| PBMCs_TIM3 | Yes | -0,124 | -0,126 | 0,791 | 0,7929 | 0,0154 |
| Granulocytes_CD86 | Yes | 0,611 | 0,429 | 0,1446 | 0,3536 | 0,374 |
| PBMCs_CD86 | Yes | -0,267 | -0,286 | 0,5621 | 0,556 | 0,0715 |
| Granulocytes_PDL1 | Yes | 0,667 | 0,571 | 0,1019 | 0,2 | 0,445 |
| PBMCs_PDL1 | Yes | 0,606 | 0,321 | 0,1494 | 0,4976 | 0,367 |
| Granulocytes_CD39 | Yes | -0,709 | -0,321 | 0,0742 | 0,4976 | 0,503 |
| PBMCs_CD39 | Yes | 0,054 | -0,321 | 0,9085 | 0,4976 | 0,00291 |
| Granulocytes_IDO1 | Yes | 0,887 | 0,536 | 0,0077 | 0,2357 | 0,787 |
| PBMCs_IDO1 | Yes | 0,859 | 0,464 | 0,0132 | 0,3024 | 0,738 |
| Granulocytes_pCREB | Yes | 0,812 | 0,321 | 0,0265 | 0,4976 | 0,66 |
| PBMCs_pCREB | Yes | 0,47 | 0,5 | 0,2871 | 0,2667 | 0,221 |
| Granulocytes_CD73 | Yes | 0,567 | 0,464 | 0,1848 | 0,3024 | 0,321 |
| PBMCs_CD73 | No | 0,633 | 0,714 | 0,127 | 0,0881 | 0,401 |
| Granulocytes_CTLA4 | | | | | | |
| PBMCs_CTLA4 | | | | | | |
| Granulocytes_GranB | No | 0,441 | 0,408 | 0,3225 | 0,5714 | 0,194 |
| PBMCs_GranB | No | 0,698 | 0,429 | 0,0809 | 0,3536 | 0,488 |
| Granulocytes_pS6 | | | | | | |

| | | | | | | |
|---|---|---|---|---|---|---|
| PBMCs_pS6 | | | | | | |
| Granulocytes_HLADR | No | 0,382 | 0,256 | 0,3979 | 0,5714 | 0,146 |
| PBMCs_HLADR | Yes | 0,0367 | 0,143 | 0,9377 | 0,7825 | 0,00135 |
| Granulocytes_PD1 | | | | | | |
| PBMCs_PD1 | | | | | | |
| B Cells - Memory_CD57 | | | | | | |
| B Cells - Naive_CD57 | | | | | | |
| Monocytic MDSCs_CD57 | | | | | | |
| Monocytes - Classical_CD57 | | | | | | |
| Monocytes - Intermediate_CD57 | No | -0,281 | -0,579 | 0,5421 | 0,1905 | 0,0788 |
| Monocytes - Non-Classical_CD57 | | | | | | |
| NKT Cells_CD57 | No | -0,211 | 0 | 0,6505 | >0,9999 | 0,0443 |
| NK Cells - CD16+_CD57 | Yes | -0,234 | 0 | 0,6129 | >0,9999 | 0,0549 |
| NK Cells - CD56+++_CD57 | | | | | | |
| Plasmablasts_CD57 | Yes | 0,106 | 0,222 | 0,821 | 0,6429 | 0,0112 |
| T Cells - CD4+ - Central Memory_CD57 | | | | | | |
| T Cells - CD4+ - Effector Memory_CD57 | No | -0,311 | -0,0394 | 0,4965 | 0,9524 | 0,097 |
| T Cells - CD4+ - Naive_CD57 | | | | | | |
| T Cells - CD8+ - Central Memory_CD57 | Yes | -0,483 | -0,464 | 0,2725 | 0,3024 | 0,233 |
| T Cells - CD8+ - Effector_CD57 | No | -0,94 | -0,714 | 0,0017 | 0,0881 | 0,883 |
| T Cells - CD8+ - Effector Memory_CD57 | Yes | -0,576 | -0,429 | 0,176 | 0,3536 | 0,332 |
| T Cells - CD8+ - Naive_CD57 | No | -0,104 | -0,0714 | 0,825 | 0,9063 | 0,0107 |
| T Cells - CD4- CD8-_CD57 | No | -0,0467 | 0,0357 | 0,9208 | 0,9635 | 0,00218 |
| T Cells - CD4+ CD8+_CD57 | No | -0,456 | -0,536 | 0,3032 | 0,2357 | 0,208 |
| T Cells - Gamma-Delta_CD57 | No | 0,632 | 0,0714 | 0,1276 | 0,9063 | 0,4 |
| Tregs_CD57 | | | | | | |
| Myeloid Dendritic Cells - Type 1_CD57 | No | -0,0157 | 0,197 | 0,9733 | 0,6857 | 0,000247 |
| Myeloid Dendritic Cells - Type 2_CD57 | | | | | | |
| Plasmacytoid Dendritic Cells_CD57 | No | -0,345 | -0,49 | 0,4491 | 0,2857 | 0,119 |
| B Cells - Memory_OX40 | Yes | 0,19 | 0,0357 | 0,684 | 0,9635 | 0,0359 |
| B Cells - Naive_OX40 | Yes | -0,306 | -0,571 | 0,5046 | 0,2 | 0,0936 |
| Monocytic MDSCs_OX40 | Yes | -0,177 | -0,5 | 0,7036 | 0,2667 | 0,0314 |
| Monocytes - Classical_OX40 | Yes | -0,169 | -0,214 | 0,7169 | 0,6615 | 0,0286 |
| Monocytes - Intermediate_OX40 | Yes | -0,559 | -0,143 | 0,1916 | 0,7825 | 0,313 |
| Monocytes - Non-Classical_OX40 | No | 0,48 | 0,788 | 0,2754 | 0,0571 | 0,231 |
| NKT Cells_OX40 | Yes | -0,0399 | -0,107 | 0,9323 | 0,8397 | 0,00159 |
| NK Cells - CD16+_OX40 | | | | | | |
| NK Cells - CD56+++_OX40 | | | | | | |
| Plasmablasts_OX40 | Yes | -0,318 | -0,464 | 0,4875 | 0,3024 | 0,101 |
| T Cells - CD4+ - Central Memory_OX40 | Yes | 0,37 | 0,357 | 0,4135 | 0,4444 | 0,137 |
| T Cells - CD4+ - Effector Memory_OX40 | Yes | 0,583 | 0,5 | 0,1699 | 0,2667 | 0,339 |
| T Cells - CD4+ - Naive_OX40 | Yes | 0,207 | 0,143 | 0,6562 | 0,7825 | 0,0428 |
| T Cells - CD8+ - Central Memory_OX40 | Yes | 0,257 | 0,214 | 0,5787 | 0,6615 | 0,0658 |
| T Cells - CD8+ - Effector_OX40 | Yes | -0,201 | -0,018 | 0,6664 | 0,9881 | 0,0402 |
| T Cells - CD8+ - Effector Memory_OX40 | Yes | 0,0924 | 0 | 0,8439 | >0,9999 | 0,00853 |
| T Cells - CD8+ - Naive_OX40 | Yes | 0,13 | 0,357 | 0,7811 | 0,4444 | 0,0169 |
| T Cells - CD4- CD8-_OX40 | Yes | 0,152 | 0,0714 | 0,7449 | 0,9063 | 0,0231 |
| T Cells - CD4+ CD8+_OX40 | Yes | -0,212 | -0,429 | 0,6476 | 0,3536 | 0,0451 |
| T Cells - Gamma-Delta_OX40 | Yes | -0,00236 | -0,214 | 0,996 | 0,6615 | 0,00000556 |
| Tregs_OX40 | Yes | 0,099 | 0,357 | 0,8327 | 0,4444 | 0,00981 |
| Myeloid Dendritic Cells - Type 1_OX40 | Yes | 0,42 | 0,571 | 0,3477 | 0,2 | 0,177 |
| Myeloid Dendritic Cells - Type 2_OX40 | Yes | -0,00442 | -0,0357 | 0,9925 | 0,9635 | 0,0000195 |
| Plasmacytoid Dendritic Cells_OX40 | Yes | 0,956 | 0,964 | 0,0008 | 0,0028 | 0,914 |
| B Cells - Memory_CD38 | Yes | -0,175 | 0,0714 | 0,7077 | 0,9063 | 0,0306 |
| B Cells - Naive_CD38 | Yes | -0,273 | -0,429 | 0,5542 | 0,3536 | 0,0743 |
| Monocytic MDSCs_CD38 | Yes | 0,453 | 0,429 | 0,308 | 0,3536 | 0,205 |
| Monocytes - Classical_CD38 | Yes | -0,0194 | 0,0357 | 0,967 | 0,9635 | 0,000378 |
| Monocytes - Intermediate_CD38 | Yes | -0,856 | -0,464 | 0,014 | 0,3024 | 0,733 |
| Monocytes - Non-Classical_CD38 | Yes | 0,119 | 0,536 | 0,7997 | 0,2357 | 0,0141 |

| | | | | | | |
|---|---|---|---|---|---|---|
| NKT Cells_CD38 | No | -0,282 | -0,179 | 0,5406 | 0,7131 | 0,0793 |
| NK Cells - CD16+_CD38 | Yes | 0,305 | -0,0357 | 0,5054 | 0,9635 | 0,0933 |
| NK Cells - CD56+++_CD38 | Yes | -0,158 | -0,107 | 0,7344 | 0,8397 | 0,0251 |
| Plasmablasts_CD38 | Yes | 0,509 | 0,357 | 0,2428 | 0,4444 | 0,26 |
| T Cells - CD4+ - Central Memory_CD38 | Yes | -0,237 | 0,0357 | 0,6083 | 0,9635 | 0,0564 |
| T Cells - CD4+ - Effector Memory_CD38 | Yes | -0,238 | 0,0357 | 0,6069 | 0,9635 | 0,0568 |
| T Cells - CD4+ - Naive_CD38 | No | 0,293 | 0,679 | 0,5238 | 0,1095 | 0,0858 |
| T Cells - CD8+ - Central Memory_CD38 | Yes | 0,382 | 0,25 | 0,3975 | 0,5948 | 0,146 |
| T Cells - CD8+ - Effector_CD38 | No | -0,0237 | 0,286 | 0,9597 | 0,556 | 0,000563 |
| T Cells - CD8+ - Effector Memory_CD38 | Yes | 0,522 | 0,571 | 0,229 | 0,2 | 0,273 |
| T Cells - CD8+ - Naive_CD38 | Yes | -0,604 | -0,821 | 0,1509 | 0,0341 | 0,365 |
| T Cells - CD4- CD8-_CD38 | Yes | -0,21 | 0 | 0,6513 | >0,9999 | 0,0441 |
| T Cells - CD4+ CD8+_CD38 | Yes | 0,481 | 0,679 | 0,2747 | 0,1095 | 0,231 |
| T Cells - Gamma-Delta_CD38 | Yes | 0,182 | -0,0714 | 0,696 | 0,9063 | 0,0331 |
| Tregs_CD38 | No | -0,563 | -0,357 | 0,1885 | 0,4444 | 0,317 |
| Myeloid Dendritic Cells - Type 1_CD38 | Yes | 0,522 | 0,643 | 0,2292 | 0,1389 | 0,273 |
| Myeloid Dendritic Cells - Type 2_CD38 | Yes | 0,341 | 0,143 | 0,4538 | 0,7825 | 0,116 |
| Plasmacytoid Dendritic Cells_CD38 | Yes | 0,542 | 0,321 | 0,2093 | 0,4976 | 0,293 |
| B Cells - Memory_LAG3 | Yes | 0,212 | 0,429 | 0,6482 | 0,3536 | 0,0449 |
| B Cells - Naive_LAG3 | Yes | -0,712 | -0,643 | 0,0729 | 0,1389 | 0,506 |
| Monocytic MDSCs_LAG3 | No | -0,687 | -0,643 | 0,0882 | 0,1389 | 0,472 |
| Monocytes - Classical_LAG3 | Yes | -0,446 | -0,357 | 0,3153 | 0,4444 | 0,199 |
| Monocytes - Intermediate_LAG3 | Yes | -0,772 | -0,536 | 0,0418 | 0,2357 | 0,597 |
| Monocytes - Non-Classical_LAG3 | Yes | -0,642 | -0,393 | 0,1201 | 0,3956 | 0,412 |
| NKT Cells_LAG3 | Yes | 0,1 | 0,296 | 0,8306 | 0,5238 | 0,0101 |
| NK Cells - CD16+_LAG3 | Yes | -0,398 | -0,286 | 0,3766 | 0,556 | 0,158 |
| NK Cells - CD56+++_LAG3 | Yes | -0,245 | -0,0714 | 0,5959 | 0,9063 | 0,0602 |
| Plasmablasts_LAG3 | No | -0,017 | 0,179 | 0,9712 | 0,7131 | 0,000288 |
| T Cells - CD4+ - Central Memory_LAG3 | | | | | | |
| T Cells - CD4+ - Effector Memory_LAG3 | No | -0,0613 | 0,204 | 0,8961 | 0,8571 | 0,00376 |
| T Cells - CD4+ - Naive_LAG3 | No | -0,215 | -0,197 | 0,6433 | 0,6857 | 0,0462 |
| T Cells - CD8+ - Central Memory_LAG3 | No | -0,0613 | 0,204 | 0,8961 | 0,8571 | 0,00376 |
| T Cells - CD8+ - Effector_LAG3 | Yes | -0,455 | -0,536 | 0,3055 | 0,2357 | 0,207 |
| T Cells - CD8+ - Effector Memory_LAG3 | | | | | | |
| T Cells - CD8+ - Naive_LAG3 | Yes | -0,539 | -0,357 | 0,2115 | 0,4444 | 0,291 |
| T Cells - CD4- CD8-_LAG3 | Yes | -0,468 | -0,286 | 0,2896 | 0,556 | 0,219 |
| T Cells - CD4+ CD8+_LAG3 | Yes | -0,109 | -0,179 | 0,8168 | 0,7131 | 0,0118 |
| T Cells - Gamma-Delta_LAG3 | Yes | -0,0327 | 0,0741 | 0,9446 | 0,8929 | 0,00107 |
| Tregs_LAG3 | No | -0,394 | -0,335 | 0,3814 | 0,4762 | 0,156 |
| Myeloid Dendritic Cells - Type 1_LAG3 | Yes | 0,0606 | 0,429 | 0,8973 | 0,3536 | 0,00368 |
| Myeloid Dendritic Cells - Type 2_LAG3 | Yes | -0,611 | -0,714 | 0,1448 | 0,0881 | 0,374 |
| Plasmacytoid Dendritic Cells_LAG3 | Yes | -0,0737 | 0,0714 | 0,8753 | 0,9063 | 0,00543 |
| B Cells - Memory_TIGIT | | | | | | |
| B Cells - Naive_TIGIT | | | | | | |
| Monocytic MDSCs_TIGIT | | | | | | |
| Monocytes - Classical_TIGIT | | | | | | |
| Monocytes - Intermediate_TIGIT | | | | | | |
| Monocytes - Non-Classical_TIGIT | | | | | | |
| NKT Cells_TIGIT | Yes | 0,0314 | 0,179 | 0,9466 | 0,7131 | 0,000989 |
| NK Cells - CD16+_TIGIT | Yes | 0,331 | 0 | 0,4681 | >0,9999 | 0,11 |
| NK Cells - CD56+++_TIGIT | Yes | -0,233 | -0,179 | 0,6149 | 0,7131 | 0,0543 |
| Plasmablasts_TIGIT | No | -0,159 | -0,134 | 0,7335 | 0,8095 | 0,0253 |
| T Cells - CD4+ - Central Memory_TIGIT | Yes | 0,0172 | 0,18 | 0,9708 | 0,7008 | 0,000297 |
| T Cells - CD4+ - Effector Memory_TIGIT | No | 0,14 | 0,482 | 0,764 | 0,2881 | 0,0197 |
| T Cells - CD4+ - Naive_TIGIT | | | | | | |
| T Cells - CD8+ - Central Memory_TIGIT | Yes | 0,402 | 0,679 | 0,3718 | 0,1095 | 0,161 |
| T Cells - CD8+ - Effector_TIGIT | Yes | -0,284 | -0,393 | 0,5377 | 0,3956 | 0,0804 |
| T Cells - CD8+ - Effector Memory_TIGIT | Yes | 0,722 | 0,929 | 0,0672 | 0,0067 | 0,521 |
| T Cells - CD8+ - Naive_TIGIT | No | 0,364 | 0,679 | 0,4218 | 0,1095 | 0,133 |

| | | | | | | |
|---|---|---|---|---|---|---|
| T Cells - CD4- CD8-_TIGIT | No | 0,311 | 0,393 | 0,4966 | 0,3956 | 0,097 |
| T Cells - CD4+ CD8+_TIGIT | Yes | -0,531 | -0,714 | 0,2206 | 0,0881 | 0,281 |
| T Cells - Gamma-Delta_TIGIT | No | -0,478 | -0,179 | 0,2778 | 0,7131 | 0,229 |
| Tregs_TIGIT | Yes | -0,295 | -0,321 | 0,5207 | 0,4976 | 0,087 |
| Myeloid Dendritic Cells - Type 1_TIGIT | No | -0,0649 | 0 | 0,89 | >0,9999 | 0,00422 |
| Myeloid Dendritic Cells - Type 2_TIGIT | | | | | | |
| Plasmacytoid Dendritic Cells_TIGIT | Yes | -0,243 | 0,107 | 0,5995 | 0,8397 | 0,0591 |
| B Cells - Memory_TIM3 | Yes | -0,16 | -0,0901 | 0,7322 | 0,8571 | 0,0255 |
| B Cells - Naive_TIM3 | | | | | | |
| Monocytic MDSCs_TIM3 | Yes | -0,181 | -0,25 | 0,6982 | 0,5948 | 0,0326 |
| Monocytes - Classical_TIM3 | Yes | -0,392 | -0,214 | 0,385 | 0,6615 | 0,153 |
| Monocytes - Intermediate_TIM3 | Yes | -0,555 | -0,393 | 0,1963 | 0,3956 | 0,308 |
| Monocytes - Non-Classical_TIM3 | Yes | -0,473 | -0,357 | 0,2839 | 0,4444 | 0,224 |
| NKT Cells_TIM3 | | | | | | |
| NK Cells - CD16+_TIM3 | Yes | -0,716 | -0,786 | 0,0703 | 0,048 | 0,513 |
| NK Cells - CD56+++_TIM3 | Yes | -0,734 | -0,857 | 0,0603 | 0,0238 | 0,539 |
| Plasmablasts_TIM3 | Yes | 0,43 | 0,357 | 0,3358 | 0,4444 | 0,185 |
| T Cells - CD4+ - Central Memory_TIM3 | | | | | | |
| T Cells - CD4+ - Effector Memory_TIM3 | | | | | | |
| T Cells - CD4+ - Naive_TIM3 | | | | | | |
| T Cells - CD8+ - Central Memory_TIM3 | | | | | | |
| T Cells - CD8+ - Effector_TIM3 | No | -0,501 | -0,612 | 0,2516 | 0,2857 | 0,251 |
| T Cells - CD8+ - Effector Memory_TIM3 | | | | | | |
| T Cells - CD8+ - Naive_TIM3 | No | -0,269 | -0,204 | 0,5603 | 0,8571 | 0,0722 |
| T Cells - CD4- CD8-_TIM3 | Yes | -0,397 | -0,286 | 0,3781 | 0,556 | 0,157 |
| T Cells - CD4+ CD8+_TIM3 | No | -0,394 | -0,429 | 0,3822 | 0,3536 | 0,155 |
| T Cells - Gamma-Delta_TIM3 | | | | | | |
| Tregs_TIM3 | No | -0,501 | -0,612 | 0,2516 | 0,2857 | 0,251 |
| Myeloid Dendritic Cells - Type 1_TIM3 | Yes | -0,864 | -0,679 | 0,0121 | 0,1095 | 0,747 |
| Myeloid Dendritic Cells - Type 2_TIM3 | Yes | -0,636 | -0,429 | 0,1247 | 0,3536 | 0,405 |
| Plasmacytoid Dendritic Cells_TIM3 | Yes | -0,576 | -0,536 | 0,1761 | 0,2357 | 0,332 |
| B Cells - Memory_CD86 | Yes | -0,385 | -0,214 | 0,3941 | 0,6615 | 0,148 |
| B Cells - Naive_CD86 | | | | | | |
| Monocytic MDSCs_CD86 | Yes | -0,629 | -0,75 | 0,1302 | 0,0663 | 0,396 |
| Monocytes - Classical_CD86 | Yes | -0,752 | -0,607 | 0,0512 | 0,1667 | 0,565 |
| Monocytes - Intermediate_CD86 | Yes | -0,336 | -0,179 | 0,4607 | 0,7131 | 0,113 |
| Monocytes - Non-Classical_CD86 | Yes | -0,427 | -0,571 | 0,3396 | 0,2 | 0,182 |
| NKT Cells_CD86 | | | | | | |
| NK Cells - CD16+_CD86 | | | | | | |
| NK Cells - CD56+++_CD86 | | | | | | |
| Plasmablasts_CD86 | No | 0,405 | 0,75 | 0,3677 | 0,0663 | 0,164 |
| T Cells - CD4+ - Central Memory_CD86 | No | 0,441 | 0,408 | 0,3225 | 0,5714 | 0,194 |
| T Cells - CD4+ - Effector Memory_CD86 | | | | | | |
| T Cells - CD4+ - Naive_CD86 | No | 0,746 | 0,612 | 0,0541 | 0,2857 | 0,557 |
| T Cells - CD8+ - Central Memory_CD86 | | | | | | |
| T Cells - CD8+ - Effector_CD86 | | | | | | |
| T Cells - CD8+ - Effector Memory_CD86 | | | | | | |
| T Cells - CD8+ - Naive_CD86 | | | | | | |
| T Cells - CD4- CD8-_CD86 | Yes | -0,557 | -0,357 | 0,1943 | 0,4444 | 0,31 |
| T Cells - CD4+ CD8+_CD86 | Yes | -0,0745 | -0,214 | 0,8738 | 0,6615 | 0,00555 |
| T Cells - Gamma-Delta_CD86 | | | | | | |
| Tregs_CD86 | Yes | 0,255 | 0,429 | 0,5816 | 0,3536 | 0,0648 |
| Myeloid Dendritic Cells - Type 1_CD86 | Yes | -0,399 | -0,357 | 0,3755 | 0,4444 | 0,159 |
| Myeloid Dendritic Cells - Type 2_CD86 | Yes | -0,501 | -0,571 | 0,2525 | 0,2 | 0,251 |
| Plasmacytoid Dendritic Cells_CD86 | No | 0,0858 | 0,393 | 0,8548 | 0,3956 | 0,00737 |
| B Cells - Memory_PDL1 | Yes | 0,469 | 0,321 | 0,2883 | 0,4976 | 0,22 |
| B Cells - Naive_PDL1 | Yes | -0,103 | -0,321 | 0,8258 | 0,4976 | 0,0106 |
| Monocytic MDSCs_PDL1 | Yes | 0,786 | 0,464 | 0,0361 | 0,3024 | 0,618 |
| Monocytes - Classical_PDL1 | Yes | 0,814 | 0,643 | 0,0258 | 0,1389 | 0,663 |

| | | | | | | |
|---|---|---|---|---|---|---|
| Monocytes - Intermediate_PDL1 | Yes | 0,469 | 0,286 | 0,2882 | 0,556 | 0,22 |
| Monocytes - Non-Classical_PDL1 | Yes | 0,587 | 0,321 | 0,1661 | 0,4976 | 0,344 |
| NKT Cells_PDL1 | Yes | 0,103 | 0,357 | 0,8259 | 0,4444 | 0,0106 |
| NK Cells - CD16+_PDL1 | Yes | 0,694 | 0,321 | 0,0837 | 0,4976 | 0,482 |
| NK Cells - CD56+++_PDL1 | Yes | 0,399 | 0,214 | 0,3749 | 0,6615 | 0,159 |
| Plasmablasts_PDL1 | Yes | 0,289 | 0,893 | 0,5292 | 0,0123 | 0,0837 |
| T Cells - CD4+ - Central Memory_PDL1 | Yes | 0,608 | 0,286 | 0,1478 | 0,556 | 0,369 |
| T Cells - CD4+ - Effector Memory_PDL1 | Yes | 0,419 | 0,107 | 0,3489 | 0,8397 | 0,176 |
| T Cells - CD4+ - Naive_PDL1 | Yes | 0,245 | -0,0357 | 0,597 | 0,9635 | 0,0598 |
| T Cells - CD8+ - Central Memory_PDL1 | Yes | 0,299 | 0,143 | 0,5149 | 0,7825 | 0,0894 |
| T Cells - CD8+ - Effector_PDL1 | Yes | 0,467 | 0,321 | 0,2913 | 0,4976 | 0,218 |
| T Cells - CD8+ - Effector Memory_PDL1 | Yes | 0,598 | 0,357 | 0,1559 | 0,4444 | 0,358 |
| T Cells - CD8+ - Naive_PDL1 | Yes | 0,565 | 0,0357 | 0,1858 | 0,9635 | 0,32 |
| T Cells - CD4- CD8-_PDL1 | No | 0,279 | -0,0714 | 0,544 | 0,9063 | 0,0781 |
| T Cells - CD4+ CD8+_PDL1 | Yes | 0,711 | 0,643 | 0,0735 | 0,1389 | 0,505 |
| T Cells - Gamma-Delta_PDL1 | Yes | 0,493 | 0,0357 | 0,2605 | 0,9635 | 0,243 |
| Tregs_PDL1 | Yes | 0,27 | 0,429 | 0,5587 | 0,3536 | 0,0727 |
| Myeloid Dendritic Cells - Type 1_PDL1 | Yes | 0,255 | 0,214 | 0,5808 | 0,6615 | 0,0651 |
| Myeloid Dendritic Cells - Type 2_PDL1 | Yes | 0,768 | 0,464 | 0,0439 | 0,3024 | 0,589 |
| Plasmacytoid Dendritic Cells_PDL1 | No | 0,674 | 0,321 | 0,097 | 0,4976 | 0,454 |
| B Cells - Memory_CD39 | Yes | 0,354 | 0,643 | 0,436 | 0,1389 | 0,125 |
| B Cells - Naive_CD39 | Yes | -0,584 | -0,321 | 0,1685 | 0,4976 | 0,341 |
| Monocytic MDSCs_CD39 | Yes | -0,588 | -0,607 | 0,1651 | 0,1667 | 0,346 |
| Monocytes - Classical_CD39 | Yes | -0,763 | -0,643 | 0,046 | 0,1389 | 0,582 |
| Monocytes - Intermediate_CD39 | Yes | -0,654 | -0,536 | 0,1108 | 0,2357 | 0,428 |
| Monocytes - Non-Classical_CD39 | No | -0,176 | -0,0357 | 0,7065 | 0,9635 | 0,0308 |
| NKT Cells_CD39 | | | | | | |
| NK Cells - CD16+_CD39 | Yes | 0,396 | 0,143 | 0,379 | 0,7825 | 0,157 |
| NK Cells - CD56+++_CD39 | Yes | -0,18 | -0,143 | 0,7001 | 0,7825 | 0,0322 |
| Plasmablasts_CD39 | Yes | 0,544 | 0,429 | 0,2069 | 0,3536 | 0,296 |
| T Cells - CD4+ - Central Memory_CD39 | No | -0,0837 | -0,223 | 0,8585 | 0,6667 | 0,007 |
| T Cells - CD4+ - Effector Memory_CD39 | No | -0,0751 | -0,0394 | 0,8728 | 0,9524 | 0,00565 |
| T Cells - CD4+ - Naive_CD39 | | | | | | |
| T Cells - CD8+ - Central Memory_CD39 | | | | | | |
| T Cells - CD8+ - Effector_CD39 | | | | | | |
| T Cells - CD8+ - Effector Memory_CD39 | No | -0,29 | -0,408 | 0,5275 | 0,5714 | 0,0844 |
| T Cells - CD8+ - Naive_CD39 | | | | | | |
| T Cells - CD4- CD8-_CD39 | No | 0,0113 | 0,355 | 0,9809 | 0,4381 | 0,000127 |
| T Cells - CD4+ CD8+_CD39 | No | 0,506 | 0,222 | 0,247 | 0,6429 | 0,256 |
| T Cells - Gamma-Delta_CD39 | | | | | | |
| Tregs_CD39 | No | -0,167 | -0,679 | 0,7197 | 0,1095 | 0,028 |
| Myeloid Dendritic Cells - Type 1_CD39 | Yes | -0,539 | -0,143 | 0,2114 | 0,7825 | 0,291 |
| Myeloid Dendritic Cells - Type 2_CD39 | Yes | -0,32 | 0,214 | 0,4848 | 0,6615 | 0,102 |
| Plasmacytoid Dendritic Cells_CD39 | Yes | 0,324 | 0,5 | 0,4786 | 0,2667 | 0,105 |
| B Cells - Memory_IDO1 | Yes | 0,409 | 0,286 | 0,3619 | 0,556 | 0,167 |
| B Cells - Naive_IDO1 | No | -0,0539 | 0,214 | 0,9086 | 0,6615 | 0,00291 |
| Monocytic MDSCs_IDO1 | Yes | 0,865 | 0,607 | 0,0119 | 0,1667 | 0,749 |
| Monocytes - Classical_IDO1 | No | 0,922 | 0,714 | 0,0031 | 0,0881 | 0,85 |
| Monocytes - Intermediate_IDO1 | No | 0,868 | 0,571 | 0,0112 | 0,2 | 0,754 |
| Monocytes - Non-Classical_IDO1 | No | 0,935 | 0,75 | 0,002 | 0,0663 | 0,874 |
| NKT Cells_IDO1 | Yes | 0,655 | 0,536 | 0,1101 | 0,2357 | 0,429 |
| NK Cells - CD16+_IDO1 | Yes | 0,871 | 0,607 | 0,0107 | 0,1667 | 0,758 |
| NK Cells - CD56+++_IDO1 | Yes | 0,8 | 0,679 | 0,0309 | 0,1095 | 0,64 |
| Plasmablasts_IDO1 | Yes | -0,0264 | 0 | 0,9551 | >0,9999 | 0,000699 |
| T Cells - CD4+ - Central Memory_IDO1 | No | 0,952 | 0,643 | 0,001 | 0,1389 | 0,906 |
| T Cells - CD4+ - Effector Memory_IDO1 | Yes | 0,859 | 0,714 | 0,0133 | 0,0881 | 0,738 |
| T Cells - CD4+ - Naive_IDO1 | No | 0,932 | 0,802 | 0,0022 | 0,0476 | 0,868 |
| T Cells - CD8+ - Central Memory_IDO1 | No | 0,925 | 0,536 | 0,0028 | 0,2357 | 0,856 |
| T Cells - CD8+ - Effector_IDO1 | Yes | 0,827 | 0,75 | 0,0219 | 0,0663 | 0,683 |

| | | | | | | |
|---|---|---|---|---|---|---|
| T Cells - CD8+ - Effector Memory_IDO1 | Yes | 0,666 | 0,464 | 0,1023 | 0,3024 | 0,444 |
| T Cells - CD8+ - Naive_IDO1 | No | 0,933 | 0,714 | 0,0022 | 0,0881 | 0,87 |
| T Cells - CD4- CD8-_IDO1 | No | 0,74 | 0,45 | 0,057 | 0,3111 | 0,548 |
| T Cells - CD4+ CD8+_IDO1 | No | 0,213 | 0,107 | 0,6471 | 0,8397 | 0,0452 |
| T Cells - Gamma-Delta_IDO1 | Yes | 0,818 | 0,75 | 0,0245 | 0,0663 | 0,669 |
| Tregs_IDO1 | Yes | 0,78 | 0,643 | 0,0386 | 0,1389 | 0,608 |
| Myeloid Dendritic Cells - Type 1_IDO1 | Yes | 0,776 | 0,714 | 0,0401 | 0,0881 | 0,603 |
| Myeloid Dendritic Cells - Type 2_IDO1 | Yes | 0,87 | 0,571 | 0,0108 | 0,2 | 0,758 |
| Plasmacytoid Dendritic Cells_IDO1 | Yes | 0,865 | 0,857 | 0,012 | 0,0238 | 0,747 |
| B Cells - Memory_pCREB | Yes | -0,108 | -0,214 | 0,8169 | 0,6615 | 0,0118 |
| B Cells - Naive_pCREB | Yes | -0,301 | -0,143 | 0,5119 | 0,7825 | 0,0906 |
| Monocytic MDSCs_pCREB | Yes | 0,756 | 0,571 | 0,0494 | 0,2 | 0,571 |
| Monocytes - Classical_pCREB | Yes | 0,866 | 0,821 | 0,0117 | 0,0341 | 0,75 |
| Monocytes - Intermediate_pCREB | Yes | 0,57 | 0,643 | 0,1815 | 0,1389 | 0,325 |
| Monocytes - Non-Classical_pCREB | Yes | 0,391 | 0,429 | 0,386 | 0,3536 | 0,153 |
| NKT Cells_pCREB | Yes | 0,2 | 0,179 | 0,6665 | 0,7131 | 0,0402 |
| NK Cells - CD16+_pCREB | Yes | 0,22 | 0,179 | 0,6359 | 0,7131 | 0,0483 |
| NK Cells - CD56+++_pCREB | Yes | 0,607 | 0,536 | 0,1484 | 0,2357 | 0,368 |
| Plasmablasts_pCREB | Yes | 0,251 | 0,143 | 0,5873 | 0,7825 | 0,063 |
| T Cells - CD4+ - Central Memory_pCREB | Yes | 0,0218 | -0,107 | 0,9629 | 0,8397 | 0,000477 |
| T Cells - CD4+ - Effector Memory_pCREB | Yes | 0,0613 | -0,0357 | 0,8962 | 0,9635 | 0,00376 |
| T Cells - CD4+ - Naive_pCREB | Yes | -0,0772 | -0,143 | 0,8692 | 0,7825 | 0,00597 |
| T Cells - CD8+ - Central Memory_pCREB | Yes | 0,0925 | -0,0714 | 0,8436 | 0,9063 | 0,00856 |
| T Cells - CD8+ - Effector_pCREB | Yes | 0,599 | 0,357 | 0,1555 | 0,4444 | 0,359 |
| T Cells - CD8+ - Effector Memory_pCREB | Yes | 0,298 | 0,286 | 0,5155 | 0,556 | 0,0891 |
| T Cells - CD8+ - Naive_pCREB | Yes | 0,454 | 0,321 | 0,3057 | 0,4976 | 0,206 |
| T Cells - CD4- CD8-_pCREB | No | 0,00772 | 0,0714 | 0,9869 | 0,9063 | 0,0000596 |
| T Cells - CD4+ CD8+_pCREB | Yes | 0,391 | 0,5 | 0,3856 | 0,2667 | 0,153 |
| T Cells - Gamma-Delta_pCREB | Yes | 0,401 | 0,25 | 0,3721 | 0,5948 | 0,161 |
| Tregs_pCREB | Yes | 0,324 | 0,357 | 0,479 | 0,4444 | 0,105 |
| Myeloid Dendritic Cells - Type 1_pCREB | No | -0,445 | -0,643 | 0,317 | 0,1389 | 0,198 |
| Myeloid Dendritic Cells - Type 2_pCREB | Yes | 0,592 | 0,714 | 0,1617 | 0,0881 | 0,35 |
| Plasmacytoid Dendritic Cells_pCREB | Yes | 0,585 | 0,5 | 0,1677 | 0,2667 | 0,342 |
| B Cells - Memory_CD73 | Yes | 0,516 | 0,607 | 0,2362 | 0,1667 | 0,266 |
| B Cells - Naive_CD73 | Yes | -0,0614 | -0,107 | 0,8959 | 0,8397 | 0,00377 |
| Monocytic MDSCs_CD73 | Yes | 0,816 | 0,571 | 0,0251 | 0,2 | 0,667 |
| Monocytes - Classical_CD73 | Yes | 0,898 | 0,929 | 0,006 | 0,0067 | 0,807 |
| Monocytes - Intermediate_CD73 | Yes | 0,666 | 0,607 | 0,1021 | 0,1667 | 0,444 |
| Monocytes - Non-Classical_CD73 | Yes | 0,305 | -0,0714 | 0,5065 | 0,9063 | 0,0928 |
| NKT Cells_CD73 | Yes | 0,96 | 0,893 | 0,0006 | 0,0123 | 0,921 |
| NK Cells - CD16+_CD73 | Yes | 0,388 | 0,143 | 0,3899 | 0,7825 | 0,15 |
| NK Cells - CD56+++_CD73 | Yes | 0,869 | 0,857 | 0,0112 | 0,0238 | 0,755 |
| Plasmablasts_CD73 | Yes | 0,214 | 0,357 | 0,6443 | 0,4444 | 0,046 |
| T Cells - CD4+ - Central Memory_CD73 | Yes | 0,89 | 0,714 | 0,0072 | 0,0881 | 0,793 |
| T Cells - CD4+ - Effector Memory_CD73 | Yes | 0,862 | 0,714 | 0,0127 | 0,0881 | 0,742 |
| T Cells - CD4+ - Naive_CD73 | Yes | 0,886 | 0,786 | 0,0079 | 0,048 | 0,785 |
| T Cells - CD8+ - Central Memory_CD73 | Yes | 0,891 | 0,786 | 0,0072 | 0,048 | 0,793 |
| T Cells - CD8+ - Effector_CD73 | Yes | 0,81 | 0,643 | 0,0271 | 0,1389 | 0,657 |
| T Cells - CD8+ - Effector Memory_CD73 | Yes | 0,902 | 0,786 | 0,0055 | 0,048 | 0,813 |
| T Cells - CD8+ - Naive_CD73 | Yes | 0,922 | 0,786 | 0,0031 | 0,048 | 0,851 |
| T Cells - CD4- CD8-_CD73 | Yes | 0,832 | 0,786 | 0,0203 | 0,048 | 0,692 |
| T Cells - CD4+ CD8+_CD73 | No | -0,0673 | -0,0357 | 0,8859 | 0,9635 | 0,00454 |
| T Cells - Gamma-Delta_CD73 | Yes | 0,875 | 0,821 | 0,0099 | 0,0341 | 0,766 |
| Tregs_CD73 | Yes | 0,802 | 0,643 | 0,0302 | 0,1389 | 0,643 |
| Myeloid Dendritic Cells - Type 1_CD73 | Yes | 0,349 | 0,179 | 0,4431 | 0,7131 | 0,122 |
| Myeloid Dendritic Cells - Type 2_CD73 | Yes | 0,884 | 0,714 | 0,0083 | 0,0881 | 0,781 |
| Plasmacytoid Dendritic Cells_CD73 | Yes | 0,698 | 0,571 | 0,0812 | 0,2 | 0,487 |
| B Cells - Memory_CTLA4 | | | | | | |
| B Cells - Naive_CTLA4 | | | | | | |

| | | | | | | |
|---|---|---|---|---|---|---|
| Monocytic MDSCs_CTLA4 | | | | | | |
| Monocytes - Classical_CTLA4 | | | | | | |
| Monocytes - Intermediate_CTLA4 | Yes | -0,0818 | 0 | 0,8617 | >0,9999 | 0,00668 |
| Monocytes - Non-Classical_CTLA4 | Yes | -0,15 | 0,0371 | 0,7487 | 0,969 | 0,0224 |
| NKT Cells_CTLA4 | | | | | | |
| NK Cells - CD16+_CTLA4 | | | | | | |
| NK Cells - CD56+++_CTLA4 | | | | | | |
| Plasmablasts_CTLA4 | | | | | | |
| T Cells - CD4+ - Central Memory_CTLA4 | | | | | | |
| T Cells - CD4+ - Effector Memory_CTLA4 | | | | | | |
| T Cells - CD4+ - Naive_CTLA4 | | | | | | |
| T Cells - CD8+ - Central Memory_CTLA4 | | | | | | |
| T Cells - CD8+ - Effector_CTLA4 | | | | | | |
| T Cells - CD8+ - Effector Memory_CTLA4 | | | | | | |
| T Cells - CD8+ - Naive_CTLA4 | | | | | | |
| T Cells - CD4- CD8-_CTLA4 | | | | | | |
| T Cells - CD4+ CD8+_CTLA4 | No | 0,746 | 0,612 | 0,0541 | 0,2857 | 0,557 |
| T Cells - Gamma-Delta_CTLA4 | | | | | | |
| Tregs_CTLA4 | Yes | -0,542 | -0,429 | 0,2085 | 0,3536 | 0,294 |
| Myeloid Dendritic Cells - Type 1_CTLA4 | Yes | 0,119 | 0,334 | 0,7988 | 0,4714 | 0,0143 |
| Myeloid Dendritic Cells - Type 2_CTLA4 | | | | | | |
| Plasmacytoid Dendritic Cells_CTLA4 | | | | | | |
| B Cells - Memory_GranB | No | 0,806 | 0,556 | 0,0287 | 0,2119 | 0,649 |
| B Cells - Naive_GranB | | | | | | |
| Monocytic MDSCs_GranB | | | | | | |
| Monocytes - Classical_GranB | | | | | | |
| Monocytes - Intermediate_GranB | No | 0,929 | 0,906 | 0,0025 | 0,0095 | 0,863 |
| Monocytes - Non-Classical_GranB | | | | | | |
| NKT Cells_GranB | Yes | 0,15 | 0,179 | 0,7482 | 0,7131 | 0,0225 |
| NK Cells - CD16+_GranB | Yes | 0,108 | 0,25 | 0,8179 | 0,5948 | 0,0116 |
| NK Cells - CD56+++_GranB | Yes | 0,551 | 0,5 | 0,1997 | 0,2667 | 0,304 |
| Plasmablasts_GranB | Yes | 0,56 | 0,571 | 0,1916 | 0,2 | 0,313 |
| T Cells - CD4+ - Central Memory_GranB | Yes | 0,752 | 0,821 | 0,0513 | 0,0341 | 0,565 |
| T Cells - CD4+ - Effector Memory_GranB | No | -0,288 | -0,0445 | 0,5308 | >0,9999 | 0,083 |
| T Cells - CD4+ - Naive_GranB | Yes | 0,84 | 0,679 | 0,0181 | 0,1095 | 0,705 |
| T Cells - CD8+ - Central Memory_GranB | No | -0,0618 | 0,536 | 0,8953 | 0,2357 | 0,00382 |
| T Cells - CD8+ - Effector_GranB | Yes | -0,284 | -0,179 | 0,5378 | 0,7131 | 0,0804 |
| T Cells - CD8+ - Effector Memory_GranB | Yes | -0,0953 | 0 | 0,8389 | >0,9999 | 0,00909 |
| T Cells - CD8+ - Naive_GranB | No | 0,505 | 0,429 | 0,2474 | 0,3536 | 0,255 |
| T Cells - CD4- CD8-_GranB | Yes | 0,518 | 0,571 | 0,2342 | 0,2 | 0,268 |
| T Cells - CD4+ CD8+_GranB | Yes | -0,076 | -0,107 | 0,8713 | 0,8397 | 0,00578 |
| T Cells - Gamma-Delta_GranB | No | -0,404 | -0,143 | 0,3688 | 0,7825 | 0,163 |
| Tregs_GranB | No | 0,443 | 0,536 | 0,3193 | 0,2357 | 0,196 |
| Myeloid Dendritic Cells - Type 1_GranB | Yes | 0,235 | 0,286 | 0,6118 | 0,556 | 0,0553 |
| Myeloid Dendritic Cells - Type 2_GranB | No | 0,624 | 0,134 | 0,1344 | 0,8095 | 0,389 |
| Plasmacytoid Dendritic Cells_GranB | Yes | 0,312 | 0,357 | 0,4951 | 0,4444 | 0,0976 |
| B Cells - Memory_pS6 | | | | | | |
| B Cells - Naive_pS6 | | | | | | |
| Monocytic MDSCs_pS6 | | | | | | |
| Monocytes - Classical_pS6 | | | | | | |
| Monocytes - Intermediate_pS6 | Yes | -0,594 | -0,5 | 0,1596 | 0,2667 | 0,353 |
| Monocytes - Non-Classical_pS6 | | | | | | |
| NKT Cells_pS6 | No | -0,0213 | 0,144 | 0,9638 | 0,7571 | 0,000454 |
| NK Cells - CD16+_pS6 | Yes | 0,126 | 0,107 | 0,7883 | 0,8397 | 0,0158 |
| NK Cells - CD56+++_pS6 | | | | | | |
| Plasmablasts_pS6 | No | 0,123 | 0,267 | 0,792 | 0,5714 | 0,0152 |
| T Cells - CD4+ - Central Memory_pS6 | | | | | | |
| T Cells - CD4+ - Effector Memory_pS6 | | | | | | |
| T Cells - CD4+ - Naive_pS6 | | | | | | |

| | | | | | | |
|---|---|---|---|---|---|---|
| T Cells - CD8+ - Central Memory_pS6 | | | | | | |
| T Cells - CD8+ - Effector_pS6 | Yes | -0,207 | -0,179 | 0,656 | 0,7131 | 0,0429 |
| T Cells - CD8+ - Effector Memory_pS6 | No | -0,181 | 0,357 | 0,6985 | 0,4444 | 0,0326 |
| T Cells - CD8+ - Naive_pS6 | | | | | | |
| T Cells - CD4- CD8-_pS6 | | | | | | |
| T Cells - CD4+ CD8+_pS6 | Yes | -0,205 | -0,108 | 0,6596 | 0,8222 | 0,0419 |
| T Cells - Gamma-Delta_pS6 | Yes | -0,0592 | -0,108 | 0,8997 | 0,8222 | 0,0035 |
| Tregs_pS6 | | | | | | |
| Myeloid Dendritic Cells - Type 1_pS6 | Yes | -0,562 | -0,714 | 0,1893 | 0,0881 | 0,316 |
| Myeloid Dendritic Cells - Type 2_pS6 | Yes | -0,568 | -0,536 | 0,1833 | 0,2357 | 0,323 |
| Plasmacytoid Dendritic Cells_pS6 | Yes | 0,7 | 0,857 | 0,08 | 0,0238 | 0,49 |
| B Cells - Memory_HLADR | Yes | -0,353 | -0,393 | 0,4369 | 0,3956 | 0,125 |
| B Cells - Naive_HLADR | Yes | -0,304 | -0,286 | 0,5069 | 0,556 | 0,0926 |
| Monocytic MDSCs_HLADR | Yes | -0,191 | -0,143 | 0,6809 | 0,7825 | 0,0367 |
| Monocytes - Classical_HLADR | Yes | -0,515 | -0,429 | 0,2365 | 0,3536 | 0,266 |
| Monocytes - Intermediate_HLADR | Yes | -0,699 | -0,393 | 0,0807 | 0,3956 | 0,488 |
| Monocytes - Non-Classical_HLADR | Yes | -0,265 | -0,214 | 0,5651 | 0,6615 | 0,0705 |
| NKT Cells_HLADR | Yes | 0,336 | 0,556 | 0,4611 | 0,2119 | 0,113 |
| NK Cells - CD16+_HLADR | No | -0,0872 | 0,259 | 0,8525 | 0,5905 | 0,00761 |
| NK Cells - CD56+++_HLADR | Yes | -0,459 | -0,429 | 0,3008 | 0,3536 | 0,21 |
| Plasmablasts_HLADR | Yes | 0,108 | 0,179 | 0,8185 | 0,7131 | 0,0116 |
| T Cells - CD4+ - Central Memory_HLADR | | | | | | |
| T Cells - CD4+ - Effector Memory_HLADR | No | -0,141 | -0,25 | 0,7637 | 0,5948 | 0,0198 |
| T Cells - CD4+ - Naive_HLADR | | | | | | |
| T Cells - CD8+ - Central Memory_HLADR | Yes | 0,424 | 0,5 | 0,3433 | 0,2667 | 0,18 |
| T Cells - CD8+ - Effector_HLADR | No | 0,00162 | 0,214 | 0,9972 | 0,6615 | 0,00000262 |
| T Cells - CD8+ - Effector Memory_HLADR | Yes | 0,458 | 0,286 | 0,3014 | 0,556 | 0,21 |
| T Cells - CD8+ - Naive_HLADR | No | 0,208 | 0,401 | 0,654 | 0,4286 | 0,0434 |
| T Cells - CD4- CD8-_HLADR | No | 0,0238 | 0,0714 | 0,9595 | 0,9063 | 0,000568 |
| T Cells - CD4+ CD8+_HLADR | Yes | -0,514 | -0,571 | 0,2382 | 0,2 | 0,264 |
| T Cells - Gamma-Delta_HLADR | No | -0,277 | -0,144 | 0,5469 | 0,7571 | 0,077 |
| Tregs_HLADR | Yes | -0,532 | -0,536 | 0,2186 | 0,2357 | 0,283 |
| Myeloid Dendritic Cells - Type 1_HLADR | Yes | -0,499 | -0,5 | 0,2542 | 0,2667 | 0,249 |
| Myeloid Dendritic Cells - Type 2_HLADR | Yes | -0,675 | -0,5 | 0,0963 | 0,2667 | 0,455 |
| Plasmacytoid Dendritic Cells_HLADR | Yes | 0,184 | 0,286 | 0,6923 | 0,556 | 0,034 |
| B Cells - Memory_PD1 | | | | | | |
| B Cells - Naive_PD1 | No | -0,0649 | 0 | 0,89 | >0,9999 | 0,00422 |
| Monocytic MDSCs_PD1 | | | | | | |
| Monocytes - Classical_PD1 | | | | | | |
| Monocytes - Intermediate_PD1 | Yes | -0,354 | -0,259 | 0,4362 | 0,5905 | 0,125 |
| Monocytes - Non-Classical_PD1 | | | | | | |
| NKT Cells_PD1 | No | -0,504 | -0,802 | 0,2487 | 0,0476 | 0,254 |
| NK Cells - CD16+_PD1 | | | | | | |
| NK Cells - CD56+++_PD1 | | | | | | |
| Plasmablasts_PD1 | No | -0,454 | -0,315 | 0,3059 | 0,5048 | 0,206 |
| T Cells - CD4+ - Central Memory_PD1 | No | 0,265 | 0,296 | 0,5664 | 0,5238 | 0,07 |
| T Cells - CD4+ - Effector Memory_PD1 | Yes | -0,0865 | 0,179 | 0,8537 | 0,7131 | 0,00748 |
| T Cells - CD4+ - Naive_PD1 | | | | | | |
| T Cells - CD8+ - Central Memory_PD1 | No | -0,0217 | 0 | 0,9631 | >0,9999 | 0,000472 |
| T Cells - CD8+ - Effector_PD1 | No | -0,332 | -0,401 | 0,4674 | 0,4286 | 0,11 |
| T Cells - CD8+ - Effector Memory_PD1 | No | -0,0868 | -0,107 | 0,8532 | 0,8397 | 0,00753 |
| T Cells - CD8+ - Naive_PD1 | No | -0,0649 | 0 | 0,89 | >0,9999 | 0,00422 |
| T Cells - CD4- CD8-_PD1 | No | -0,175 | -0,267 | 0,7073 | 0,5714 | 0,0307 |
| T Cells - CD4+ CD8+_PD1 | Yes | -0,139 | 0,126 | 0,7659 | 0,7929 | 0,0194 |
| T Cells - Gamma-Delta_PD1 | No | 0,288 | 0,0591 | 0,5306 | 0,9048 | 0,0831 |
| Tregs_PD1 | Yes | -0,261 | -0,179 | 0,5719 | 0,7131 | 0,0681 |
| Myeloid Dendritic Cells - Type 1_PD1 | Yes | -0,13 | 0,107 | 0,7812 | 0,8397 | 0,0169 |
| Myeloid Dendritic Cells - Type 2_PD1 | Yes | -0,523 | -0,643 | 0,2285 | 0,1389 | 0,273 |
| Plasmacytoid Dendritic Cells_PD1 | No | -0,136 | 0,0197 | 0,771 | >0,9999 | 0,0185 |
