## Supplemental Table S4 for "Peripheral blood leukocyte signatures as biomarkers in relapsed ovarian cancer patients receiving combined anti-CD73/anti-PD-L1 immunotherapy in Arm A of the NSGO-OV-UMB1/ENGOT-OV30 trial"

**Online supplemental table S4** Results of correlation testing between IDO1 and PD-L1 expression at baseline.

| Subset | Normal? | Pearson - r | Pearson - p | $R^2$ |
|---|---|---|---|---|
| Granulocytes | Yes | 0,791 | 0,0342 | 0,6256 |
| PBMCs | Yes | 0,8252 | 0,0223 | 0,681 |
| B Cells - Memory | Yes | 0,2904 | 0,5276 | 0,08431 |
| B Cells - Naive | Yes | 0,2857 | 0,5345 | 0,08162 |
| Monocytic MDSCs | Yes | 0,9084 | 0,0046 | 0,8252 |
| Monocytes - Classical | Yes | 0,953 | 0,0009 | 0,9082 |
| Monocytes - Intermediate | Yes | 0,8032 | 0,0296 | 0,6452 |
| Monocytes - Non-Classical | Yes | 0,7886 | 0,0351 | 0,6219 |
| NKT Cells | Yes | 0,4798 | 0,2758 | 0,2302 |
| NK Cells - CD16+ | Yes | 0,8312 | 0,0205 | 0,691 |
| NK Cells - CD56+++ | Yes | 0,2908 | 0,527 | 0,08454 |
| Plasmablasts | Yes | -0,35 | 0,4416 | 0,1225 |
| T Cells - CD4+ - Central Memory | Yes | 0,6742 | 0,0967 | 0,4546 |
| T Cells - CD4+ - Effector Memory | Yes | 0,6913 | 0,0854 | 0,4778 |
| T Cells - CD4+ - Naive | Yes | 0,4692 | 0,2881 | 0,2202 |
| T Cells - CD8+ - Central Memory | Yes | 0,5003 | 0,2528 | 0,2503 |
| T Cells - CD8+ - Effector | Yes | 0,8303 | 0,0208 | 0,6894 |
| T Cells - CD8+ - Effector Memory | Yes | 0,4825 | 0,2728 | 0,2328 |
| T Cells - CD8+ - Naive | Yes | 0,7772 | 0,0397 | 0,6041 |
| T Cells - CD4- CD8- | No | 0,7468 | 0,0538 | 0,5578 |
| T Cells - CD4+ CD8+ | Yes | 0,4746 | 0,2818 | 0,2253 |
| T Cells - Gamma-Delta | Yes | 0,8028 | 0,0297 | 0,6445 |
| Tregs | Yes | 0,3271 | 0,474 | 0,107 |
| Myeloid Dendritic Cells - Type 1 | Yes | 0,1326 | 0,7769 | 0,01757 |
| Myeloid Dendritic Cells - Type 2 | Yes | 0,9087 | 0,0046 | 0,8258 |
| Plasmacytoid Dendritic Cells | No | 0,6284 | 0,1307 | 0,3948 |
