## Supplemental Methods for "Peripheral blood leukocyte signatures as biomarkers in relapsed ovarian cancer patients receiving combined anti-CD73/anti-PD-L1 immunotherapy in Arm A of the NSGO-OV-UMB1/ENGOT-OV30 trial"

**Fixation of Whole Blood Using Phosflow LyseFix**

**Foreword:** This protocol is a modified version of the official manufacturer's protocol for the usage of BD Phosflow™ Lyse/Fix buffer (Becton Dickinson, REF 558049). This protocol was followed for the processing and storage of both blood samples taken during the clinical trial, and of healthy donor blood samples used for mass cytometry panel titration and staining controls during this study. All blood samples were collected in 10 mL EDTA Vacutainer® tubes (Becton Dickinson, REF 367525). To save time on heating 1x Lyse/Fix in step 2, one can pre-heat $diH_2O$ to 37°C for use in step 1 beforehand, afterwards keeping the 1x Lyse/Fix buffer in a 37°C water bath for a short time until blood samples are ready to be processed. It is also advisable to pre-cool the centrifuge, first used in step 7, to 4°C prior to starting the protocol. Furthermore, at least 50 mL of saline (0,9% w/V (g/L) NaCl in deionized water) per 3-3,3 mL of blood to be processed should be prepared and cooled to 4°C prior to starting the protocol.

1. For each 3-3,3 mL of whole blood to be processed, prepare 40 mL of 1x Lyse/Fix buffer in a conical 50 mL tube by mixing 8 mL of 5x Lyse/Fix buffer and 32 mL of deionized water (henceforth "$diH_2O$").

**NOTE:** Perform steps involving the Lyse/Fix buffer and samples containing it in a fume hood due to the Lyse/Fix buffer containing (para)formaldehyde.

2. Heat the 1x Lyse/Fix buffer to 37°C in a water bath.
3. Gently invert the tubes containing the blood 8-10 times to resuspend the cells. Do not shake the tubes.
4. Gently transfer 3-3,3 mL of blood to each conical 50 mL tube containing 40 mL of pre-heated 1x Lyse/Fix.
5. Mix the samples by inverting the tubes multiple times.
6. Incubate the samples in a water bath at 37°C for 10 min.
7. Centrifuge the samples (500 G, 8 min, 4°C).
8. Decant supernatant into a (para)formaldehyde-appropriate waste container.

**NOTE:** Keep the samples cold / on ice from this point forward.

9. Gently vortex the samples to resuspend the cell pellets. If necessary, add 1 mL of saline (4°C) before resuspending.

**NOTE:** If there are blood clots present in the sample, follow the following steps:

   a) Add 10 mL of saline (4°C) to the sample.
   b) Attempt to break up the clots and resuspend the cells by pipetting and/or vortexing.
   c) Filter the sample through a 40 µm mesh into a new conical 50 mL tube.
   d) Add 30 mL of saline (4°C) to the old tube.
   e) Invert the old tube several times to wash out any remaining leukocytes.
   f) Pour the washout into the new tube through the 40 µm mesh, washing out the mesh in the process.
   g) Centrifuge (500 G, 8 min, 4°C).
   h) Decant supernatant.
   i) Continue from step 9.

10. Add 45 mL of saline (4°C) to each sample.
11. Vortex the samples to resuspend and mix.

12. Centrifuge the samples (500 G, 8 min, 4°C).
13. Decant supernatant into a (para)formaldehyde-appropriate waste container.
14. Add 1 mL of saline (4°C) to each sample.
15. Gently vortex the samples to resuspend the cell pellets.

**NOTE:** If multiple conical 50 mL tubes contain leukocytes from the same blood sample, combine them for a simpler counting procedure by following steps 16-18. Keep track of the volumes of the combined leukocyte resuspensions.

16. Combine the resuspensions made in step 15 belonging to the same blood sample in a new tube.
17. Add 1 mL of saline (4°C) to each old tube.
18. Wash out each old tube and transfer the washouts to the new tube with the combined cell resuspensions.
19. Count the cells using your preferred cell counting method, using saline (4°C) to dilute and resuspend samples in case of excessive cell density.
20. Ideally, samples should be frozen at a maximum cell density of $1*10^7$ cells / mL. Therefore, prepare a number of cryogenic storage vials that enables the storage of processed leukocyte samples at this maximal cell density.
21. Transfer the samples to the cryogenic storage vials.
22. Store the vials at -80°C.

### *In Vitro* PBMC Stimulation

**Foreword:** Work in a sterile environment. All solutions are presumed sterile and at room temperature (RT) unless otherwise specified. The density gradient medium used in this study was Lymphoprep™ (Stemcell Technologies, Cat.No. 07861). Prepare cold PBMC isolation buffer beforehand - dissolve EDTA in phosphate-buffered saline (PBS) to a 2 mM concentration; Add bovine serum albumin to a concentration of 0,5% w/V (5 mg/mL); Adjust the pH to 7.2 and sterilize the solution by filtering it through a cellulose acetate syringe filter with a 0,2 µm pore size. Prepare cell culture medium beforehand - RPMI1640 cell culture medium containing 10% V/V fetal calf/bovine serum, 2 mM L-glutamine, 100 UI/mL penicillin and 100 U/mL interleukin-2 (1 µg of interleukin-2 contains 18.000 U); warm the medium to 37°C prior to the start of the protocol. Myeloid cells may likely not be present in the final PBMC isolate.

#### 1 - Isolation of peripheral blood mononuclear cells (PBMCs) by density gradient centrifugation

1. Determine the total volume of the collected whole blood intended for PBMC isolation.
2. For every 1 mL of blood, aliquot 1 mL of density gradient medium into a conical 50 mL tube (**NOTE:** To avoid overflowing in future protocol steps, each conical 50 mL tube should contain a maximum of 15 mL of density gradient medium).
3. Dilute the blood with PBS at a 1:1 ratio.
4. Distribute the diluted blood into the tubes containing density gradient medium by very slowly dispensing the diluted blood on top of the density gradient medium. Dispense 2 mL of diluted blood for every 1 mL of density gradient medium (max. 30 mL of diluted blood on top of 15 mL of density gradient medium) (**NOTE:** Hold the tube at a low angle and slowly dispense the diluted blood along the inner wall of the tube, close to the surface of the density gradient medium. At first, dispense the diluted blood as slowly as possible, after which you may carefully increase dispensing speed as the layer of diluted blood builds up on top of the density gradient medium. The diluted blood will somewhat mix with the top of the density gradient medium at the very start of dispensation; however, clear separation should occur once more blood is dispensed).
5. Centrifuge the samples (400 G, 25 min, RT, brakes off, minimal acceleration). If more than two hours have passed since the blood was sampled, increase centrifugation time to 30 minutes.
6. After centrifugation, PBMCs will form an opaque interphase/band between the yellow plasma layer on top and the clear density gradient medium layer below. Erythrocytes will have settled to the bottom of the tube. Transfer as much of the PBMCs into new conical 50 mL tubes (**NOTE:** Transferring small quantities of the plasma and density gradient medium along with the PBMCs to achieve the highest possible PBMC recovery should not present an issue in the following steps. Do not combine PBMCs from the same blood donor - this will be done in a later step).
7. Fill the conical 50 mL tubes containing the PBMCs to 45 mL with PBS.
8. Mix the samples gently by inverting the tubes.
9. Centrifuge the samples (500 G, 5 min, RT).
10. During centrifugation, prepare fresh 1x Red Blood Cell Lysis Solution by diluting 10x Red Blood Cell Lysis Solution (Miltenyi Biotec, Cat.No. 130-094-183) in distilled water (**NOTE:** Do not use deionized water).
11. Remove supernatant by aspiration. Do not decant, as the pellet is not cohesive enough.
12. Loosen the cell pellets by gentle vortexing / flicking of the tubes.
13. Add 2 mL of PBS to each sample.
14. Vortex or pipet the samples gently to resuspend.

15. Add 20 mL of 1x Red Blood Cell Lysis Solution to each sample.
16. Mix the samples gently by inverting the tubes.
17. Incubate the samples at room temperature for 8 minutes.
18. Fill the conical 50 mL tubes containing the PBMCs to 45 mL with PBMC isolation buffer.
19. Mix the samples gently by inverting the tubes.
20. Centrifuge the samples (300 G, 10 min, RT).
21. Remove supernatant by aspiration. Do not decant, as the pellet is not cohesive enough.

**NOTE:** If there are multiple tubes containing PBMCs from the same blood donor, follow steps 22-25. Otherwise, skip to step 26.

22. Add 1 mL of PBMC isolation buffer to each sample.
23. Resuspend PBMC pellets gently by pipetting.
24. Transfer all PBMCs from the same blood donor to a new conical 50 mL tube.
25. Wash out each old tube with 1 mL of PBMC isolation buffer and transfer the washout to the new tube.
26. Fill the conical 50 mL tubes containing the PBMCs to 40 mL with PBMC isolation buffer.
27. Resuspend and mix by pipetting.
28. Filter the samples through 40 μm meshes into new conical 50 mL tubes.
29. Wash out each filter mesh with 5 mL of PBMC isolation buffer.
30. Count the cells in each sample using your preferred cell counting method, using PBMC isolation buffer to make sample dilutions in case of excessive cell density.
31. Centrifuge the samples (300 G, 10 min, RT).

### 2 - *In vitro* stimulation of PBMCs

32. Resuspend the PBMCs to a cell density of $2*10^6$ cells/mL in the cell culture medium.
33. Distribute the PBMC suspension into ventilated cell culture flasks intended for suspension cell culture, having between 0,2 and 0,5 mL of cell suspension per $cm^2$ of flask surface area (**NOTE:** It is advisable to set aside a smaller quantity of PBMCs into a separate cell culture flask to serve as an unstimulated control for visual comparison during incubation).
34. Incubate the PBMCs in an incubator (37°C, 5% V/V $CO_2$, humidified) for 1 hour to acclimate them to the medium.
35. Add phytohemagglutinin-P (Sigma, Cat.No. 61764) (dissolved in PBS to a concentration of 1 μg/μL) to the (non-control) PBMC suspensions to a final concentration of 2,5 μg/mL.
36. Incubate the PHA-P-stimulated PBMCs for 48 hours in an incubator (37°C, 5% V/V $CO_2$, humidified).
37. View the stimulated and control PBMCs under a microscope every 24 hours to confirm (non-)stimulation. Stimulated PBMCs should have formed aggregates already before 24 hours have elapsed, while controls cells will remain in a single-cell suspension throughout the 48-hour incubation period:

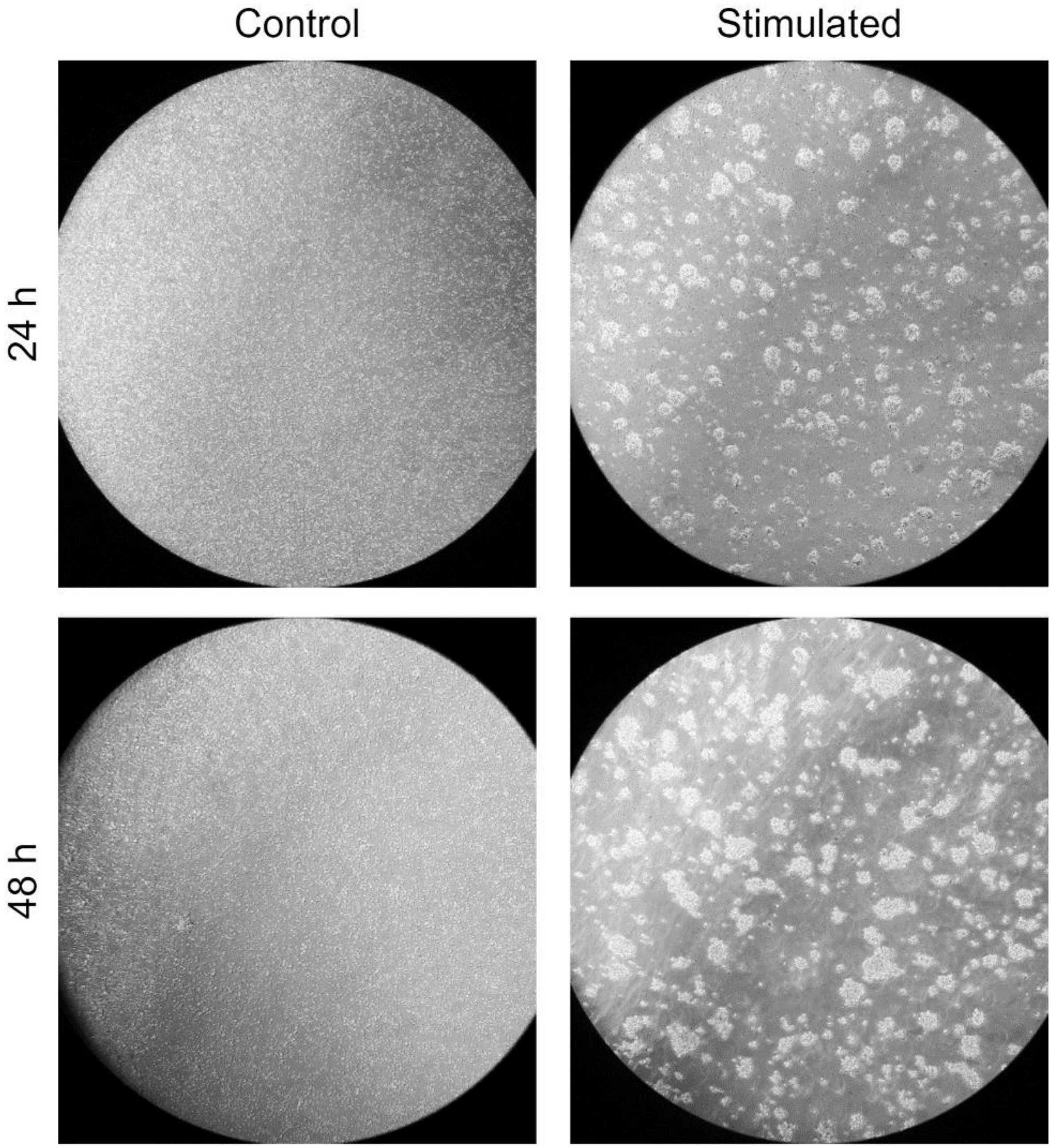


#### 3 - Collection of *in vitro* stimulated PBMCs

**NOTE:** From this point onward, it is no longer necessary to work in a sterile environment. Additionally, control cells can be discarded after visualization.

38. Prepare 0,4 mL of ice-cold 2 mM EDTA solution in PBS (henceforth “EDTA solution”) per $cm^2$ of the surface of the cell culture flasks used in the incubation of the stimulated PBMCs.
39. Resuspend the cultured PBMCs by pipetting.
40. Transfer the cell culture medium containing resuspended PBMCs to conical 50 mL tubes.
41. Immediately add 0,15 mL of ice-cold 2 mM EDTA solution in PBS per $cm^2$ of the surface of the emptied cell culture flasks.
42. Place the cell culture flasks containing EDTA solution on ice.

43. Check cell detachment under a microscope after 5 and 10 minutes.
44. If the cells have not sufficiently detached after 10 minutes, gently use a cell scraper to detach them manually.
45. Thoroughly resuspend the EDTA-treated cells and transfer the resuspensions to the conical 50 mL tubes.
46. Centrifuge the tubes (300 G, 5 min, RT).
47. Aspirate supernatant.
48. Add 10 mL of EDTA solution to each tube.
49. Pipet to resuspend the cell pellets.
50. Centrifuge the tubes (300 G, 5 min, RT).
51. Aspirate supernatant.
52. Add 1 mL of cold PBS to each cell pellet.
53. Pipet to resuspend the cell pellets.

**NOTE:** If there are multiple tubes containing PBMCs from the same blood donor, follow steps 54 and 55. Otherwise, skip to step 56.

54. Transfer all PBMCs from the same blood donor into a new tube.
55. Wash out each old tube with 1 mL of cold PBS and transfer the washout to the new tube.
56. Count the cells in each sample using your preferred cell counting method, using cold PBS to make sample dilutions in case of excessive cell density.
57. Resuspend the samples to the desired cell density of using cold PBS and continue with cell fixation (supplemental protocol “Fixation of Whole Blood Using BD Phosflow LyseFix” in the online supplemental appendix) (**NOTE:** The density of stimulated PBMC at the start of the fixation procedure in this study was set to $4{,}55*10^6$ cells/mL, mimicking the lower end of the range of leukocytes per mL of human blood. However, it is advised to use a higher cell density to avoid cell losses during the fixation process).

**Sample Barcoding and Staining**

**Foreword:** This protocol presumes that leukocytes had been fixed using a (para)formaldehyde-based method and stored in 1 mL aliquots in cryogenic storage vials at -80°C according to the supplemental protocol “Fixation of Whole Blood Using Phosflow LyseFix”. Furthermore, this protocol presumes that the antibody mixes for surface staining and intracellular staining had been prepared, aliquoted according to pre-calculated sample batch cell numbers and frozen prior to the start. While performing the protocol, it is good practice to re-cap the round-bottom 5 mL tubes after every step in order to mitigate sample loss in case of tube mishandling. Barcoding in the protocol is done using palladium isotopes (Standard BioTools, Cat.No. 201060). If there are multiple sample batches, to minimize batch effects it is highly advisable to distribute samples among batches in a way that avoids one or more batches consisting of only/mostly samples with identical parameters (study site, patient, treatment arm, sampling timepoint, etc.). Additionally, it is advisable to include at least one anchor sample in each batch for batch effect correction during data clean-up. A second anchor sample in each batch can be used for the confirmation of efficacy of batch correction done according to the first anchor sample.

**1 - Barcoding**

**1.1 - Thawing fixed cells**

1. For each sample, pre-label one round-bottom 5 mL polystyrene test tube with a cell strainer cap (Corning, Cat.No. 352235) (henceforth “round-bottom 5 mL tube”).
2. For each sample, prepare 3 mL of DNAse I (Sigma, SKU: DN25) dissolved in Dulbecco’s phosphate-buffered saline containing $Ca^{2+}$ and $Mg^{2+}$ (Sigma, SKU: D8662) to a final concentration of 0.25 mg/mL (henceforth “DNAse”). The purpose of the DNAse is the degradation of any extranuclear DNA that may lead to the formation of cell aggregates.
3. Aliquot 2 mL of DNAse into each round-bottom 5 mL tube. Condition the DNAse to room temperature (RT).
4. Thaw the samples intended to comprise a batch by floating the cryogenic storage vials in cold tap water.

**NOTE:** Perform steps 5-7 on one cryogenic storage vial at a time.

5. Transfer the contents of the cryogenic storage vial to the corresponding round-bottom 5 mL tube containing DNAse.
6. Wash out the cryogenic storage vial with 1 mL of DNAse and transfer the washout to its corresponding round-bottom 5 mL tube.
7. Incubate cells in DNAse at RT for a minimum of 5 min.
8. Centrifuge the round-bottom 5 mL tubes with the DNAse/sample mix (800 G, 5 min, RT).
9. Decant supernatant (**NOTE:** After decanting, before rotating the round-bottom 5 mL tube upright, wick away the last of the supernatant by dabbing the rim of the round-bottom 5 mL tube on a paper towel).
10. Vortex the samples to resuspend the cells in the remaining liquid volume.
11. Add 2 mL of MaxPar Cell Staining Buffer (CSB) (Standard BioTools, Cat.No. 201068) to each sample and place it on ice.
12. Label a new round-bottom 5 mL tube for each sample.
13. Transfer each sample to the corresponding new round-bottom 5 mL tube through the cap filter, 1 mL at a time (**NOTE:** Place the opening of the pipette tip directly against the strainer mesh, being careful not to force the mesh out of the cap).
14. Wash out each old round-bottom 5 mL tube with 1 mL of MaxPar CSB and transfer the washout through the cap of the corresponding new round-bottom 5 mL tube.

15. Rinse each cap filter with 1 mL of MaxPar CSB (**NOTE:** While dispensing the MaxPar CSB, drag the pipette tip around the mesh to ensure a thorough washout of the filter).
16. Centrifuge the samples (800 G, 5 min, RT).
17. Decant supernatant.
18. Place the samples on ice while counting the cells in the following steps.
19. Vortex the sample(s) currently intended for cell counting to finely resuspend the cells.
20. Count the cells in each sample using your preferred cell counting method, using MaxPar phosphate-buffered saline (PBS) (Standard BioTools, Cat.No. 201058) to dilute and resuspend samples in case of excessive cell density (**NOTE:** The cell counting in this study was done electronically, using two 10µL aliquots from each sample on the Countess™ II automated cell counter (ThermoFisher Scientific, Cat.No. AMQAX1000) for each sample).
21. Once finished with counting the cells in a sample, place said sample back on ice.
22. For the barcoding process, if a sample contains fewer than $1*10^6$ cells, the volumes of the barcoding reagents should be adjusted according to the manufacturer's protocol. Otherwise, if a sample contains more than $3.5*10^6$ cells, $3.5*10^6$ cells should be transferred to a new round-bottom 5 mL tube so the excess can either be re-frozen or barcoded as a separate sample.
23. After establishing samples containing appropriate numbers of cells ($1 - 3.5*10^6$), centrifuge the samples (800 G, 5 min, RT). At this point, any excess samples can also be centrifuged to prepare them for storage.
24. During centrifugation, thaw the palladium barcodes at RT.
25. During centrifugation, prepare 1 mL of 1x Barcode Perm Buffer per sample from nine parts MaxPar PBS and one part 10x Barcode Perm Buffer (Standard BioTools, Cat.No. 202157, included in barcoding kit) by volume (i.e. 900 µL of MaxPar PBS and 100 µL of 10x Barcode Perm Buffer per sample).
26. Decant supernatant. If there are any excess samples intended for storage, place them on ice and process them for storage later.

**1.2 - Barcoding cells**

27. Vortex the samples to resuspend the cells in the remaining liquid volume.
28. Add 800 µL of 1X Barcode Perm Buffer to each sample.
29. Vortex the samples to mix.
30. Resuspend each palladium barcode in 100 µL of 1X Barcode Perm Buffer and transfer it to the corresponding sample.
31. Immediately pipet the sample to mix in the barcode completely.
32. Incubate samples in the barcode reagent at RT for a minimum of 45 min. During incubation, either keep the samples agitated or vortex the samples periodically.
33. During incubation, store or discard any excess samples.
34. Once the incubation time elapses, centrifuge the samples (800 G, 5 min, RT).
35. Decant supernatant.
36. Vortex the samples to resuspend the cells in the remaining liquid volume.
37. Add 2 mL of MaxPar CSB to each sample.
38. Vortex the samples to mix.
39. Centrifuge the samples (800 G, 5 min, RT).

40. Decant supernatant.
41. Vortex the samples to resuspend the cells in the remaining liquid volume.
42. Once more, add 2 mL of MaxPar CSB to each sample.
43. Vortex the samples to mix.
44. Centrifuge the samples (800 G, 5 min, RT).
45. Decant supernatant.
46. Resuspend each sample using 100 µL of MaxPar PBS (**NOTE:** The same pipette tip can now be used for all samples. Here, it is recommended to use a pipette (tip) with more than 100 µL capacity due to the need to include the volume of the sample in future transferring steps).
47. Transfer all of the resuspended samples to a single 15 mL conical tube. For the purposes of accurate cell counting in the next step, keep track of the total volume transferred to the 15 mL conical tube.
48. Wash out each old round-bottom 5 mL tube with 100 µL of MaxPar PBS and transfer the washout to the 15 mL conical tube, keeping track of the total volume.
49. Once more, wash out each old round-bottom 5 mL tube with 100 µL of MaxPar PBS and transfer the washout to the 15 mL conical tube, keeping track of the total volume.

**1.3 - Freezing barcoded cells**

50. Count the cells in the 15 mL conical tube containing the barcoded sample batch using your preferred cell counting method.
51. Ideally, the barcoded sample batch should be frozen at a maximum cell density of $1*10^7$ cells / mL. Therefore, if necessary, centrifuge the batch (800 G, 5 min, RT), decant the supernatant and re-adjust the volume using MaxPar PBS.
52. Pipet or vortex the samples gently to resuspend the cells.
53. Transfer the sample batch to one or more cryogenic storage vials.
54. Store the vials at -80°C.

**NOTE:** Freezing a barcoded sample batch enables the staining to be done at a later point in time.

### 2 - Staining

#### 2.1 - Thawing barcoded cells

55. For each cryogenic storage vial to be thawed, pre-label one round-bottom 5 mL tube with a cell strainer cap.
56. Thaw barcoded sample batches by floating the cryogenic storage vials in cold tap water.

**NOTE:** Steps 2-17 of this protocol can be repeated here if the samples contain cell aggregates.

**NOTE:** Perform steps 57-61 on one cryogenic storage vial at a time. If a batch is split across multiple cryogenic storage vials, keep the parts of the batch separate throughout the rest of the staining protocol. This is necessary to be able to fit the samples in 5 mL round-bottom 5 mL tubes and use the strainer cap mesh for filtering cell aggregates.

57. Transfer the contents of the cryogenic storage vial into the appropriately marked round-bottom 5 mL tube.
58. Wash out the cryogenic storage vial with 1 mL of MaxPar CSB and transfer the washout to the corresponding round-bottom 5 mL tube.
59. Once more, wash out the cryogenic storage vial with 1 mL of MaxPar CSB and transfer the washout to the corresponding round-bottom 5 mL tube.
60. Pipet to mix and resuspend the contents of the round-bottom 5 mL tube (i.e. sample).
61. Place the sample on ice.
62. Centrifuge the samples (800 G, 5 min, RT).
63. During centrifugation, retrieve aliquots of the surface staining antibody mix from -80°C and place on ice to thaw.
64. During centrifugation, label a new round-bottom 5 mL tube for each sample.
65. Decant supernatant.

#### 2.2 - Surface staining

66. Vortex the samples to resuspend the cells in the remaining liquid volume.
67. Add MaxPar CSB to each sample to dilute the cells to a density appropriate for counting (e.g., 3 mL).
68. Vortex the samples to mix.
69. Count the cells in each sample using your preferred cell counting method.
70. Transfer a number of cells from each sample rounded to a multiple of $1{,}5*10^6$ to the corresponding new round-bottom 5 mL tube labeled in step 64. Keep track of the number of cells transferred to each tube throughout the staining protocol (**NOTE:** Rounding the number of cells in each sample to a multiple of 1,5 million simplifies calculation of reagent volumes and the attainment of the staining cell density of 3 million cells / 100 µL. Instead of keeping track of the cell count in each sample, one can keep track of *N* for each sample, where *N* represents the number of multiples of 1,5 million cells in each sample).
71. Centrifuge the samples (800 G, 5 min, RT).
72. Decant away as much supernatant as possible without compromising the sample.

**NOTE:** Perform steps 73-77 on one sample at a time. The same pipette tip should be used when manipulating cells in order to mitigate cell losses. This pipette tip should be prevented from drying out.

73. Gently vortex the sample to resuspend the cells in the remaining liquid volume.

74. Carefully, not introducing bubbles, measure the volume of the resuspension using a pipette: Adjust the pipette to its lowest volume, pipet this minimal volume into the pipette tip, position the pipette tip at the bottom of the tube, and adjust the pipetting volume upwards up until the entirety of the sample is aspirated. Write down the volume on the pipette and pipet the sample back out into the tube (**NOTE:** This pipette tip should not be discarded and should be used for manipulating the cells in step 73).
75. Using a separate pipette (tip), prepare an aliquot of MaxPar CSB equal to the amount needed to bring the measured cell volume to [*N* * 20 µL], plus 10 µL of volume as a buffer.
76. Use the pipette tip from step 74 to adjust the volume of the cell resuspension with aliquoted MaxPar CSB to [*N* * 20 µL] (**NOTE:** Keep in mind the current volume of the sample, measured in step 74. To wash as many cells as possible out of the pipette tip, aspirate a volume of MaxPar CSB larger than the one measured in step 74 one or more times).
77. Vortex the sample gently to mix.
78. Taking into account the total cells in all samples, mix together [*N* * 2,5 µL] of both 1000 U/mL heparin (Ratiopharm, Reg.No. 5394.00.00) and human FcR Blocking Reagent (Miltenyi Biotec, Cat.No. 130-059-901) (e.g., if there is a total of $30*10^6$ (20 x $1,5*10^6$) cells across all samples, mix together 50 µL (20 x 2,5 µL) of both heparin and human FcR Blocking Reagent together in a new tube, for a total mix volume of 100 µL). Add either 10 µL or 10% of volume on top as a volume buffer (**NOTE:** Heparin mitigates the unspecific accumulation of positively-charged metal-conjugated antibodies on eosinophils, while Fc Blocking Reagent blocks the unspecific binding of CyTOF antibodies to human Fc receptors).
79. Add [*N* * 5 µL] of the heparin / human FcR Blocking Reagent mix to each sample.
80. Vortex the samples gently to mix.
81. Incubate the samples at RT for 20 min. During incubation, either keep the samples agitated or vortex the samples periodically.
82. During incubation, spin down surface staining antibody mix aliquots in a desktop centrifuge.
83. Ensure there are no or few bubbles in the samples. Pulse the samples at 800 G three times to attempt to remove excess bubbles.
84. Add [*N* * 25 µL] of surface staining antibody mix to the samples. Save leftover antibody mix on ice for same-day staining of control beads according to the supplemental protocol “Staining of Control Beads” in the online supplemental appendix.
85. Vortex the samples gently to mix.
86. Incubate the samples at RT for 30 min. During incubation, either keep the samples agitated or vortex the samples periodically.
87. Add 3 mL of MaxPar CSB to each sample.
88. Vortex the samples to mix.
89. Centrifuge the samples (800 G, 5 min, RT).
90. Decant supernatant.
91. Vortex the samples to resuspend the cells in the remaining liquid volume.
92. Once more, add 3 mL of MaxPar CSB to each sample.
93. Vortex the samples to mix.
94. Centrifuge the samples (800 G, 5 min, RT).
95. Decant supernatant.

**2.3 - Cell membrane permeabilization**

96. Vortex the samples to resuspend the cells in the remaining liquid volume.
97. Add 2 mL of MaxPar PBS to each sample (**NOTE:** The cells are washed with MaxPar PBS instead of MaxPar CSB in this step in order to prevent the aggregation of proteins from the MaxPar CSB in the methanol).
98. Vortex the samples to mix.
99. Centrifuge the samples (800 G, 5 min, RT).
100. Decant supernatant.
101. Add 100 µL of MaxPar PBS to each cell pellet.
102. Resuspend cell pellets to a homogeneous cell suspension by vortexing.

**NOTE:** Perform steps involving methanol and samples containing admixtures of methanol in a fume hood.

103. Add 2 mL of pure -20°C methanol (Sigma, Cat.No. 32213) to each sample.
104. Cap the round-bottom 5 mL tubes with a non-porous cap or laboratory film.
105. Resuspend cell pellets to a homogeneous cell suspension by vortexing.
106. Incubate the samples in methanol at -20°C for 10 min.
107. Add 2 mL of MaxPar PBS to each sample.
108. Mix the samples by vortexing.
109. Centrifuge the samples (800 G, 5 min, RT).
110. During centrifugation, retrieve aliquots of the intracellular staining antibody mix from -80°C and place on ice to thaw.
111. Slowly aspirate supernatant using a pipette down to approximately 0,5 mL (**NOTE:** Avoid using vacuum pumps for aspiration due to weak cell pellet cohesion in methanol).
112. Add 2,5 mL of MaxPar PBS to each sample.
113. Resuspend cell pellets to a homogeneous cell suspension by vortexing.
114. Centrifuge the samples (800 G, 5 min, RT).
115. During centrifugation, label a new round-bottom 5 mL tube for each sample.
116. Decant supernatant.

**NOTE:** Perform steps 117-122 on one sample at a time. The same pipette tip should be used when manipulating cells in order to mitigate cell losses. This pipette tip should be prevented from drying out.

117. Add 1 mL of MaxPar CSB to the sample.
118. Pipet the sample to resuspend the cells. (**NOTE:** This pipette tip should not be discarded and should be used for manipulating the cells in steps 119 and 121).
119. Transfer the sample to the corresponding round-bottom 5 mL tube through the cap filter.
120. Using a separate pipette (tip), add 1 mL of MaxPar CSB to the old 5 mL round-bottom tube.
121. Using the pipette tip from step 117, wash out the old round-bottom 5 mL tube with the 1 mL of MaxPar CSB and transfer the washout through the cap of the corresponding new round-bottom 5 mL tube.
122. Rinse the cap filter with 1 mL of MaxPar CSB.
123. Centrifuge the samples (800 G, 5 min, RT).
124. Decant away as much supernatant as possible without compromising the sample.

**2.4 - Intracellular staining**

**NOTE:** Perform steps 125-130 on one sample at a time. The same pipette tip should be used when manipulating cells in order to mitigate cell losses. This pipette tip should be prevented from drying out.

125. Gently vortex the sample to resuspend the cells in the remaining liquid volume.
126. Ensure there are no or few bubbles in the samples. Pulse the samples at 800 G three times to attempt to remove excess bubbles.
127. Carefully, not introducing bubbles, measure the volume of the resuspension using a pipette: Adjust the pipette to its lowest volume, pipet this minimal volume into the pipette tip, position the pipette tip at the bottom of the tube, and adjust the pipetting volume upwards up until the entirety of the sample is aspirated. Write down the volume on the pipette and pipet the sample back out into the tube (**NOTE:** This pipette tip should not be discarded and should be used for manipulating the cells in step 129).
128. Using a separate pipette (tip), prepare an aliquot of MaxPar CSB equal to the amount needed to bring the measured cell volume to [*N* * 25 µL]. Add either 10 µL or 10% of volume on top as a volume buffer.
129. Use the pipette tip from step 127 to adjust the volume of the cell resuspension with aliquoted MaxPar CSB to [*N* * 25 µL] (**NOTE:** Keep in mind the current volume of the sample, measured in step 127. To wash as many cells as possible out of the pipette tip, aspirate a volume of MaxPar CSB larger than the one measured in step 127 one or more times).
130. Vortex the sample gently to mix.
131. Spin down the intracellular staining antibody mix aliquots in a desktop centrifuge.
132. Add [*N* * 25 µL] of intracellular staining antibody mix to the samples. Save leftover antibody mix on ice for same-day staining of control beads according to the supplemental protocol "Staining of Control Beads" in the online supplemental appendix.
133. Vortex the samples gently to mix.
134. Incubate the samples at RT for 30 min. During incubation, either keep the samples agitated or vortex the samples periodically.
135. Prepare 3 mL of DNAse per sample and condition it to room temperature.

**NOTE:** Perform steps involving (para)formaldehyde and samples containing admixtures of (para)formaldehyde in a fume hood.

136. Prepare 1 mL of 250 nM Cell-ID™ Intercalator-Ir (Standard BioTools, Cat.No. 201192B) in 4% V/V paraformaldehyde per sample. For 1 mL of intercalator solution, mix 750 µL of MaxPar PBS, 250 µL of 16% paraformaldehyde solution, and 20 µL of 12,5 µL of Cell-ID™ Intercalator-Ir. Keep the solution in the dark at 4°C.
137. After the intracellular antibody incubation, add 3 mL of DNAse to each sample.
138. Vortex the samples to mix.
139. Incubate the samples in DNAse at RT for 5 min.
140. Centrifuge the samples (800 G, 5 min, RT).
141. Decant supernatant into a (para)formaldehyde-appropriate waste container.
142. Vortex the samples to resuspend the cells in the remaining liquid volume.
143. Add 3 mL of MaxPar CSB to each sample.
144. Vortex the samples to mix.
145. Centrifuge the samples (800 G, 5 min, RT).

146. Decant supernatant into a (para)formaldehyde-appropriate waste container.
147. Vortex the samples to resuspend the cells in the remaining liquid volume.
148. Once more, add 3 mL of MaxPar CSB to each sample.
149. Vortex the samples to mix.
150. Centrifuge the samples (800 G, 5 min, RT).
151. Decant supernatant.

**2.5 - DNA staining**

152. Vortex the samples to resuspend the cells in the remaining liquid volume.

**NOTE:** Perform steps involving (para)formaldehyde and samples containing admixtures of (para)formaldehyde in a fume hood.

153. Into each sample, add 1 mL of the 250 nM Cell-ID™ Intercalator-Ir solution prepared in step 136.
154. Vortex the samples to mix.
155. Incubate the samples at RT in the dark for 20 min.
156. Centrifuge the samples (800 G, 5 min, RT).
157. Decant supernatant.
158. Vortex the samples to resuspend the cells in the remaining liquid volume.
159. Add 2 mL of MaxPar CSB to each sample.
160. Vortex the samples to mix.
161. Centrifuge the samples (800 G, 5 min, RT).
162. Decant supernatant.
163. Vortex the samples to resuspend the cells in the remaining liquid volume.
164. Once more, add 2 mL of MaxPar CSB to each sample.
165. Vortex the samples to mix.
166. Centrifuge the samples (800 G, 5 min, RT).
167. Decant supernatant.

**2.6 - Freezing stained cells**

168. Ideally, the stained sample batches should be frozen at a maximum cell density of $1*10^7$ cells / mL. Therefore, adjust the volumes in the following protocol steps accordingly.
169. Prepare 1,5 mL of a solution of one part dimethyl-sulfoxide (DMSO) (Sigma, Cat.No. W387520) and nine parts fetal bovine serum (FBS) by volume (Sigma, Cat.No. F7524) (henceforth “FBS/DMSO”) for each cryogenic storage vial the stained samples will be stored in (i.e. 1350 µL of FBS and 150 µL of DMSO per sample).

**NOTE:** Perform steps 170-178 on one sample at a time. The same pipette tip should be used when manipulating cells in order to mitigate cell losses. This pipette tip should be prevented from drying out.

170. To the pelleted sample of stained cells, add 500 µL of FBS/DMSO for each cryogenic storage vial the sample will be split amongst.

171. Using the same pipette tip, gently resuspend the cell pellet in FBS/DMSO.
172. Using the same pipette tip, transfer the resuspended cells to cryogenic storage vials.
173. Using a different pipette (tip), once more, for each cryogenic storage vial the sample was split amongst, add 500 µL of FBS/DMSO to the round-bottom tube that contained the sample.
174. Using the pipette (tip) from step 170, use the added FBS/DMSO to gently wash the inner walls of the round-bottom tube that contained the sample.
175. Using the same pipette tip, transfer equal parts of the washout to the corresponding cryogenic storage vials.
176. Using a different pipette (tip), for a third time, for each cryogenic storage vial the sample was split amongst, add 500 µL of FBS/DMSO to the round-bottom tube that contained the sample.
177. Using the pipette (tip) from step 170, use the added FBS/DMSO to gently wash the inner walls of the round-bottom tube that contained the sample.
178. Using the same pipette tip, transfer equal parts of the washout to the corresponding cryogenic storage vials.
179. Store the vials at -80°C.

**NOTE:** Freezing a stained sample batch enables the data acquisition to be done at a later point in time.

### 3 - Preparation of stained cells for data acquisition by suspension mass cytometry

#### 3.1 - Thawing stained cells

180. Pre-label one round-bottom 5 mL tube for each cryogenic storage vial you plan to thaw.
181. Pre-label one round-bottom 5 mL tube for each sample batch you plan to thaw one or more cryogenic storage vials of.
182. Prepare 2 mL of DNAse per cryogenic storage vial you plan to thaw and condition it to room temperature.
183. Thaw the predetermined number of cryogenic storage vials of cells belonging to a barcoded and stained sample batch by floating the cryogenic storage vials in cold tap water.

**NOTE:** Perform steps 184-186 on one cryogenic storage vial at a time.

184. Transfer the contents of the cryogenic storage vial to the corresponding round-bottom 5 mL tube labeled in step 180.
185. Wash out the cryogenic storage vial with 1 mL of MaxPar CSB and transfer the washout to its corresponding round-bottom 5 mL tube.
186. Once more, wash out the cryogenic storage vial with 1 mL of MaxPar CSB and transfer the washout to its corresponding round-bottom 5 mL tube.
187. Centrifuge the samples (800 G, 5 min, RT).
188. Decant supernatant.

**NOTE:** Perform steps 189-199 on one set of round-bottom 5 mL tubes containing cells from the same sample batch at a time. The cells need to incubate in room-temperature DNAse solution for at least 5 minutes.

189. Vortex the samples comprising a batch gently to resuspend the cells in the remaining liquid volume.
190. Add 500 µL of DNAse to each round-bottom 5 mL tube.

191. Gently mix by pipetting.
192. Transfer all samples comprising the batch through the filter cap of the corresponding round-bottom 5 mL tube labeled in step 181.
193. Add 500 µL of DNAse to each old round-bottom 5 mL tube.
194. Gently wash the inner walls of the old round-bottom tube using the added DNAse and transfer the washout to the corresponding round-bottom 5 mL tube for that sample batch, through the filter cap.
195. Once more, add 500 µL of DNAse to each old round-bottom 5 mL tube.
196. Gently wash the inner walls of the old round-bottom tube using the added DNAse and transfer the washout to the corresponding round-bottom 5 mL tube for that sample batch, through the filter cap.
197. Count the cells in the round-bottom 5 mL tube containing the sample batch using your preferred cell counting method, using DNAse to dilute and resuspend samples in case of excessive cell density.
198. Label a number of round-bottom 5 mL tubes corresponding to the number of cells in the sample batch divided by $2,4*10^6$, rounded up (**NOTE:** When acquiring data using suspension mass cytometry, it is recommended to acquire only up to 3 mL of cell suspension at a time to avoid cells settling on the bottom of the tube, potentially increasing the event rate to the point of clogging the mass cytometer. Additionally, at an event rate of 400 events per second, which, with a sample intake rate of 30µL/min, corresponds to 800 cells/µL or $8*10^5$ cells/mL, a good number of stained cells to acquire per round-bottom 5 mL tube is $2,4*10^6$ cells).
199. Transfer the cells of the sample batch to the newly labeled tubes, dividing the volume equally among the tubes.
200. Centrifuge the samples (800 G, 5 min, RT).
201. Decant supernatant.

#### 3.2 - Conditioning stained cells for suspension mass cytometry

202. Vortex the samples gently to resuspend the cells in the remaining liquid volume.
203. Add 2 mL of MaxPar Cell Acquisition Solution (Standard BioTools, Cat.No 201240) to each round-bottom 5 mL tube.
204. Vortex the samples to mix.
205. Centrifuge the samples (800 G, 5 min, RT).
206. Decant supernatant.
207. Keep the cell pellets on ice until acquisition on the mass cytometer.
208. Acquire the cells resuspended in an appropriate volume of a 1:10 dilution of EQ™ Six-Element Calibration Beads (Standard BioTools, Cat.No. 201245) in MaxPar Cell Acquisition Solution Plus (Standard BioTools, Cat.No. 201244) ($8*10^5$ cells/mL).

**Staining of Control Beads**

**1 - Staining**

1. Determine how much volume is left in each of the antibody mixes (surface and intracellular). It is advised to have at least 5 µL of each antibody mix for staining beads.
2. If either of the antibody mix leftovers has a volume lower than 20 µL, determine which of the mixes has a smaller volume, then pipet that volume of both mix leftovers into the same 5 mL round-bottom tube. Otherwise, pipet a maximum of 20 µL of each of the staining mix leftovers into the same 5 mL round-bottom tube.
3. If less than 40 µL of antibody is present in the tube, adjust the volume to 40 µL using MaxPar Cell Staining Buffer (CSB) (Standard BioTools, Cat.No. 201068).
4. Add 10 µL of AbC™ Total Compensation capture beads (ThermoFisher Scientific, Cat.No. A10497-A) to the tube.
5. Gently vortex the tube to mix.
6. Incubate the beads in antibody for 30 minutes at room temperature (RT). During incubation, either keep the samples agitated or gently vortex the samples periodically.
7. Add 3 mL of CSB to the tube.
8. Vortex the tube to mix.
9. Centrifuge the tube (800 G, 5 min, RT).
10. Decant supernatant.
11. Vortex to resuspend the beads.
12. Once more, add 3 mL of CSB to the tube.
13. Vortex the tube to mix.
14. Centrifuge the tube (800 G, 5 min, RT).
15. Decant supernatant.
16. Vortex to resuspend the beads.
17. For a third time, add 3 mL of CSB to the tube.
18. Vortex the tube to mix.
19. Centrifuge the tube (800 G, 5 min, RT).
20. Decant supernatant.
21. If the beads will not be acquired on a mass cytometer on the same day, keep the stained beads pelleted in CSB overnight at 4°C and acquire them the next day.

**2 - Acquisition**

22. Vortex to resuspend the beads.
23. Add 2 mL of MaxPar Cell Acquisition Solution Plus (CAS+) (Standard BioTools, Cat.No. 201244) to the beads.
24. Vortex the tube to mix.
25. Centrifuge the tube (800 G, 5 min, RT).
26. Decant supernatant.
27. Keep the beads pelleted in CAS+ until acquisition on a mass cytometer.

Acquire the beads in 0,3 mL of a 1:10 dilution of EQ™ Six-Element Calibration Beads (Standard BioTools, Cat.No. 201245) in CAS+.
